# How and why do Quality Circles work for General Practitioners - a realist approach

**DOI:** 10.1101/2021.11.07.21265092

**Authors:** Adrian Rohrbasser, Geoff Wong, Sharon Mickan, Janet Harris

## Abstract

**Objectives:** To understand how and why general practitioners in quality circles (QC) reflect on and improve routine practice over time. To provide practical guidance for participants and facilitators to implement and for policy makers to organise this complex social intervention.

**Design:** A theory-driven mixed method

**Setting:** Primary health care

**Method:** We collected data in four stages to develop and refine the programme theory of QCs: 1) co-inquiry with Swiss and European stakeholders to develop a preliminary programme theory; 2) realist review with systematic searches in MEDLINE, Embase, PsycINFO, and CINHAL (1980-2020) to extend the preliminary programme theory; 3) programme refinement through interviews with participants, facilitators, tutors and managers of quality circles; 4) consolidation through interviews and iterative searches for theories enabling us to strengthen the programme theory.

**Sources of data:** The co-inquiry comprised 3 interviews and 3 focus groups with 50 European experts. From the literature search we included 108 papers to develop the literature-based programme theory. In stage 3, we used data from 40 participants gathered in 6 interviews and 2 focus groups to refine the programme theory. In stage 4, five interviewees from different health care systems consolidated our programme theory.

**Result:** Requirements for successful QCs are governmental trust in GPs’ abilities to deliver quality improvement, training, access to educational material and performance data, protected time, and financial resources. Group dynamics strongly influence success; facilitators should ensure participants exchange knowledge and generate new concepts in a safe environment. Peer interaction promotes professional development and psychological well-being. With repetition, participants gain confidence to put their new concepts into practice.

**Conclusion:** QCs can improve practice, promote professional development, and psychological well-being given adequate professional and administrative support.

**Strengths and limitations of this study:**

- To our knowledge, this is the first published research that explains how and why general practitioners participating in quality circles may improve standard practice and their psychological well-being over time.
- The findings can be used to inform practice and policy decisions.
- The resulting theory relies on the detail and depth of the reports in the literature and the veracity and adequacy of the information participants revealed in interviews and focus groups.
- To mitigate the risk of selection bias if researchers choose underlying theories and synthesise them ad hoc, we used stakeholders’ mental model and programme documentation as our framework for analysis.

## Introduction

Quality circles (QCs) are made up of 6–12 health care professionals who regularly meet to reflect on and improve their standard practice. The terms Practice Based Small Group Work, Peer Review Group, Problem Based Small Group Learning, Practice Based Research Group, Quality Circle, Continuous Medical Education (CME) Group, and Continuous Professional Development (CPD) Group were used interchangeably and varied among countries. The labels suggest the basic, original intent of the group. We decided to use the umbrella term Quality Circle to describe all of them.^1^ In the UK and Europe, QC are commonly used by general practitioners (GPs) for continuous professional development (CPD). The focus of discussion is usually a critical evaluation of an aspect of quality which participants themselves identify as important to them. GPs seek to improve the quality of their care by linking evidence to practice, learning to deal with uncertainty, discussing and reflecting on practice issues.^2^ Participation in QCs can raise self-esteem create a sense of belonging and improve psychological well-being in GPs.^1^ QCs may be especially helpful in crisis situations like the current Covid-19 pandemic, where working continuously under high pressure can undermine the professionalism and mental health of GPs.^3^

QCs can improve prescription patterns and diagnostic habits, whilst enhancing professional development and psychological well-being, but the results of randomized controlled trials are inconsistent and offer only limited behavioural explanations for these positive effects. As a complex social intervention, QCs combine didactic methods like brainstorming and reflective thinking with quality improvement (QI) techniques like audit and feedback or purposeful use of local experts. Their activities must be tailored to address local problems in primary health care (PHC) that participants want to solve.^4 5^ Our understanding of QCs is incomplete, and we need to learn more about these complex social interventions and their context-dependent outcomes and effects. This study seeks to clarify the contexts in which QCs are conducted, when they change GP behaviour and improve psychological well-being and why. We intended to develop a programme theory for QCs that explain show and why they work, with the aim of creating a common language and understanding,^6 7^ to engage stakeholders in discussions about improving QC processes in a participatory way and prepare the ground for further empirical testing.^8 9^ Our end goal was to develop an initial set of policy recommendations for setting up optimal QC processes and maintaining them. ^10-12^

## Methods

A project protocol was registered with PROSPERO (CRD42013004826) and published in 2013.^5^

We answered our research question in four stages. In stage one, we conducted a co-inquiry with stakeholders on QCs from Switzerland and other European countries, in which we narrowed the research question and provided a preliminary programme theory. In stage two, we synthesised evidence from a literature review and built a literature-based programme theory. In stage three, we collected evidence from interviews and focus groups with QC participants, facilitators, tutors, and managers and refined the programme theory. In stage four we consolidated our work, integrating interview data with participants across Europe and examining existing theories.

We conducted this research between 2013 and 2020, when the first author (AR) was completing his DPhil (PhD) project at the University of Oxford. AR’s thesis research engaged key Swiss and European experts and stakeholders at all stages; these were QC participants, facilitators, tutors, managers, and policy makers. The different players shared their perspectives when we developed the research questions, methods and analysis, and when we considered the implications of the results.

Pawson and Tilley’s realist logic was used to analyse the collected data because this form of analysis addressed the complexity of QCs as an intervention. ^13 14^ We sought to provide an in-depth explanation of QCs that showed how mutual learning in a social context improves standard practice and raises professional self-esteem, and increases well-being. The realist approach examines causal explanations of outcomes and then expresses them in their simplest form: context (C) ‘triggers’ or ‘activates’ a mechanism (M) that produces an outcome (O). These context-mechanism-outcome (CMO) configurations are ‘mini’ theories situated within a programme theory.^15^ As we develop CMO configurations, we can more clearly see the contexts that produce desired outcomes. Once we identify these contexts, we can more easily select activities to change a given context to match our desired outcome.

## Ethics Approval

The project was approved by the Central University Research Ethics Committee in Oxford (MSD-IDREC-C1-2015-002); it fulfilled the requirements of informed consent: handling of personal information and confidentiality conformed to the operational principles of the Declaration of Helsinki and adhered to the Belmont Report principles mandating respect for persons, beneficence, and justice.

## Stage one: the co-inquiry

From May to December 2013, we consulted with stakeholders and experts from Switzerland and with the European Society of Quality and Safety in Family Medicine (EQuiP). They shared their perspectives on our research questions and helped us construct a mental model of QCs function. For characteristics of participants, see supplemental material 1. We also collected information from QC programme documentation and training materials, extracting QC aims, detailed objectives, and roles. This preliminary programme theory built the framework for our realist review.

### Stage two: realist review

#### Searching for evidence

Our search strategy was informed by an earlier scoping review that reported on the intentions and benefits, historical development, and spread of QCs.^1^. In collaboration with a librarian, we refined our search strategy, combining terminology like ‘Programme’, ‘Quality Improvement,’ and ‘Group’ terms with a PHC search filter.^16^ We ran the search in Medline, EMBASE, PsycINFO, and CINHAL, without language restrictions (supplemental material 2) from 1974, to reflect the emergence of QCs in 1974, at McMaster, Canada, and in 1979 at the University of Nijmegen, the Netherlands. We conducted the search in October 2013 and updated it in December 2020. We broadened the search by examining citations in reference lists and Web of Science and searched manually for closely related papers (kinship papers) that had contextual features and theoretical background similar to those found in the referring studies.^5^

#### Searching for theories

In principle, any theory that explained QCs was a candidate for our realist review, including those from psychology, social, or economic sciences. We first identified key components of QCs; these were theories that described groups, their dynamics within organisations, and the role of the facilitator. We searched for theories about motivation, learning, and behaviour change to inform professional development and improve quality of care. After this search we had identified 52 threads of theories across several levels. Since the overlapping theories were complex, we deviated from the original protocol and used the preliminary programme theory (stage one) as an organising framework.

#### Selecting articles

Criteria for inclusion were: 1) the studies focused on small group work, 2) took place in the PHC setting, and 3) had a quantitative or qualitative outcome. We managed search results in EndnoteX8. SM and JH each assessed half of the retrieved papers and AR examined them all. The authors resolved disagreements through discussion. AR updated the search and included papers published from November 2013 to December 2020. GW checked the process paper selection and interpretation of the new data.

We appraised the relevance and rigour of each paper’s contribution. Data were relevant if they helped us understand a specific element or thread of theory in the larger programme theory. Threads of theory were rigorous if they met three explanatory criteria: consilience (the theory accounts for most of the data), simplicity (the theory is straightforward, without exceptions) and analogy (the theory relates to already known principles). ^15 17 18^

#### Analysis and synthesis of the data

We created a data extraction framework from the preliminary programme theory and implemented it in Microsoft Excel. For each study, we extracted data on mechanisms, contexts, and outcomes (Table 1).^10^ At least two authors (AR, SM, or JH) reviewed extracted data and all authors reviewed the analysis and interpretation.

**Table 1.** Data analysis process throughout the study.

| Step | Description of the analytical step |
| --- | --- |
| **One** | We collected data on the following key elements of QCs:<br>➤ Outcomes<br>➤ Participant characteristics: who was doing what and why?<br>➤ Activities: what was being done and why?<br>➤ Implementation context: where and how were QCs implemented?<br>➤ Patterns of outcomes over time or intermediate outcomes. |
| **Two** | Outcomes: each intermediate outcome, or final outcome received a new code. |
| **Three** | To identify the components of CMO configurations, we linked activities to intermediate outcomes, or final outcomes, and noted any corresponding contextual features and mechanisms that were mentioned. |
| **Four** | We linked activities to outcomes and sought explanation for when and why they had these outcomes (if the source mentioned context or underlying reasoning or mechanism) and then built CMO configurations. |
| **Five** | We categorised and ordered the CMO configurations to create a chain of outcomes and explained how CMO configurations related to each other. |
| **Six** | We compared and contrasted patterns identified in different sources. |
| **Seven** | We formulated, revised, and consolidated the programme theory foundation of quality circles. |

Initially, for each paper, we extracted components of context along with descriptions of mechanisms that led to an outcome. We summarised these configurations into descriptions of interaction between context and mechanisms to either facilitate or constrain QCs. Since papers were often closely related, we grouped them based on their kinship, which helped us look for and confirm CMO configurations between papers within the same (family) study. We iteratively arranged and rearranged the CMO configurations, moving between the papers, their data, and families, and built semi-predictable patterns of outcomes (demi-regularities) to develop the programme theory (see supplemental material 3).

### Stage three: refining the programme theory

AR conducted interviews and focus groups to refine and test the configuration, interpretation, and underlying mechanisms of each CMO configuration and its relative position/contribution to the programme theory.^19^

We invited a broad range of participants to participate in interviews, including experts and stakeholders from stage one, so we could capture a range of professional backgrounds and roles. Those we invited included tutors who train QC facilitators, facilitators who guide small groups, participating GPs, and QC managers.^20 21^ We applied the concepts of data saturation and stopped collecting data when additional information added no further relevant evidence. None of the invited participants declined. Throughout the process, we reflected critically on assumptions that AR or participants might have made during the interviews or focus groups.^20^ AR conducted six 30–60-minute interviews in Swiss German between March and May 2015. After explaining the literature-based programme theory in plain words, AR offered contrasting options for participants to discuss and then asked them to share their understanding of the underlying reasoning for QC interactions.

In April 2015, during an EQuiP conference, we held two focus group sessions with GPs from over 19 European countries. Participants were given written descriptions of the emerging programme theory, phrased as conditional clauses that did not suggest mechanisms. During the focus group, participants were asked if and how much they agreed with the statements, and then the group discussed whether and why parts of the programme would or would not work in certain contexts. We summarise the characteristics of interview and focus group participants in supplemental material 4.

### Stage four: consolidating the programme theory

To consolidate the programme theory, AR invited representatives from countries with different PHC provision systems to a one-hour online interview to discuss the ways that different professional associations, institutional settings, and other contexts affect QC outcomes.^21^ Participant characteristics are summarised in supplemental material 5.

We then compared and contrasted this emerging programme theory with formal theories to explain intermediate and final QC outcomes. Formal theories capture a programme theory’s underlying mechanisms and explain how its threads weave into patterns across different disciplines. Programme theories that are based on formal, existing, theories may also provide better explanations of phenomena than those that are not.^7^ Our candidate formal theories came from four sources: the scoping review;^5^ the realist review; theories described by interviewees; and theories identified during iterative searches when we were looking for and testing possible mechanisms. We chose theories with the highest level of explanatory coherence, based on the three criteria of consilience, simplicity, and analogy.^17 18^

## Results

### Stage one: the co-inquiry

50 QC experts and stakeholders narrowed the research question and provided data in three interviews and three focus groups, and also provided programme documentation and training materials. The interviews and focus groups sessions took place after the meetings of professional associations (Swiss Society of General Internal Medicine). This co-inquiry resulted in the following preliminary programme: 6–12 health-care professionals meet regularly to reflect on and improve their standard practice, employing didactic methods and QI techniques to identify gaps in their knowledge. Two fundamental concepts shape QCs from the beginning. The first is the cycle of learning, or QI, and the second is the social context in which the group functions. We have described in detail the CMO configurations we developed in this stage in supplemental material 6.

#### Stage two: realist review

Our search strategy returned 2,812 results (Figure 1), out of which AR, JH, and SM assessed 73 papers. An update in December 2020 yielded 35 more papers.

**Figure 1.**
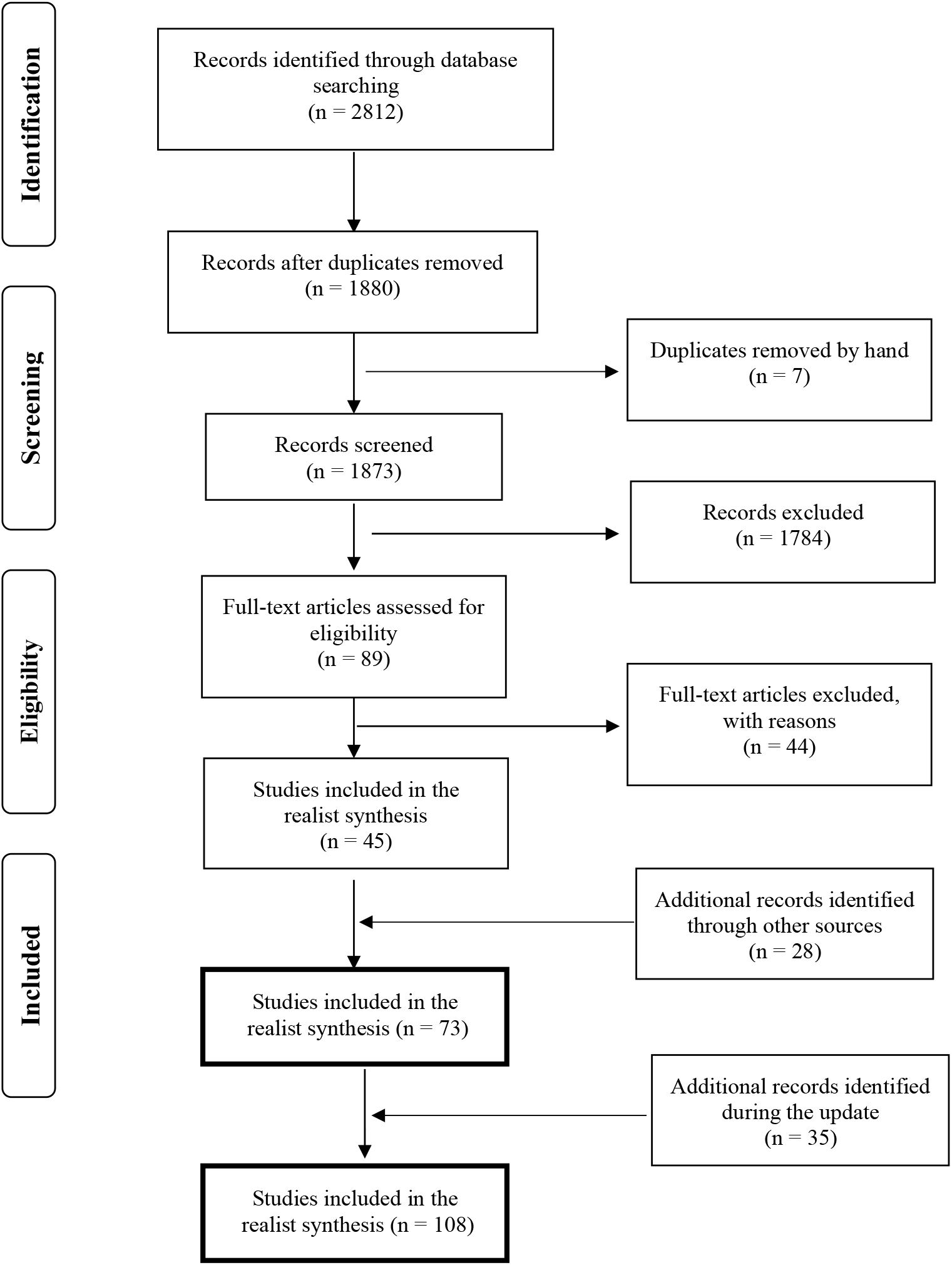
Paper flow realist review.

The literature mainly covered QCs in which GPs participated. We found 24 relevant articles about German QCs, 12 about Dutch QCs, and two about Swiss QC; 10 papers were about CME groups in Canada and Scotland, 6 about a QC research project in Norway, 3 about QCs on osteoporosis in Canada, and 5 about the Drug Education Project in Sweden, Norway, the Netherlands and Slovakia; 6 papers covered QC projects in England, Austria, Belgium and France; 5 other relevant papers were from South Africa, the US (Hawaii and California), New Zealand, and Australia. We categorised these papers into groups to clarify their kinship network, including an underlying trial, common themes, common contexts like geographical area, and common methods of organising QCs (e.g., papers that tested similar didactic methods or similar QI tools in QCs).

Study designs varied by research question. Our search returned 5 study protocols, 2 case series, 14 before-and-after studies, 13 controlled before-and-after studies, 9 randomised controlled trials, nine cluster randomised controlled trials, 12 surveys and 9 qualitative research papers that used data from interviews or focus groups. Few papers studied the performance of well-established QCs; data were often limited to interventions in newly formed groups. In pre-existing QCs (German, Dutch, or Norwegian trials), researchers introduced their own interventions on prescription or test-ordering patterns rather than studying interventions chosen and designed by the QC group. For full details of study characteristics, see supplemental material 7. We present the literature-based programme theory and supporting quotations from the literature in supplemental material 8. The data we retrieved from the update search did not change our CMO configurations or programme theory.

### Stage three: the refined programme theory

We used data from 40 participants, collected during six interviews and two focus group sessions held at the EQuiP meeting in Fischingen, Switzerland. For each CMO configuration, we tested its configuration, interpretation, underlying mechanism, and time relationship to others. We refined the wording of six CMO configurations and added three new configurations that linked the chains of outcomes. See supplemental material 9 for the resulting intermediate programme theory and supporting quotations and data from focus group sessions.

### Stage four: consolidating the programme theory

We consolidated the intermediate programme theory and explored its contextual layers during interviews with participants from five European countries. Interviewees provided rich data and detailed descriptions about what they deemed necessary preconditions for successful QCs and added an additional CMO configuration (1b ‘being embedded in a QI system’). For supporting quotations during these interviews, see supplemental material 10. Figure 2 shows the final CMO configurations of the consolidated programme theory (iteratively developed from stages one to four).

**Figure 2.**
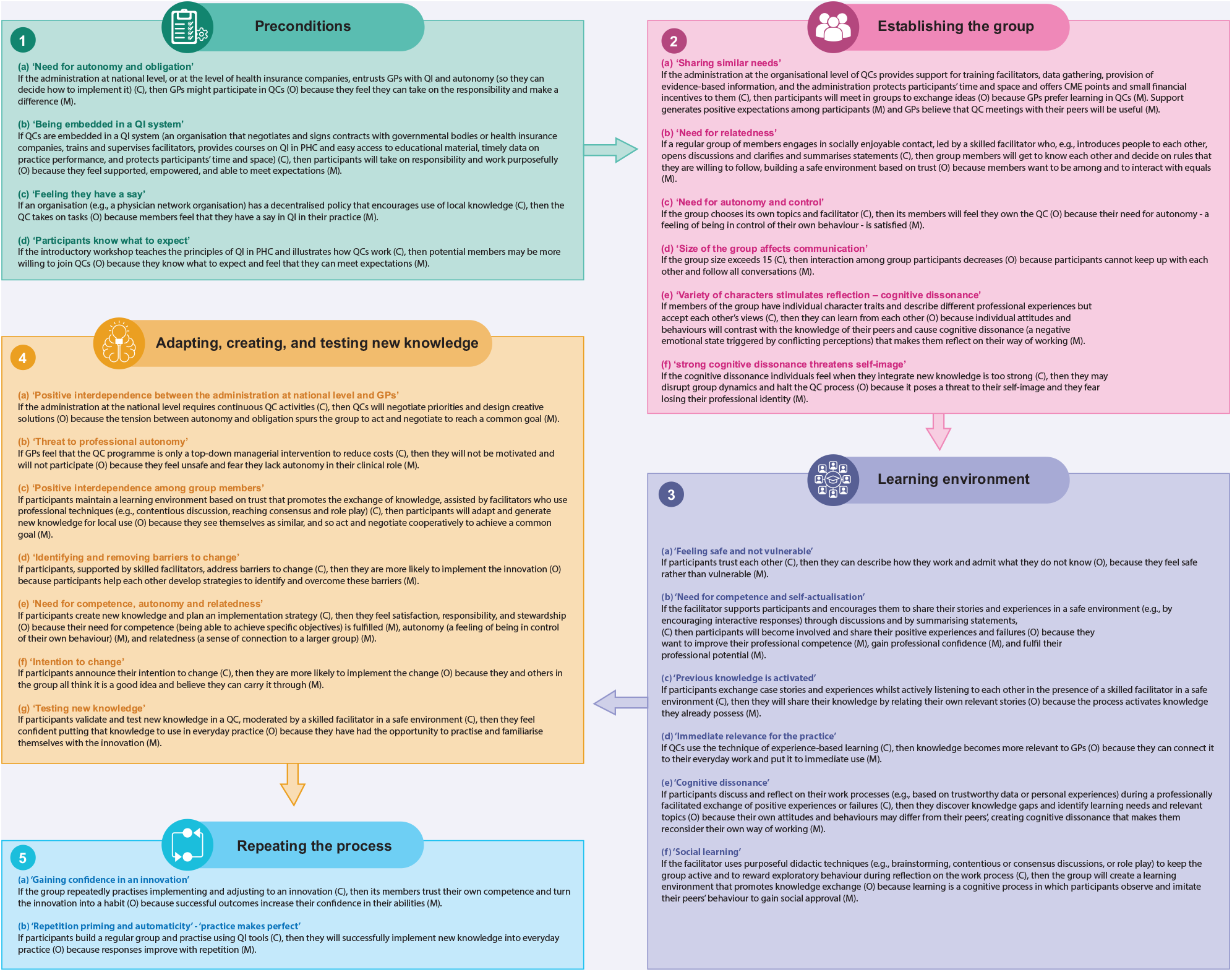
Consolidated programme theory on quality circles.

To further consolidate the programme theory, we used our candidate formal theories we found during the research process. Some theories about organisational context, groups, learning, knowledge exchange, development of innovations and their implementation were relevant. Some CMO configurations fit well with, or are directly supported by, existing theories, whilst others seem to clarify how existing theories work when they are applied to QCs. Table 2 summarises the theories and their corresponding CMO configurations.

**Table 2.**
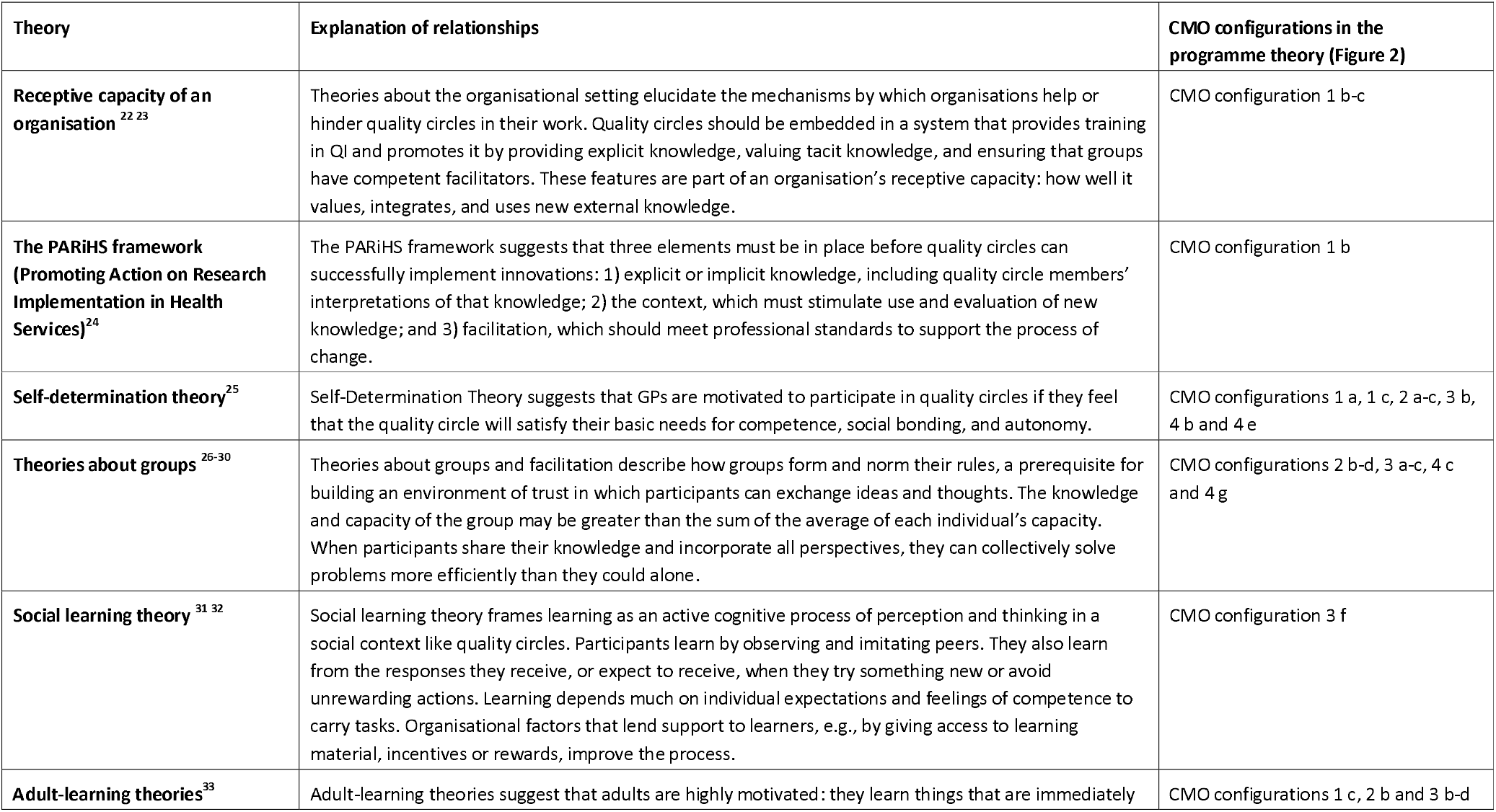
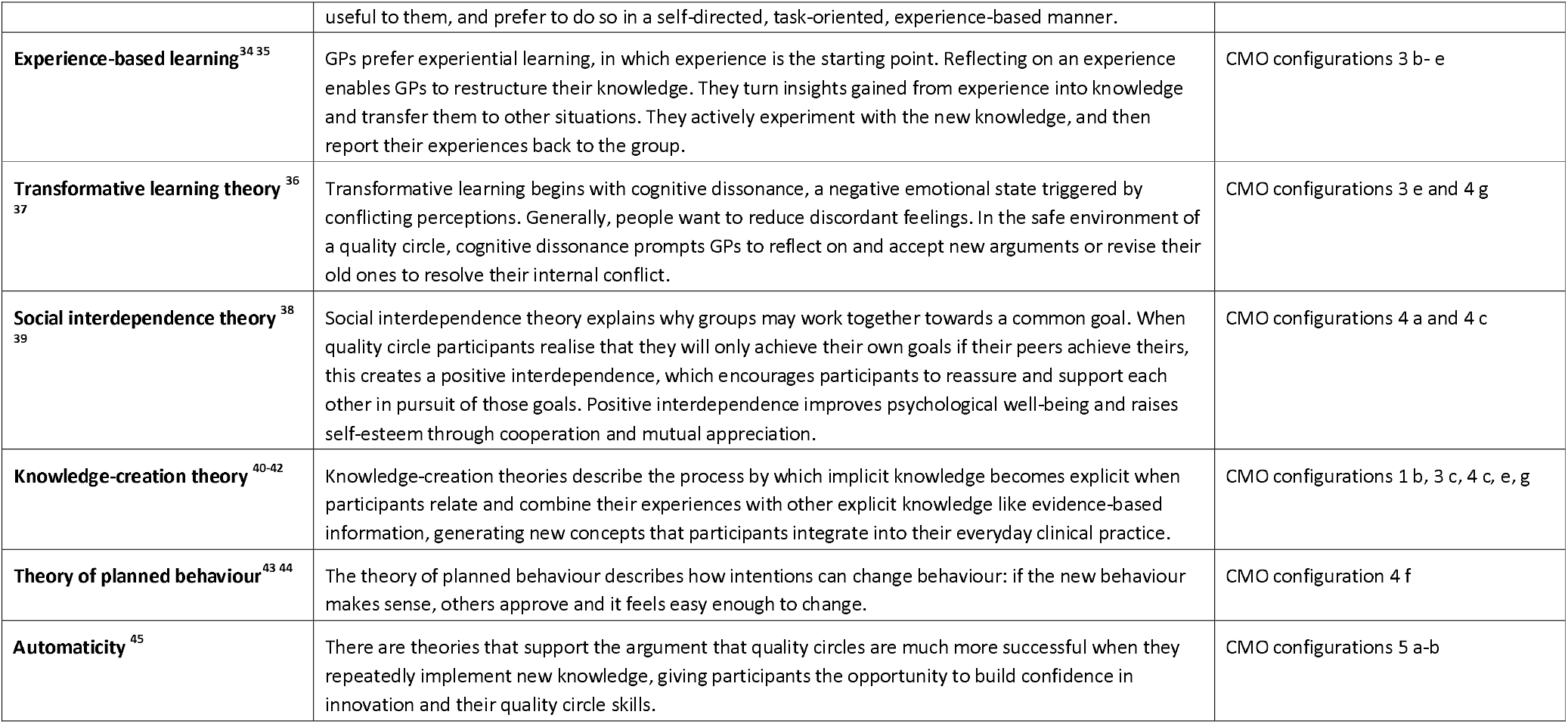
CMO configurations from the programme theory and their relationships to existing theories.

## Discussion

### Summary of the consolidated programme theory

The most important contextual requirements for successful QCs are governmental trust in the ability of GPs to deliver QI and appropriate professional and administrative support for QC work. Professional support includes training in QI techniques, easy access to teaching materials, and trustworthy personalised performance data. Administrative support includes providing protected time, an appropriate venue, and financial resources for meetings. If QC groups are to be successful, participants must feel that they have a say in their CPD and QI work, but the additional workload from participating in QCs must be manageable.

Several factors in QCs influence practitioner performance. QC members and their group dynamics are at the core of the process. Facilitators help participants build social bonds and mutual trust so that the QC becomes a safe environment that fosters open discussions and where participants link insights to everyday practice, manage uncertainty, and develop their professional role. Members reflect on personal experiences, add information from relevant sources, including evidence-based information and personal performance data, and then develop new ideas and concepts to improve their practice. With skilful facilitation, participants work towards a common goal and test their new ideas in the group, knowing that success depends on the individual member contributions. The QC process raises self-esteem and fosters psychological well-being. QI is cyclical, so putting innovations into practice is a continuous and repetitive process that increases participants’ confidence in their innovation and QI skills with each repetition.

### How the programme theory contributes to our understanding of QCs and relates to existing QC literature

Our understanding that QCs should be embedded in a system of QI that values, integrates, and uses new external knowledge aligns with the existing literature. ^23 46^ Health systems should provide training in QI tools and give access to trustworthy data (explicit knowledge) that help participants identify their own learning needs (CMO configuration 1 b-c and 3 e in Figure 2). ^22 47-50^

Our research confirmed that well-functioning groups are essential to the QC process. The group’s capacity for problem-solving surpasses the ability of individual when members share and pool their experiences and views ^29 48^. Supportive facilitation in a non-threatening environment of mutual trust eases learning in the group and opens possibilities for sharing, creating and integrating new knowledge. ^23 48 51-53^ Trust implies that participants operate on the basis of equality and mutual respect, according to the principle of benevolence, when they take risks and participate actively in the group (CMO configurations 1 c, 2 b 3 a-c, 4 c and 4 g in Figure 2). ^26 54^

We had several insights that had not been reported in current QC literature. Cognitive dissonance, like conflicting attitudes, beliefs or behaviours that create unease, is a mechanism that compels GPs to reflect on, accept, and adopt new reasoning to resolve inner conflict. According to our interview data, GPs can risk doing this in a QC group where they feel safe and confident, a process described in educational literature (CMO configurations 3 e and 4 g in Figure 2).^55-59^

Our data show that reflecting on an experience enables GPs to restructure their knowledge for transfer to other situations. When they share knowledge and experience, they can validate their clinical reasoning and thus integrate tacit and explicit knowledge and develop professional values like integrity and empathy; this process is recognised in the literature on psychology of learning as important to professional development.^60 61^ Explicit knowledge can be easily expressed through language or in writing because it is factual, e.g., evidence-based information, or a measurement of practice performance; whereas implicit or tacit knowledge is embodied in the knowledge or skills that a GP accumulates through experience but may find difficult to communicate. ^62^ CMO configurations 3 b-e, 4 g, and 5 a (in Figure 2) show GPs’ need for tangible experiences and repeated attempts to absorb new knowledge.^37^

According to our data, the mechanism of positive interdependence explained how and why collective or social learning can flourish and create a sense of ownership in QCs. When QC participants realise that they will only achieve their own goals if their peers achieve theirs, they are encouraged to reassure and support each other. Peers create new ideas and the cooperation and mutual appreciation that results improves their psychological well-being, increases their self-esteem, and may reduce their risk of burnout (CMO configurations 4 a and 4 c, e in Figure 2).^1 39 63-65^

Participants relate and combine their experiences with other explicit knowledge and generate new concepts or improve quality of care — a process described in business literature as knowledge creation.^40-42 60 66-68^ A key function of QCs is to merge familiar knowledge, local context, and personal experience with evidence-based knowledge and extend this from the micro view of single-patient care to a wider view of the whole system (CMO configurations 3 c, 4 e, 4 g and 5 g in Figure 2).

The literature, data from the realist review, and our interview data together suggest that participants may change their behaviour if it makes sense to do so, if others approve, and if change is not too demanding.^69^ But to embed these behaviour changes in everyday practice, the QC processes must be repeated, especially during the phase when GPs are implementing new knowledge, ^70 71^ (CMO configurations 4 f, 5 a and 5 b in Figure 2).

### Implications for policy and practice

Based on our findings, we summarised the recommendations for organising and performing QCs to increase the likelihood that GPs successfully improve the quality of their work (Figure 3). Each recommendation is based on one or more CMO configurations. Not all recommendations will apply to every QC. These recommendations should be considered as a form of decision support that QCs can draw on to determine if action is needed in their specific circumstances.

**Figure 3.**
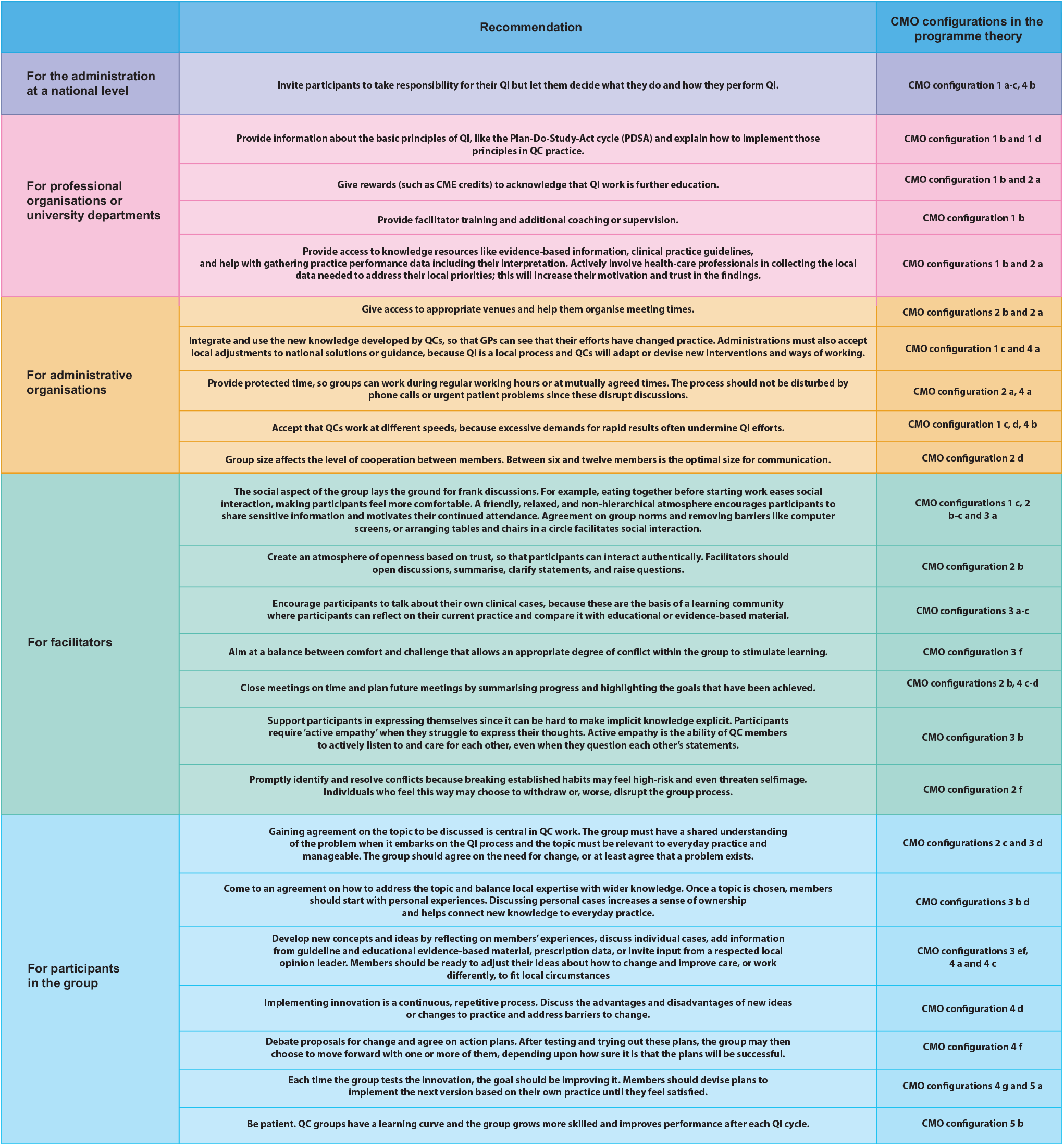
Recommendations and principles for organising successful quality circles.

The QC process and its implications are summarised as an infographic in supplemental material 11.

### Limitations

These realist approaches have two major limitations. First, the resulting theory relies on the detail and depth of the reports we identified in our literature review. To ensure we searched broadly, we looked for related reports and papers (kinship papers) including qualitative papers and evaluations that discussed different aspects of the research project and proposed other possible explanations for their findings. Our results also depend on the veracity and adequacy of the information participants revealed in interviews. To check the consistency and accuracy of this data, we relied upon sequential interviews to refine and consolidate our programme theory, step-by-step, as it emerged and interviewed groups of people with different perspectives (QC participants, facilitators, tutors, organisers and managers) to ensure our CMO configurations were adequate and clear. To mitigate the risk of social desirability bias, AR carefully posed neutral interview questions and tried to avoid embedding assumptions in his questions.

Second, the realist approach carries the risk of selection bias if researchers choose underlying theories and synthesise them ad hoc. To mitigate this risk, we used stakeholders’ mental model, programme documentation, and training material for facilitators to build the preliminary programme theory that served as our framework for analysis.

### Future research

Future researchers can build on this programme theory to design, implement and evaluate new QC interventions. We encourage researchers to test our programme theory to confirm, refute or refine it for specific settings and/or professional groups.

## Conclusion

Our consolidated programme theory explains how QCs can improve practice, foster professional development, and increase psychological well-being among participants. Group dynamics are at the core of the process. Facilitators help participants exchange knowledge in a safe environment where they generate new concepts to improve their practice. With repetition, QC participants gain confidence in their QI skills and put their innovations into practice. The requirements for successful QCs are 1) governmental trust in GPs’ abilities to deliver QI and appropriate support like professional facilitation, 2) training in QI techniques, 3) access to educational material and personal performance data; 4) granting protected time, appropriate venues, and financial resources for QC group members.

## Supporting information

Qualt Standards RAMESES

Ethics_1

Ethics_2

Consens sheet

Information sheet

## Data Availability

All data produced in the present work are contained in the manuscript

## Author Contributions

AR performed the research as part of formal postgraduate studies (DPhil Programme in Evidence-Based Health Care, University of Oxford, Oxford, UK). SM and JH supervised the development of the research and actively participated in the review process (eligibility, selection, data extraction followed by discussions). GW, as AR’s main supervisor, provided important input regarding the methodology and supervised the whole research process. All authors critically reviewed the text, assisted with editing read and approved the final manuscript. GW, SM and JH contributed independently to this project from their academic and methodological experience.

