## Supplemental material 1 for "How and why do Quality Circles work for General Practitioners - a realist approach"

**Experts and stakeholders participating in interviews and focus groups**

| Experts / groups | Type of qualitative data | Date and location | Length of time |
|---|---|---|---|
| Economist, CEO of Santémed network of primary health care centres owned by health insurance company | Interview | 01/05/2013 Zürich | 1½ h |
| GP, tutor in MediX Bern, a doctor-owned network of primary health care centres | Interview | 02/05/2013 Bern | 1½ h |
| Two social scientists, representatives of the professional body, Swiss Medical Association | Interview | 02/05/2013 Bern | 1½ h |
| 8 Participants, GPs, and a QC facilitator belonging to MediS Zürich, a doctor-owned network of primary health care centres | Focus group | 14/05/2013 Zürich | 1¼ h |
| GP and manager, facilitator and member of Sanacare's management board, a network of primary health care centres owned by health insurance companies | Interview | 15/05/2013 Bern | 1¼ h |
| 12 Representatives of the quality committee of the Swiss Society of General Internal Medicine, GPs | Focus group | 13/06/2013 Bern | 1h |
| 24 delegates of the European Society of Quality and Safety in Family Medicine, all GPs | Focus group | 16/11/2013 Bologna | 1h |
