## Supplemental material 2 for "How and why do Quality Circles work for General Practitioners - a realist approach"

**Supplemental material 2**
**Purposive search strategy in OVID Medline / EMBASE / PsycInfo**

### PRIMARY HEALTH CARE TERMS

1. general practice/ or family practice/
2. Primary Health Care/
3. general practitioners/ or physicians, family/ or physicians, primary care/
4. community health services/ or community health nursing/ or community mental health services/
5. (family adj3 (practice or practitioner* or physician*)).ti,ab.
6. (general adj3 (practice or practitioner* or physician*)).ti,ab.
7. (primary adj3 (care or healthcare)).ti,ab.
8. practice nurs*.ti,ab.
9. (community adj2 nurs*).ti,ab.
10. 1 or 2 or 3 or 4 or 5 or 6 or 7 or 8 or 9
11. Management Quality Circles/
12. quality circle*.ti,ab.
13. (group* adj3 (learning or work* or teaching or education*)).ti,ab.
14. (group* adj2 (intervention* or strateg* or program* or review*)).ti,ab.
15. (quality improvement* adj3 (intervention* or strateg* or program* or initiative* or tool*)).ti,ab.
16. (audit adj3 feedback).ti,ab.
17. peer review*.ti,ab.
18. reflective practice.ti,ab.
19. (learning adj3 (intervention* or strateg* or program* or initiative*)).ti,ab.
20. (education* adj3 (intervention* or strateg* or program* or initiative*)).ti,ab.
21. (continuing adj2 (education or development)).ti,ab.
22. Peer Review, Health Care/
23. medical audit/ or nursing audit/
24. exp Education, Continuing/
25. 11 or 12 or 13 or 14 or 15 or 16 or 17 or 18 or 19 or 20 or 21 or 22 or 23 or 24

### TERMS for QUALITY IMPROVEMENT

26. Quality Assurance, Health Care/
27. Total Quality Management/
28. Quality Improvement/
29. "Quality of Health Care"/
30. evidence-based practice/ or evidence-based medicine/ or evidence-based nursing/
31. Physician's Practice Patterns/
32. exp Professional Competence/
33. Guideline Adherence/

34. (quality adj3 (improv* or assurance or change)).ti,ab.

35. (practice adj3 (improv* or change)).ti,ab.

36. ((care or healthcare) adj3 (improv* or change)).ti,ab.

37. ((professional or physician* or medical or clinical or nurs*) adj competenc*).ti,ab.

38. ((guideline* or guidance or standard* or protocol*) adj2 (adhere* or complian* or concord* or implement*)).ti,ab.

39. (evidence based adj2 (practice or prescrib*)).ti,ab.

40. 26 or 27 or 28 or 29 or 30 or 31 or 32 or 33 or 34 or 35 or 36 or 37 or 38 or 39

41. Peer Groups/

42. Group Process/

43. Group Practice/

44. practice based.ti,ab.

45. 41 or 42 or 43 or 44

ADDITIONAL GROUP TERM

46. facilitator.ti,ab.

GROUP TERMS IN PRIMARY CARE

47. 10 and 46

PRIMARY CARE **AND** PROGRAM TERMS **AND** QUALITY IMPROVEMENT TERMS **AND** GROUP TERMS

48. 10 and 25 and 40 and 45

ADDING THE “QUALITY CIRCLES” **AND** “GROUP FACILITATION” IN TITEL **AND** ABSTRACT

49. 12 or 47 or 48

50. limit 49 to yr="1974 -Current"
