## Supplemental material 3 for "How and why do Quality Circles work for General Practitioners - a realist approach"

**CMO configurations across studies and demi-regularities**

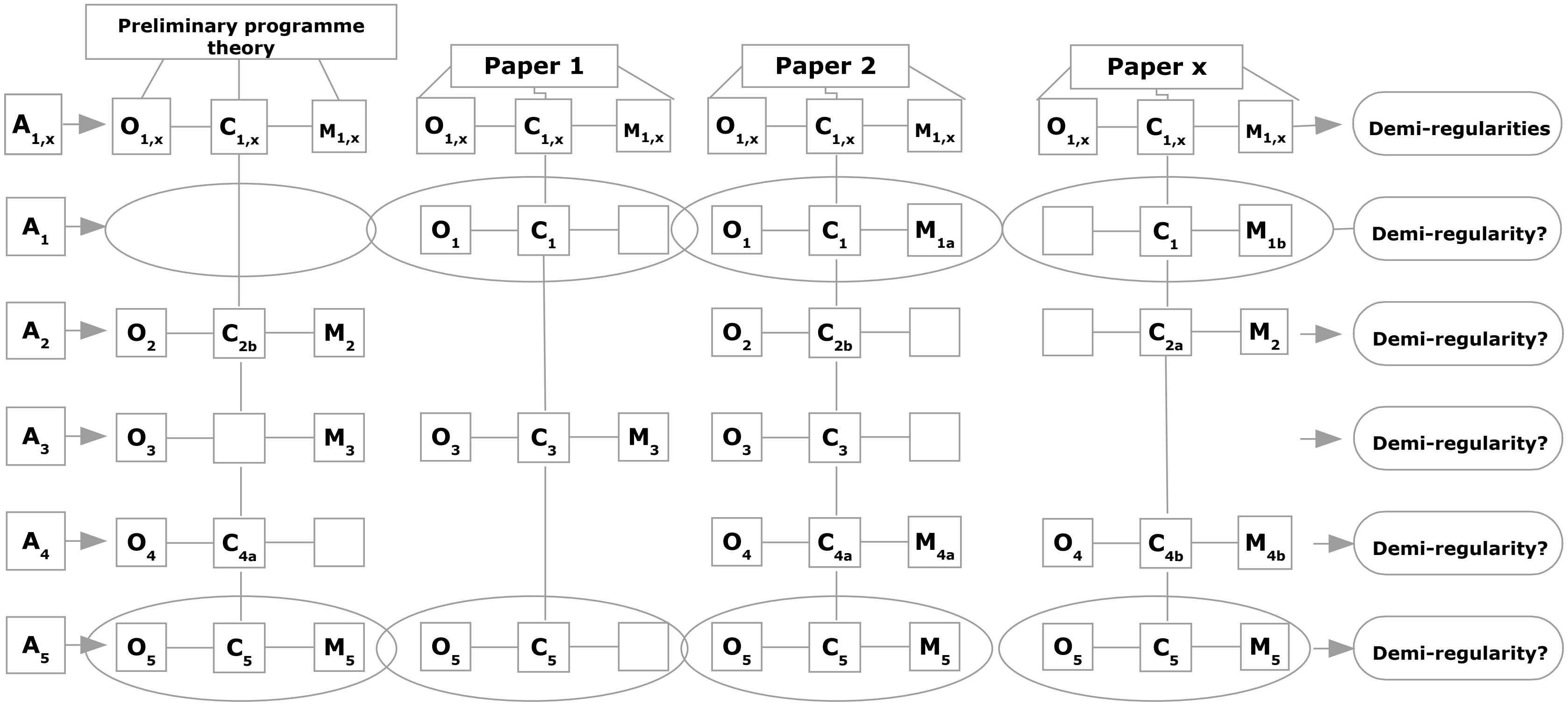


Legend
A: Activity. Activities 1 to 5.
O: Outcome. Outcomes 1 to 5.
C: Context. Contexts 1 to 5, although some may be missing.
M: Mechanism. Mechanisms at different levels of activity; the same outcome may have several mechanisms; if no mechanism was mentioned, the square is blank.
Comparing groups of CMOs across studies may create a demi-regularity.
The marked demi-regularities are examples of how I built CMO configurations across the papers.
