## Supplemental material 4 for "How and why do Quality Circles work for General Practitioners - a realist approach"

**Supplemental material 4 Participants in interviews and focus groups, refining the literature-based programme theory**

| **Participants: number of people** | **Type of qualitative data** | **Date and location** | **Length of time** |
|---|---|---|---|
| GP and manager, CEO of Santémed network of primary health care centres owned by health insurance company | Interview | 18/03/2015 Wil | 23’26’’ |
| GP, tutor in MediX Bern, a doctor-owned network of primary health care centres in Bern | Interview | 19/03/2015 Bern | 64’21’’ |
| GP and manager, facilitator and member of Sanacare’s management board, a network of primary health care centres owned by health insurance companies | Interview | 25/03/2015 Bern | 42’40’’ |
| Two social scientists, representatives of the professional body Swiss Association of Medicine | Interview | 02/04/2015 Bern | 72’47’’ |
| GP, facilitator of a QC belonging to MediS Zürich, a doctor-owned network of primary health care centres, Professor at the Institute of General Practice in Zürich | Interview | 02/04/2015 Zurich | 61’26’’ |
| Group of 21 GPs at the open meeting in Fischingen on QCs organized by EQuiP | Focus group | 24/04/2015 Fischingen | 90’ |
| Group of 12 GPs at the open meeting in Fischingen on QCs organized by EQuiP | Focus group | 25/04/2015 Fischingen | 90’ |
| GP, executive for General Practice at the hospital in Aarau | Interview | 08/05/2015 Aarau | 37’17’’ |
