## Supplemental material 5 for "How and why do Quality Circles work for General Practitioners - a realist approach"

Supplemental material 5 Participants in interviews designed to consolidate the programme theory

| Participant's characteristics | Country | Date of interview | Length of interview | Characteristics of the health care system |
|---|---|---|---|---|
| GP in a rural practice, teacher at the University of Ghent | Belgium | 07/02/2018 | 49' | Belgium's health-care system is funded principally through social insurance contributions on a fee-for-service basis, and these fees support doctors. The mandatory insurance can be replaced by a voluntary health insurance. Self-employed doctors provide the majority of outpatient services. GPs in Belgium are paid a small capitation fee but are not gatekeepers who refer patients to specialists. |
| GP in a rural practice, small group educator for 18 years | Ireland | 09/02/2018 | 64' | The Health Service Executive (HSE) funds the CME small-group network but the Irish College of General Practice (ICGP) has a governance role. The ICGP receives funds from the HSE to cover the majority of costs for tutors and group leaders, and some of the funds for group leaders come directly from the ICGP. At present, most of the funding is from the HSE. Ireland has a centrally organised PHC system which has a public and a private branch. GPs in Ireland are gatekeepers, with exceptions similar to France. |
| Certified facilitator in GP vocational training, active in quality improvement and patient safety | Norway | 08/02/2018 | 71' | Norway's health-care system is mainly public; insurance is covered by a percentage of income and tax subsidies. Almost everyone is registered with a primary care physician. The GP is the first point of contact for the patients and is a gatekeeper. Only 10% of GPs are directly employed by municipalities; 90% are self-employed and are licensed by their municipality, which guarantees them a basic salary. In addition, there are capitation fees for each GP. |
| GP working in an urban area, facilitating a QC, researcher | France | 16/02/2018 | 79' | Health insurance is compulsory in France. Depending on the occupational sector, workers pay a small proportion of their salary for their health insurance, with the employer paying the remainder of the cost; the amount depends on the worker's income. Immigrants and the unemployed have separate health insurance. As in Belgium, GPs in France are principally remunerated by a fee-for-service system. GPs are gatekeepers, except to paediatricians, gynaecologists and ophthalmologists. |

Supplemental material 5 Participants in interviews designed to consolidate the programme theory

| | | | | |
|---|---|---|---|---|
| GP in a rural practice, teacher for GP vocational training | Croatia | 07/12/2018 | 53' | All Croatian citizens are covered by the state health insurance fund. The health-care system is public and paid for by social security contributions. Despite financial constraints, the health-care system has expanded and covers the whole country, providing primary health care and specialised hospital-based care. GPs act as gatekeepers with certain exceptions, as in France. They are remunerated through a combination of salary, capitation fees and fee-for-service systems. |
