## Supplemental material 6 for "How and why do Quality Circles work for General Practitioners - a realist approach"

**Supplemental material 6**
**Preliminary programme theory based on the scoping search und consultations with stakeholders**

| |
|---|
| CMO configuration I: **'sharing similar needs'**<br>If health insurance companies require QI, and physician network organisations provide protected time and space, CME points and small financial incentives for participants (C), then GPs meet in groups to exchange ideas (O), because the organisational support generates positive expectations among participants and they believe these meetings with their peers will be useful (M). |
| CMO configuration II: **'size of the group affects communication'**<br>If group size exceeds 15 (C), then communication becomes difficult (O), because participants cannot keep track of so many people (M). |
| CMO configuration III: **'need for relatedness'**<br>If a steady group of members engages in socially enjoyable contact, led by a skilled facilitator who e.g. introduces people to each other, opens discussions and clarifies and summarises statements (C), then group members will get to know each other and decide on rules that they are willing to follow, building a safe environment based on trust (O) because members want to be among and to interact with equals (M). |
| CMO configuration IV: **'need for autonomy'**<br>If the group chooses its own topics and facilitator (C), then it has a sense of ownership (O), because this satisfies the need for autonomy and control (M). |
| CMO configuration V: '**need for competence and self-actualisation'**<br>If participants can tell their stories and experiences with the facilitator's support (e.g. encouragement of interactive responses and discussions, and summary of statements) in a safe environment (C), then they are involved in exchanging experiences and failures (O), because they want to be competent, gain professional confidence and fulfil their professional potential (M). |
| CMO configuration VI: **'previous knowledge is activated'**<br>If participants exchange case stories and experiences while actively listening to each other in the presence of a skilled facilitator (C), then they will be motivated to share their knowledge through telling such relevant stories (O), because the process activates the knowledge they already possess (M). |
| CMO configuration VII: **'cognitive dissonance'**<br>If participants discuss and reflect on their work processes during a professionally facilitated exchange of positive experiences or failures (C), then they become aware of knowledge gaps and identify learning needs and relevant topics (O), because conflicting attitudes and behaviours, together with differences between their own and other participants' knowledge, cause a cognitive dissonance (a negative emotional state triggered by conflicting perceptions) (M). |
| CMO configuration VIII: **'social learning'**<br>If the participants know what they need to learn or know what topic they want to discuss, and if they reflect on their own and other participants' trustworthy data (their own cases, diagnostic habits or prescription patterns, or evidence-based material such as guidelines) and if the facilitator uses purposeful didactic techniques (such as brain-storming, discussions and role play) to keep the group active and to reward exploratory behaviour (C), then the group will create a learning environment that promotes knowledge exchange (O), because learning is a cognitive process in which participants observe and imitate their peers' behaviour to gain social approval (M). |
| CMO configuration IX: **'interdependence between health insurance companies and physician network organisations/QCs; tension between autonomy and obligation'** |

**Supplemental material 6**
**Preliminary programme theory based on the scoping search und consultations with stakeholders**

| |
|---|
| If physician network organisations require continuous QC activities (C), then QCs will negotiate priorities and design creative solutions (O), because the tension between autonomy and obligation spurs the group to act and negotiate together to reach a common goal (M). |
| CMO configuration X: **'interdependence among group members'**<br><br>If participants maintain a learning environment based on trust that promotes knowledge exchange, assisted by facilitators who use professional techniques (e.g. contentious discussion, reaching consensus and role play) (C), then participants will adapt and generate new knowledge for local use (O), because they see themselves as similar, and thus act and negotiate cooperatively to achieve a common goal (M). |
| CMO configuration XI: **'gaining confidence in QC techniques'**<br><br>If the group repeatedly practises implementing and adjusting to an innovation (C), then they trust their own competence and turn the innovation into a habit (O), because successful outcomes increase their confidence in their abilities (M). |
| CMO configuration XII: **'repetition priming and automaticity'**<br><br>If participants establish a regular group and practise using QI tools (C), then they will successfully implement new knowledge in everyday practice (O), because responses improve with repetition: 'practice makes perfect' (M). |
