## Supplemental material 7 for "How and why do Quality Circles work for General Practitioners - a realist approach"

**Supplemental material 7 Study characteristics**

| Author year | Country | Study design | Set-ting | Participants, professional background | Study duration | Objective and intervention setting | Facilitation and group dynamics | Didactic and QI technique | Outcome oriented data | Common characteristics of the cluster (kinship) |
|---|---|---|---|---|---|---|---|---|---|---|
| **Norwegian papers on peer groups** | | | | | | | | | | |
| Gjelstad 2006[1] | Norway | Study protocol | PHC | 80 CME groups; 7–8 GPs in each group located in the southern part of Norway | 6 months: meetings once a month; the study covered 3 meetings. | Reduce prescription of antibiotics for upper respiratory tract infections and prescription of inappropriate drugs for elderly. Pre-existing CME groups. | Trained tutor serving 3 CME groups, reflection on own prescription strategies, disclosure of areas for individual improvement. | Discussions, reflective thinking on individual prescription data, one-day introductory workshop, audit and feedback, group educational outreach visits, academic detailing. | After one year, improvement of prescription patterns was expected. | Norwegian QC studies on improving drug prescriptions, accompanied by a qualitative study. Brekke provided the baseline study for the trial. |
| Gjelstad 2013[2] | Norway | Cluster randomised controlled trial | PHC | 80 CME groups; 7–8 GPs in each group located in the southern part of Norway | 6 months: meetings once a month; the study covered 3 meetings. | As in Gjelstad 2006<br><br>Each group acted as blind control for the other groups (Rognstad 2013). | As in Gjelstad 2006 | Authors consider the key element in the study to be 'what happens to a general practitioner's prescribing behaviour when they reflect on their prescriptions'. | After one year, reduction of prescription rate of antibiotics and increase of prescription rate of penicillin compared to control groups. | |
| Straand 2006[3] | Norway | Study protocol | Norwegian PHC | 80 CME groups; 7–8 GPs in each group located in the southern part of Norway | 6 months: meetings once a month; the study covered 3 meetings. | Reduce prescription of inappropriate drugs for elderly people and prescription of antibiotics in upper respiratory tract infections. Pre-existing CME groups. | Trained tutor serving 3 CME groups, reflection on own prescription strategies, disclosure of areas for individual improvements. | As in Gjelstad 2006 | After one year: reduction of inappropriate prescription patterns to elderly out-patients ≥ 70 years. | |

**Supplemental material 7 Study characteristics**

| **Author year** | **Country** | **Study design** | **Set-ting** | **Participants, professional background** | **Study duration** | **Objective and intervention setting** | **Facilitation and group dynamics** | **Didactic and QI technique** | **Outcome oriented data** | **Common characteristics of the cluster (kinship)** |
|---|---|---|---|---|---|---|---|---|---|---|
| Brekke 2008[4] | Norway | Cross-sectional study | PHC | 454 GPs in 80 CME groups, 85,836 patients | 6 months | Baseline data of ongoing CME groups for one year | Ongoing CME groups without intervention | Ongoing CME groups | After 1 year:18.4% of the patients received at least one inappropriate prescription. | Norwegian QC studies on improving drug prescriptions, accompanied by a qualitative study. Brekke provided the baseline study for the trial. |
| Rogn-stad 2013[5] | Norway | Cluster randomised controlled trial | PHC | 80 CME groups; 7–8 GPs in each group located in the southern part of Norway | 6 months: meetings once a month; the study covered 3 meetings. | As in Straand 2006<br><br>Each group acted as blind control for the other groups (Gjelstad 2013). | Training in drug treatment of elderly people, the rationale for the 13 listed inappropriate drugs, how to facilitate learning within a group setting. | Audit and feedback, tailored feedback, tailored academic detailing, discussions of own prescribing pattern. | After one year, reduction of inappropriate prescriptions for elderly people. Potentially more harmful combinations were more likely to be reduced. | |
| Frich 2010[6] | Norway | Qualitative study to explore experiences with academic detailing | PHC | 39 GPs and 20 tutors who were also GPs, 9 focus groups | 6 months: meetings once a month; the study covered 3 meetings. | Qualitative analysis of the RCTs, focusing on three meetings with the CME groups. | Groups have their own cultures; tutors perceived themselves as members of the group. | Consensus discussions, audit and feedback, academic detailing, discussions of their own cases. | Reflective thinking increased; inappropriate results upset some GPs. | |
| **Dutch papers on peer groups** | | | | | | | | | | |
| Geboers 1999[7] | The Nether-lands | Case series | PHC | All staff of 20 general practices (each working as a group) tested the model over a period of 18 months. | 18 months. Monthly quality meetings. | Evaluate the feasibility of a model for continuous quality improvement (CQI) in small practices. | Trained facilitators: practice assistants with managerial experience. Involving all staff at regular meetings. | Course on CQI: choose topic, observe practice, compare performance with targets, implement change, plan care and repeat cycle. | After 18 months, this model seemed feasible to the authors. | Dutch QC studies of a continuous quality improvement model. |

**Supplemental material 7 Study characteristics**

| Author year | Country | Study design | Set-ting | Participants, professional background | Study duration | Objective and intervention setting | Facilitation and group dynamics | Didactic and QI technique | Outcome oriented data | Common characteristics of the cluster (kinship) |
|---|---|---|---|---|---|---|---|---|---|---|
| Geboers 2001[8] | The Nether-lands | Mixed methods: before-and-after study and qualitative inquiry | PHC | 20 practices (each working as a group): 53 physicians and 57 medical practice assistants | 18 months. Monthly quality meetings. | Measure the attitude towards CQI model in small practices before and after study. | As in Geboer, 1999 | Feedback on practice assessment, introductory meeting, support for adoption of the model. | After 18 months, participants experienced perceived success and were willing to continue. | Dutch QC studies of a continuous quality improvement model. |
| Engels 2003[9] | The Nether-lands | Controlled before-and-after study | Mid-wives, mainly PHC | 255 midwives in 28 groups | Study period 1998 to 2000 | Measure CQI effect on clinical practice of midwives in PHC in a before-and-after study. | Three-day training of facilitators. Peer groups of midwives in the same geographical area. Regular group meetings. | Allocated topics with no choice, using the CQI model. | Positive effect on change of clinical practice was noted. Technical skills could not be improved. | |
| Engels 2006[10] | The Nether-lands | Ran-domised controlled trial | PHC | 26 sites in the intervention and 23 sites as controls. Size and composition of groups unknown. | December 2001 - February 2004; inclusion October 2001 - April 2003 | Examine the effects of a team-based model for CQI on primary-care practice management in small-scale practices. | Medical practice assistants as facilitators after 3 days' training. | Visitation Instrument for Practice (VIP) provided topics, CQI model with detailed oral and written feedback, monthly team meetings. | Evaluation after one year showed an increased number of CQI projects compared to control group, but the study was statistically underpowered. | |

**Supplemental material 7 Study characteristics**

| Author year | Country | Study design | Set-ting | Participants, professional background | Study duration | Objective and intervention setting | Facilitation and group dynamics | Didactic and QI technique | Outcome oriented data | Common characteristics of the cluster (kinship) |
|---|---|---|---|---|---|---|---|---|---|---|
| Verstap-pen 2003[11] | The Nether-lands | Multicentre, randomised controlled trial | PHC | 26 QCs consisting of 174 GPs. | 6 months baseline followed by 6 months' interventio n. | Determine the effects of a multifaceted strategy aimed at improving test ordering patterns in existing QCs. | Discussion and comparison of feedback reports among colleagues, communication course. | 3 consecutive, personal-feedback reports, comparison of results with guidelines, plans for change, discussion of Bayesian rules. | Modest improvement in test ordering when comparing the two intervention groups | Dutch QCs on improving test ordering |
| Verstap-pen 2004[12] | The Nether-lands | Cluster randomised controlled trial | PHC | 27 QCs consisting of 194 GPs. | 6 months of baseline followed by 6 months' inter-vention. | A multifaceted strategy aimed at improving test-ordering patterns in pre-existing QCs; 13 QCs followed a new strategy while 14 only received feedback. | Discussion and comparison of feedback reports among colleagues, communication training. 3 meetings. | As in Verstappen 2003 | Compared to feedback, the tailored intervention decreased test ordering significantly. | |
| Verstap-pen 2004[13] | The Nether-lands | Cluster randomised trial | PHC | 27 QCs consisting of 194 GPs. 13 QCs used a new strategy while 14 only received feedback | 6 months of baseline followed by 6 months' inter-vention. | Determine the effects of a multifaceted strategy in pre-existing QCs aimed at improving test ordering patterns. 3 meetings took place. | Discussion and comparison of feedback reports among colleagues, communication course. | As in Verstappen 2003 | Mean costs were reduced by cutting unnecessary tests. | Dutch QCs on improving test ordering |
| Verstap-pen 2004[14] | The Nether-lands | Cluster randomised trial; surveys | PHC | 27 QCs consisting of 194 GPs. Mean group size was 7.4 | 6 months of baseline followed by 6 months' | A process evaluation of a multifaceted strategy in pre-existing QCs aimed at | Discussion and comparison of feedback reports among colleagues using feedback in pairs, | As in Verstappen 2003 | Individual plans for change and group plan changes were made with a high level of | |

**Supplemental material 7 Study characteristics**

| Author year | Country | Study design | Set-ting | Participants, professional background | Study duration | Objective and intervention setting | Facilitation and group dynamics | Didactic and QI technique | Outcome oriented data | Common characteristics of the cluster (kinship) |
|---|---|---|---|---|---|---|---|---|---|---|
| | | | | | inter-vention. | improving test ordering patterns. | communication course. | | satisfaction. | |
| Smeele 1999[15] | The Nether-lands | Ran-domised controlled trial | PHC | 2 QCs, 17 GPs in each group | Pre-measure-ment and post-measure-ment after one year. | Evaluate the effects of a QC programme on guideline adherence. 4 sessions for GPs and 1 session for medical practice assistants. | The group education was conducted in two small groups with 9 and 8 GPs respectively. Facilitator was a GP. Not all GPs participated in all sessions. | Lectures, role-play, skills training, peer review of performance, group consensus discussions and problem-solving of hypothetical situations involving patients. | No significant changes were found for care provided and patient outcomes compared with the control group. | Dutch QC studies on guideline adherence. |
| Kasje 2006[16] | The Nether-lands | Cluster randomised trial using a balanced incomplete block design. | PHC | 10 peer review groups (97 GPs): chronic heart failure. 6 peer review groups (46 GPs): hypertension and diabetes mellitus type 2. | One educa-tional meeting followed by data collection after 6 months | Evaluate the effects of a QC programme on guideline adherence in pre-existing groups. One group received a programme on chronic heart failure, the other on diabetes mellitus type 2. | Facilitators adhered to a specific process. | One meeting: consensus about guideline statements, evaluation of current management of five of their own patients, listing barriers and possible solutions, formulation of personal intentions | No effect was shown. High dropout rate especially in the group dealing with diabetic patients. The programme was not implemented as intended. | |

**Supplemental material 7 Study characteristics**

| Author year | Country | Study design | Setting | Participants, professional background | Study duration | Objective and intervention setting | Facilitation and group dynamics | Didactic and QI technique | Outcome oriented data | Common characteristics of the cluster (kinship) |
|---|---|---|---|---|---|---|---|---|---|---|
| Van Eijk 2001[17] | The Netherlands | Randomised controlled trial with three parts: individual visits, group visits, and a control group. | GPs and pharmacists in PHC | Individual approach: 70 GPs and 14 pharmacists; Group approach: 52 GPs and 9 pharmacists in five QCs; Control: 68 GPs and their pharmacists. | 12 months | Comparison of individual educational visits versus group visits to improve inappropriate prescriptions for elderly people. Pre-existing groups of GPs. 3 visits at 4-month intervals. | There was no description about the process that took place in the groups or at the individual level. | First visit: guidelines about appropriate prescription of drugs for elderly.<br>Second visit: personal prescription habits were highlighted.<br>Third visit: short follow up | The individual and the group approach led to a reduction in the rate of starting inappropriate drugs and to an increase of prescription of appropriate drugs. | Dutch QC studies on improving drug prescriptions involving pharmacists. |
| Welschen 2004[18] | The Netherlands | Randomised controlled trial. | GPs and pharmacists in Dutch PHC | 12 peer review groups including 100 GPs with their collaborating pharmacists. | Approx. 6 months. Evaluation after 9 months. | Reduce prescription of antibiotics to patients with upper respiratory tract infections in pre-existing groups. | Group education with consensus procedure. One meeting followed by individual feedback after 2 weeks and 6 months. | Group education meeting about guidelines, communication skills training, patient leaflets. After 2 weeks and 6 months, individualised feedback. | Prescription rate for antibiotics was reduced after 9 months. After 15 months, the effect was lasting. Satisfaction among patients remained high. | Dutch QC studies on improving drug prescriptions involving pharmacists. |
| **Problem Based Small Group Learning (PBSGL) in Canada, Scotland and England** | | | | | | | | | | |
| Davis 1999[19] | Canada | Case series | PHC | 54 GPs in 4 newly formed groups. | A 2.5-hour workshop | Develop and evaluate a CME programme on osteoporosis for PHC. 54 family physicians participated in 1 of 4 pilot PBSG learning sessions. | GP trained as a facilitator. The facilitator elicited interactive responses using specific predetermined prompting questions. | Practice-based case scenarios to increase awareness of risk factors for osteoporosis. | Participants' satisfaction was high. Participants increased their knowledge scores (not significant because of size of the study). | Papers about Practice Based Small Group Learning in Canada, Scotland and England |

**Supplemental material 7 Study characteristics**

| Author year | Country | Study design | Set-ting | Participants, professional background | Study duration | Objective and intervention setting | Facilitation and group dynamics | Didactic and QI technique | Outcome oriented data | Common characteristics of the cluster (kinship) |
|---|---|---|---|---|---|---|---|---|---|---|
| Mc Sherry 2000[20] | Canada | Before-and-after study | PHC | 544 GPs in 75 workshops with a mean of 7 GPs in each newly formed group. | A 2-hour workshop with question-naires before and after. | Pilot study to introduce PBSGL groups in PHC. Topic: a patient-centred approach to managing benign prostate problems and evaluate 'intent to change'. | Initial needs assessment, problem-based educational materials, opportunities for participants to develop implementation strategies through discussion with peers. | Educational video case studies illustrating various presentations of prostatism, a handbook with detailed information on the case studies. A toll-free telephone line was provided for scientific and technical support. | Practice behaviours were improved, especially those linked to a patient-centred approach not commonly practised before the workshops. | Papers about Practice Based Small Group Learning in Canada, Scotland and England |
| Peloso 2000[21] | Canada | Qualitative inquiry over three years | PHC | 12–15 GPs, a facilitator and sometimes an expert. | 3 years | Discuss a 3-year experience with the small-group format, comprising more than 25 sessions as either learners or facilitators. Facilitators have 20 hours of training. Monthly meetings, each session takes 1.5 to 2 hours. | Sessions took place in the evenings with a meal in a relaxed atmosphere. The group chose their topics. Presentation of own clinical cases. Experts did not lecture but answered questions. | Learner-directed agenda of topics, information from trusted peers, opportunity for feedback. Information from several sources – printed materials, peer discussion, patient questions – the perception of need for change is enhanced. | GPs can discuss topics relevant to day-to-day practice and obtain access to local experts. They compare their practice with that of others. The group and the inter-active format are fun. Experts are comfortable with the format. | |

**Supplemental material 7 Study characteristics**

| Author year | Country | Study design | Set-ting | Participants, professional background | Study duration | Objective and intervention setting | Facilitation and group dynamics | Didactic and QI technique | Outcome oriented data | Common characteristics of the cluster (kinship) |
|---|---|---|---|---|---|---|---|---|---|---|
| Herbert 2004[22] | Canada | Ran-domised controlled trial | PHC | 200 GPs in 28 pre-existing groups. | 6 months, com-parison of data 6 months before and after the inter-vention. | Assess the impacts of individualised prescribing feedback. 4 groups: control, prescribing portrait only, educational module only, both portrait and educational module. | 3 representative patient cases were discussed, evidence-based information to guide management. Facilitation 'as usual' in the CME group. | Histograms comparing an individual's prescribing rates with those of the group and of all GPs in the study. A succinct evidence-based message to guide future prescribing. | The group that received both the module and the portrait had the greatest increase in preferred prescriptions. | Papers about Practice Based Small Group Learning in Canada, Scotland and England |
| Mc Vicar 2006[23] | Scotland | Before-and-after study (pilot) | PHC | 5 small groups, 7–9 GPs in each group | 12 months | Assess effectiveness of the PBSG approach in developing participants' knowledge, skills and attitudes in interpreting, discussing and applying current medical evidence. | Facilitators establish and maintain a learning environment. They create a culture of openness, honesty and willingness to acknowledge unawareness as a precursor to learning. | Educational material, a tool that triggers reflection, discussion of personal experiences and acknowledge-ment of gaps between current and best practice. | The study was statistically underpowered. Participants highlighted general enjoyment, professional reassurance and personal learning. | |

**Supplemental material 7 Study characteristics**

| Author year | Country | Study design | Set-ting | Participants, professional background | Study duration | Objective and intervention setting | Facilitation and group dynamics | Didactic and QI technique | Outcome oriented data | Common characteristics of the cluster (kinship) |
|---|---|---|---|---|---|---|---|---|---|---|
| Armson 2007[24] | Canada | Description of the programme | PHC | 4–10 GPs | Meeting of around 90 minutes once or twice a month | Identify gaps between current practice and best available evidence, to encourage reflection on individual practice, and promote changes in patient care, using an educational approach. | The facilitator's tasks are to focus discussion, to encourage the group to identify barriers to the implementation of new know-ledge and to establish a safe, supportive environment for learning. | Facilitation of discussions based on educational material and a tool (log sheet) that triggers reflection. The group starts with personal experiences and reflects on and acknowledges gaps between current practice and best practice. | Groups of various compositions function effectively in this particular small group environ-ment. If the facilitator lost the group's interest, disintegration of the group was likely. | Papers about Practice Based Small Group Learning in Canada, Scotland and England |
| Kelly 2007[25] | Scotland | Qualitative study: semi-structured interviews | PHC | One-to-one interview to evaluate the process in 5 small pre-existing groups. | Interviews among partici-pants of the Mc Vicar 2006 study | Explore the perceptions and experiences of PBSG participants to gain an understanding of how PBSGL works. | Facilitator opens discussions, clarifies statements, summarises what was said and questions issues, creating a learning environment. | Case discussions make evidence-based material relevant to participants and stimulate reflection. Mutual learning is important. Discussing data with others stimulates reflection. | Participants joined PBSGL groups because of the need to update medical knowledge, to compare personal practice with peer practice. | |

**Supplemental material 7 Study characteristics**

| Author year | Country | Study design | Set-ting | Participants, professional background | Study duration | Objective and intervention setting | Facilitation and group dynamics | Didactic and QI technique | Outcome oriented data | Common characteristics of the cluster (kinship) |
|---|---|---|---|---|---|---|---|---|---|---|
| Overton 2009[26] | Scotland | Qualitative approach: theory-driven framework developed by Chen and Rossi | PHC | 19 GPs and practice nurses | Interviews among partici-pants of PBSGL groups | Study the experiences of GPs and practice nurses in PBSGL. Data sources: logbooks, e-mail, telephone conversations and one-to-one interviews. | Qualitative study of the process in PBSGL groups: Group cohesion grew and mutual emotional support increased. With increasing trust, open discussions were possible. | Qualitative study of the process in PBSGL groups: case discussions kept people going and different perspectives could be considered. Self-esteem increased, as did mutual respect. | Motivation for joining the groups: preferred learning style, keeping up to date, learning in multi-professional groups, group atmosphere. and increased self-esteem. | Papers about Practice Based Small Group Learning in Canada, Scotland and England |
| Cunning ham 2011[27] | Scotland | Qualitative study: focus group | PHC | Two focus groups of PBSGL facilitators. | Focus groups | Learn about motivators to become a facilitator in PBSGL groups. | Qualitative study of the process in PBSGL groups | Qualitative study of the process in PBSGL groups | Motivators to become a facilitator were positive past experience of group learning, the chance of career advancement. Support for facilitators after initial training.t. | |

**Supplemental material 7 Study characteristics**

| Author year | Country | Study design | Set-ting | Participants, professional background | Study duration | Objective and intervention setting | Facilitation and group dynamics | Didactic and QI technique | Outcome oriented data | Common characteristics of the cluster (kinship) |
|---|---|---|---|---|---|---|---|---|---|---|
| Rial 2013[28] | England | Before-and-after study | Trai-nee GPs in PHC | 2 newly-founded groups of seven future GPs. | After 8 months, 4 meetings | Identify whether they were supported in making the transition from trainee to independent practitioner through attending PBSGL groups. | One group member was trained as a facilitator. | Canadian PBSGL approach was used | Improved ability to identify and use evidence in practice, shifting the focus from postgraduate exams towards 'real world' practice. The PBSGL groups still meet. | |
| **QCs in Canada** | | | | | | | | | | |
| Ioan-nidis 2007[29] | Canada | Before-and-after study (pilot) | PHC | 5 QCs, 52 physicians, GPs and some osteoporosis specialists | 12 months | Assess whether use of QCs could improve family physicians' adherence to osteoporosis guidelines. 3 training meetings for the facilitators, 3 meetings for participants. | QC facilitators were local family physicians recruited and trained specifically to lead study meetings. | Educational material, interactive group meetings, use of local opinion leaders, audit and feedback, reminders, multi-professional collaboration, financial incentives and information distributed to patients. | The intervention seemed to be feasible and was well received among GPs. 84% agreed that the feedback helped them understand their current practice patterns and decide on areas that needed improvement. | Papers on guideline adherence using continuous quality improvement cycles in Canada. |

**Supplemental material 7 Study characteristics**

| Author year | Country | Study design | Set-ting | Participants, professional background | Study duration | Objective and intervention setting | Facilitation and group dynamics | Didactic and QI technique | Outcome oriented data | Common characteristics of the cluster (kinship) |
|---|---|---|---|---|---|---|---|---|---|---|
| Ioan-nidis 2008[30] | Canada | Before-and-after study | PHC | 340 participants (GPs) in 34 QCs and local opinion leaders | 1 year | Increase guideline adherence concerning osteoporosis. 5 meetings (60–90 minutes) for two years. | 5 educational meetings | As in Ioannidis 2007 | Physicians' awareness of osteoporosis risk factors and appropriate bone mineral density testing increased. | Papers on guideline adherence using continuous quality improvement cycles in Canada |
| Ioan-nidis 2009[31] | Canada | Before-and-after study | PHC | As in Ioannidis 2008 | 2 years | As in Ioannidis 2008 | As in Ioannidis 2008 | As in Ioannidis 2008 | Guideline adherence increased | |
| **German QCs** | | | | | | | | | | |
| Szecse-nyi 1994[32] | Germany | Before-and-after study | PHC | 10 GPs | 2 years | Observation of the initialisation and establishment of a QC. Monthly meetings. | Presentation round, discussion of possible topics, choice of a topic impacting all participants; a GP facilitates the process. | Setting priorities, analysing the situation, developing criteria for improving quality, analysis of present practice, general priorities for necessary changes, comparison with evidence-based literature, change of practice. | GPs are interested in everyday practice-related topics. The gap between existing knowledge and clinical practice is acknowledged. | Papers about establishing QCs in Germany: pilot stage. |

**Supplemental material 7 Study characteristics**

| Author year | Country | Study design | Set-ting | Participants, professional background | Study duration | Objective and intervention setting | Facilitation and group dynamics | Didactic and QI technique | Outcome oriented data | Common characteristics of the cluster (kinship) |
|---|---|---|---|---|---|---|---|---|---|---|
| Gerlach 1995[33] | Germany | Survey among 138 QC participants | PHC | 138 GPs taking part in QCs, 8–12 GPs in each one. | Not applicable | Evaluation of case-based QC process focussing on a topic. | GPs use their own medical records, patient data or video recordings as a basis for problem-based learning. Facilitation by a GP. | Case-based discussions may indicate a need to change everyday practice. Evidence-based material and/or local opinion leaders may contribute to the discussion and consensus finding. | 79% of the GPs thought that cases from daily practice should be the starting point of QCs. The process led to locally adapted guidelines. | |
| Hart-mann 1995[34] | Germany | Controlled before-and-after study | PHC | 2 QCs, 10 GPs in each group compared to control group | 4 months. Evalua-tion after 5 meetings | Increase guideline adherence in diabetic care. Test training modules for facilitators (GPs). | 2 GPs in each group received training in facilitating small groups. | Didactic techniques as in Gerlach 1995, role play to practise patient–doctor communication. | Guideline adherence improved compared to control group. | Papers about establishing QCs in Germany: pilot stage. |
| Murad 1998[35] | Germany | Before-and-after study | PHC | 1 QC including 10 GPs | 12 months | Improve guideline adherence for patients with diabetes mellitus type 2. 23 existing QCs meeting once a month. | GPs use their own medical records, patient data or video recordings as a basis for problem-based learning. Facilitation by a GP. | Use of practice data, medical records and case discussions involving a local opinion leader. | According to QC documents, improved guideline adherence. | |
| Tausch 1995[36] | Germany | Before-and-after study (protocol) | PHC | 23 QCs, 10 GPs in each group | Evalua-tion over 18 months | Evaluate facilitators' manuals on different common diseases. 23 existing QCs met | The facilitators prompted and encouraged participants to identify common problems in their | The manual may provide a starting point for developing consensus guidelines. | Evaluation on three levels: reasons for participation in QCs, usability of the manual, | Papers about establishing QCs in Germany using manuals. |

**Supplemental material 7 Study characteristics**

| Author year | Country | Study design | Set-ting | Participants, professional background | Study duration | Objective and intervention setting | Facilitation and group dynamics | Didactic and QI technique | Outcome oriented data | Common characteristics of the cluster (kinship) |
|---|---|---|---|---|---|---|---|---|---|---|
| | | | | | | once a month. | practice. | | assessing behaviour change. | Papers about establishing QCs in Germany using manuals. |
| Tausch 1996[37] | Germany | Survey | PHC | 25 QCs, 246 GPs | Evalua-tion after 12 months and 10 meetings | Capture the objectives of the participants. 25 pre-existing QCs met once a month. | As above | Case vignettes, discussion of adequate diagnostic and therapeutic procedures in relation to evidence-based material. | Reasons for participating in QCs: exchange among colleagues, improved self-confidence. | |
| Tausch 2001[38] | Germany | Before-and-after study | PHC | 23 QCs, 243 GPs | Evalua-tion after 18 months | Evaluate reasons for participation, usability of manuals and assessment of behaviour change (self-reported improvement). To expand QCs within short time. | Voluntary participation in monthly meetings, 6–12 GPs in each group, trained facilitator. | Moderator-manuals that allow self-evaluation provide information about appropriate diagnostic and therapeutic recommendations for common diseases. | Reasons for participating: exchange of experiences among colleagues, increased competence and high level of satisfaction. | |
| Andres 1997[39] | Germany/ Hessen | Controlled before-and-after study | PHC | 32 GPs were grouped into 3 QCs promoted by the association of statutory health insurance | 12 months | Evaluate the process in the groups after 10 meetings. Participating GPs exceeded average prescription costs. | Participants felt forced to join QCs to change their behaviour. They had to overcome the feeling of being controlled. | Case discussions, audit charts to analyse prescription habits, interactive learning, reflective thinking and consensus finding as to rational prescription practice. | 66% reported change in behaviour. 22 of 27 wanted to continue with QCs. | Papers about establishing QCs in Germany using data on everyday practice to improve prescription patterns. |

**Supplemental material 7 Study characteristics**

| Author year | Country | Study design | Set-ting | Participants, professional background | Study duration | Objective and intervention setting | Facilitation and group dynamics | Didactic and QI technique | Outcome oriented data | Common characteristics of the cluster (kinship) |
|---|---|---|---|---|---|---|---|---|---|---|
| Andres 2004[40] | Germany/ Lower Saxony | Survey among 797 QC participants | PHC | 648 out of 797 participants answered the survey | Evalua-tion after 1 year | Evaluate QC participants' experiences in QCs intended to improve prescription patterns. | 7–10 GPs, monthly meetings, facilitator guiding through the process, support by academic staff members if necessary. | Case discussions, peer-led academic detailing allowing comparison with colleagues, reflective thinking, consensus discussions, evidence-based material, patient information. | Main problems were initial prescribing in hospitals and communication with patients when changing drugs. | Papers about establishing QCs in Germany using data on everyday practice to improve prescription patterns. |
| Wen-sing 2004[41] | Germany / Saxony-Anhalt | Controlled before-and-after study | PHC | 87 GPs in 10 groups of 7–12; control group: 90 GPs not participating in the intervention. | Evalua-tion after 2 years | Determine the impact of a large-scale programme of QCs on quality and costs of prescribing, 11 meetings of 2 hours, existing QCs promoted by the association of statutory health insurance. | A trained facilitator (GP) supported the group. | Structured feedback report, patient video, evidence-based material, interactive learning and reflective thinking about willingness to change. | High satisfaction with QCs. Prescriptions decreased in the intervention group while increasing in the control group. Aspects of quality of prescriptions improved. | |
| Andres 2004[42] | Germany /Hessen | Survey | PHC | 483 out of 612 GPs (57 QCs) answered. | Evalua-tion after 2 years | Evaluate participants' experiences of existing QCs taking part in a large project. | 7–10 GPs in each QC, facilitator guiding through the process, support by academic staff members. | Personal prescription data with the opportunity to compare with colleagues. | Positive effects on medical practice and increase in knowledge. | |

**Supplemental material 7 Study characteristics**

| Author year | Country | Study design | Set-ting | Participants, professional background | Study duration | Objective and intervention setting | Facilitation and group dynamics | Didactic and QI technique | Outcome oriented data | Common characteristics of the cluster (kinship) |
|---|---|---|---|---|---|---|---|---|---|---|
| Fessler 2006[43] | Germany (Rhine Main) | Controlled before-and-after study | PHC | 90 GPs participating in QCs were compared to non-participants in another area | Evalua-tion after 2 or 3 years | Improve prescription patterns concerning statins, antidiabetics, other drugs for cardiovascular diseases. Intervention in existing QCs. | Facilitated group work every 4–6 weeks. | QC process according to German standards; discussion of any results not in line with guidelines. | Guideline adherence increased. | Papers about establishing QCs in Germany using data on everyday practice to improve prescription patterns. |
| Papen-dick 2006[44] | Germany (Rhine Main) | Controlled before-and-after study | PHC | 59 GPs participating in QCs compared to 52 non-participants | Evalua-tion after 12 months | Examine the development of drug costs among GPs participating in existing QCs. | Facilitated group work every 4–6 weeks. | QC process according to German standards; discussion of any results not in line with guidelines. | The cost of medical drugs and the increase in expenditure were lower compared to the control group. | |
| Wen-sing 2009[45] | Hesse, Lower Saxony, Saxony-Anhalt | 3 controlled before-and-after studies with baseline in 2001 and follow-up in 2003 | PHC | 1090 GPs in the inter-vention group and 2090 in the control group. | Baseline data 3 months; evaluation using another 3 months' data after 24 months. | Determine the effectiveness of the QC process on prescribing patterns in existing and new QC groups. Data were gathered on different groups of drugs. One QC meeting a month. | 8–14 physicians in a group, trained facilitator (GP) | Repeated feedback on prescribing patterns, evidence-based information, reasons for variations were discussed, case-based discussions, objectives for improvement were formulated and specific plans made. | Attendance rate 71–79%, high satisfaction >80%. Reduction of mean prescription costs per patient, increased prescription of recommended drugs compared to the control group. | |

**Supplemental material 7 Study characteristics**

| Author year | Country | Study design | Set-ting | Participants, professional background | Study duration | Objective and intervention setting | Facilitation and group dynamics | Didactic and QI technique | Outcome oriented data | Common characteristics of the cluster (kinship) |
|---|---|---|---|---|---|---|---|---|---|---|
| Andres 2010[46] | Hesse, Saxony-Anhalt, Westfalen-Lippe, Schleswig-Holstein | Interrupted time series 1995–2007 | PHC | 1242 QCs documented 27,255 meetings. Evaluation of QCs only if they meet at regular intervals and have done so for at least one year. | 12 years | Assess the quality of the structure, processes and results of existing QCs promoted by the association of statutory health insurance. | Facilitators questioned the groups and tried to detail an agreement on best practice. | A group of GPs met at regular intervals to consider their standard practice. Their work was based on personal experience, own data and was target-oriented to promote quality in their own practice. | 8 and 12 meetings per year, group atmosphere was generally very good; the proposed method was actually used in the groups; consensus was often achieved. | |
| Beyer 1999[47] | Saxony-Anhalt, Bremen | Cross-sectional survey | PHC | 2412 out of 4270 answered | Not applicable | Analysis of demands and expectations on supporting institutions | Not applicable | Not applicable | GPs reported good emotional support from colleagues, improved professional self-confidence, but also fear of control and excessive demands. | Paper about evaluation of reasons for and against participation in QCs. |
| Aubke 2003[48] | West-phalia-Lippe | Cross-sectional survey 1995–2001 | PHC | 520 QCs with 7350 participants: 3260 meetings were evaluated | 5 years | Assessment of QI cycle in existing QCs using a checklist. 15 GPs in each group, meeting time 120 minutes on average | Not applicable | QCs work both continuous and topic-centred, based on documentation from own practice with the aim of promoting their quality of care. | 29.6% of all QCs had implemented the PDCA cycle, 54.9% had partially implemented the characteristics. | Paper on QCs about evaluation of adherence to the PDCA cycle. |

**Supplemental material 7 Study characteristics**

| Author year | Country | Study design | Set-ting | Participants, professional background | Study duration | Objective and intervention setting | Facilitation and group dynamics | Didactic and QI technique | Outcome oriented data | Common characteristics of the cluster (kinship) |
|---|---|---|---|---|---|---|---|---|---|---|
| Beyer 2003[49] | Germany and European countries | Cross-sectional survey among EQuiP delegates | Euro-pean PHC | Reports of EQuiP delegates from 26 countries | Cross-sectional | Provide an overview of QC activities across Europe. | Facilitator is usually a GP. | A consistent group of 8 to 15 health-care professionals meet at regular intervals to consider and reflect on their standard practice. | High activity of QCs (i.e. > 10% of all GPs are involved) in 9 European countries. | Paper about the spread of QCs across Europe (Update Rohrbasser 2019). |
| Mols 2005[50] | Germany (Black Forest region) | Controlled before-and-after study | PHC | 36 GPs in QCs treated 75 patients, 25 GPs in the control group treated 51 patients | Baseline after 6 months, evaluation after 18 months. | Study the effect of existing QCs on secondary prevention of stroke. | Facilitated group work every 6 to 8 weeks. | QC process according to German standards. | QCs did not have an additional effect on secondary prevention after stroke compared to the control group. | Paper on QCs about testing guideline adherence. |
| Schnei-der 2007[51] | Germany | Ran-domised controlled trial | PHC | 12 QCs involving 96 GPs; out of 256 partici-pants, 185 responded to the follow-up. | Evalua-tion after 1 year | Evaluate the efficacy of QCs for asthma care working with individual feedback with and without benchmarking. | Trained facilitators supported the groups in the process. | Collective discussion of evidence-based pharmacotherapy and management of patients on the basis of prescribing data. | Both groups improved their guideline adherence. | Testing the question whether benchmarking in QCs improves guideline adherence - or not. |
| Vollmar 2007[52] | Germany (North-Rhine West-phalia) | Protocol of a randomised controlled trial | PHC | 174 GPs in approx. 20 QCs | Evalua-tion after 3 meetings (6 months) | Improve GPs knowledge and skills about people with dementia. | QCs are facilitated by a trainer rather than by a facilitator. | Study concept A: e-learning followed by case discussions in QCs. Study concept B: oral presentation of evidence-based information followed by a discussion led by a presenter. | Possible change of behaviour, use and acceptance of new learning tools. | Papers about evaluation of e-learning methods in QCs. |

**Supplemental material 7 Study characteristics**

| Author year | Country | Study design | Set-ting | Participants, professional background | Study duration | Objective and intervention setting | Facilitation and group dynamics | Didactic and QI technique | Outcome oriented data | Common characteristics of the cluster (kinship) |
|---|---|---|---|---|---|---|---|---|---|---|
| Vollmar 2009[53] | Germany (North-Rhine West-phalia) | Cross-sectional survey | PHC | 264 out of 449 GPs answered the questionnaire | Cross-sectional | Gain understanding of German GPs' preferences for different forms of educational methods, such as e-learning. | Not applicable | Not applicable | Approx. 70% wanted to discuss everyday practice with colleagues. Meeting experts and e-learning were not favoured. | . |
| Vollmar 2010[54] | Germany (North-Rhine West-phalia) | Ran-domised controlled trial | Ger-man PHC | 166 GPs in 26 QCs | 1 year after study start | Compare knowledge acquisition about dementia management between blended learning and QC methods alone. | QCs are facilitated by a trainer rather than by a facilitator | Study concept A: e-learning followed by case discussions in QCs. Study concept B: oral presentation of evidence-based information and its discussions in a QC. | Groups A and B improved their knowledge. A blended learning approach was not superior to the QC approach. | |
| Siebolds 2012[55] | Germany | Survey | PHC | 83 facilitators received survey | Cross-sectional | Evaluation of training and support for facilitators by tutors. | To support facilitators, the KBV (National Association of Statutory Health Insurance) developed structured didactic handouts for the QC work. | Guidelines of the National Association of Statutory Health Insurance for Quality Assurance Procedures. | High level of satisfaction with didactic handouts (manuals) and training opportunities. | Paper about the quality of training and support for facilitators. |

**Supplemental material 7 Study characteristics**

| Author year | Country | Study design | Set-ting | Participants, professional background | Study duration | Objective and intervention setting | Facilitation and group dynamics | Didactic and QI technique | Outcome oriented data | Common characteristics of the cluster (kinship) |
|---|---|---|---|---|---|---|---|---|---|---|
| **Swiss QCs** | | | | | | | | | | |
| Bugnon 2004[56] | Switzer-land | Controlled before-and-after study | PHC | 6–10 GPs in 1 QC. | Develop-ment over 3 years | Improve prescription patterns and reduce costs for drug prescriptions. | A pharmacist facilitated the group through the process of academic detailing. The group engaged in local networking. Group cohesion increased with time. | Evidence-based information, feedback on prescriptions including information about possible substitutions. Consensus discussions and agreement on best choices. | Improvement of prescription patterns (antibiotics, antidiabetic and antihypertensive drugs, NSAIDs); reduction of costs compared to control groups. | Papers about pharmacist-led QCs in Switzerland. |
| Niquille 2010[57] | Switzer-land | Controlled before-and-after study | PHC | 24 GPs in 6 QCs | Develop-ment over 9 years | Improve prescription patterns and to reduce costs for drug prescriptions. | A pharmacist facilitated groups of 3–6 GPs through the process of academic detailing. Group cohesion increased with time. | As in Bugnon 2004 | 42% decrease in drug costs, improved adherence to prescription guidelines compared to control group. | |

**Supplemental material 7 Study characteristics**

| Author year | Country | Study design | Set-ting | Participants, professional background | Study duration | Objective and intervention setting | Facilitation and group dynamics | Didactic and QI technique | Outcome oriented data | Common characteristics of the cluster (kinship) |
|---|---|---|---|---|---|---|---|---|---|---|
| **Drug Education Project** | | | | | | | | | | |
| Lund-borg 1999[58] | Sweden | Ran-domised controlled trial | PHC | 18 groups (104 GPs) compared with 18 groups (100 GPs), 3–10 GPs in each group | 6 months | Improve the treatment of asthma and urinary tract infections. The two study groups served as controls for each other. | Pharmacists facilitated the GP groups, two meetings, each meeting 1.5 hours. | Information on their judgements of written simulated cases. Discussion of actual decisions taken on the simulated cases. Discussion of personal experience of difficult clinical cases and underlying reasons for prescriptions. | Guideline adherence increased for patients with urinary tract infections and patients with asthma. | QC study on improving drug prescriptions in Sweden, Norway, The Netherlands and Slovakia: Drug Education Project. |
| Lund-borg 1999[59] | Sweden | GPs' evaluation of the trial: survey | Swe-dish PHC | 82 out of 104 GPs and 83 out 100 GPs responded. | 6 months | Capture GPs' experiences of the trial through a questionnaire. | As above in Lundborg 1999 | As above in Lundborg 1999 | 87% of participating GPs wanted to take part in similar CME activities for other conditions. | |
| Lager-lov 2000[60] | Norway | Ran-domised controlled trial | Norwe gian PHC | 32 groups (199 GPs), 4–8 GPs in each group | 6 months | As above in Lundborg 1999 | As above in Lundborg 1999 | As above in Lundborg 1999. | Guideline adherence increased. | |

**Supplemental material 7 Study characteristics**

| Author year | Country | Study design | Set-ting | Participants, professional background | Study duration | Objective and intervention setting | Facilitation and group dynamics | Didactic and QI technique | Outcome oriented data | Common characteristics of the cluster (kinship) |
|---|---|---|---|---|---|---|---|---|---|---|
| Veninga 1999[61] | Sweden, Slovakia, The Nether-lands | Evaluation of a randomised controlled trial | Swe-dish, Norwegian, Dutch and Slova-kian PHC | The Netherlands: 24 groups, 181 GPs; Sweden: 36 groups, 204 GPs; Norway: 32 groups, 199 GPs; Slovakia: 20 groups, 81 GPs. | 6 months | Explore whether a specific educational approach for implementation of guidelines has a similar effect when used in different health care settings. | As above in Lundborg 1999 (Slovakia only one meeting). | As above in Lundborg 1999 | Attitudes changed and prescription patterns improved. | QC study on improving drug prescriptions in Sweden, Norway, The Netherlands and Slovakia: Drug Education Project (DEP). |
| Veninga 2000[62] | The Nether-lands | Ran-domised controlled trial | PHC | 24 groups (181 GPs) | 6 months | As above in Lundborg 1999 | As above in Lundborg 1999 | As above in Lundborg 1999 | Guideline adherence increased. | |
| **European single studies** | | | | | | | | | | |
| Eliasson 1999[63] | Sweden | Literature review, survey and authors' reflections | PHC | 5–10 GPs in each of approx. 230 groups | Meeting once to twice a month | Give an overview of CME group work in Sweden and describe its strengths and weaknesses. | Facilitated group discussions. Reflection on emotional responses was part of the group process. | Prearranged modules with short introductions and facts on a topic. Discussions based on experiences. | 80% of the group members assessed the pedagogical value of the group sessions as more valuable than direct instruction. | Paper on Swedish QCs |

**Supplemental material 7 Study characteristics**

| Author year | Country | Study design | Set-ting | Participants, professional background | Study duration | Objective and intervention setting | Facilitation and group dynamics | Didactic and QI technique | Outcome oriented data | Common characteristics of the cluster (kinship) |
|---|---|---|---|---|---|---|---|---|---|---|
| Watkins 2004[64] | England | Qualitative study: focus group | PHC | 6 different facilitators with different backgrounds. A total of 19 GPs in four practices and one practice manager took part. 11 GPs were interviewed. | 7 monthly sessions taking place at midday | Reflect on inappropriate and costly prescribing. Investigate feasibility of educational sessions for GPs: acceptability among GPs and possible barriers. | 'Reflective practice' as a potential solution to high-cost prescribing. GPs felt that participation was to appease their prescribing adviser. No or little sense of ownership. Information overload was a problem. | Video-tape of a scenario, followed by brainstorming, and personal responses in the group. 'Best buy' response was selected. Identification of barriers to implementation and discussion of means to overcome barriers. | Low response for participation (4 out of 61 practices). There was friction between clinical autonomy and the experience of a top-down intervention. | Paper on English QCs (reflective groups) |
| Tonies 2006[65] | Austria | Survey | PHC | In 2001, 29 GPs out of 169 (17%) responded; in 2002, 46 out of 272 (27%) responded. | Evalua-tion after 4 years of offering QCs | Improve care of patients with drug replacement therapy using synthetic opioids in PHC. | A GP facilitated the group and had the support of an experienced local opinion leader. | Local opinion leaders introduced topics. Stimulation of discussions to increase self-awareness and frustration tolerance. | High level of satisfaction with the teaching. Communication skills improved. Topic-specific knowledge increased. | Topic-specific QC activities in Austria. |
| Riou 2007[66] | France | Controlled before-and-after study | PHC | Number of groups is not mentioned, 7–11 GPs per group, 24 participating GPs, 3–6 local pharmacists in each area. | 12 months (Dec 2001 to Dec 2002) | Improve prescription patterns in three semi-rural areas of Brittany, France. Financial incentive. | 4 plenary meetings with consultants lecturing on pre-specified topics. QCs every 6th week using personalised feedback. | Expert input during plenary sessions, voluntary feedback, peer review and specific recommendations for changes during QCs. | Increase in generic prescription rates and decreased prescription of drugs with no evidence-based efficacy. | French QC study on improving drug prescription patterns. |

**Supplemental material 7 Study characteristics**

| Author year | Country | Study design | Set-ting | Participants, professional background | Study duration | Objective and intervention setting | Facilitation and group dynamics | Didactic and QI technique | Outcome oriented data | Common characteristics of the cluster (kinship) |
|---|---|---|---|---|---|---|---|---|---|---|
| van Driel 2007[67] | Belgium | Cluster randomised controlled trial | PHC | 9 QCs (122 GPs) in the intervention and 9 QCs (134 GPs) in the control group | November 2004 to March 2005 | Improve antibiotic prescribing in patients with rhinosinusitis. Existing QCs. | The group meetings were scheduled as regular QC sessions without the presence of an external expert. | Dissemination of the guidelines by e-mail; facilitators received educational material concerning antibiotics. | A single intervention in QCs did not have a significant effect on prescription patterns. | Belgian QC study on improving drug prescription. |
| Spiegel 2012[68] | Austria | Qualitative evaluation | PHC | 445 out of 821 GPs took part in the groups, 8–10 participants in each group | 2 years: 2004 and 2005 | Explore GPs' perception of QCs concerning prescribing habits. Qualitative analysis was used to evaluate QC protocols. | Facilitators' duties were to schedule dates for QCs, give introductory talks on intended topics and facilitate the group process. | Use of educational material on various issues of pharmacotherapy; costs were addressed; provision of personal feedback on prescription habits. | Prescription of generic drugs increased. | Austrian QC study on improving drug prescription. |
| **OTHER AREAS** | | | | | | | | | | |
| de Villiers 2003[69] | South Africa | Qualitative evaluation using Nominal Group Technique followed by survey | PHC | 64 GPs answered (response rate 38%), 51 out of 101 responding GPs had participated in QC, 8 out of 12 facilitators responded | Evalua-tion of 9 months CME/ CPD activity | A nominal group technique was used to compose two questionnaires (for participants and facilitators) | Facilitated small-group activities | Activities built on previous experience, involved the learners, focussed on relevant problems; solutions were applicable in practice; the process followed a cycle of action-reflection and GPs acquired technical skills. | 91% of the respondents indicated improved knowledge, 73% indicated improvements in their patient care and 61% improved clinical skills | South African QCs |

**Supplemental material 7    Study characteristics**

| Author year | Country | Study design | Set-ting | Participants, professional background | Study duration | Objective and intervention setting | Facilitation and group dynamics | Didactic and QI technique | Outcome oriented data | Common characteristics of the cluster (kinship) |
|---|---|---|---|---|---|---|---|---|---|---|
| Rich-ards 2003[70] | New Zealand | Pilot study: retro-spective, controlled before-and-after study | PHC | 52 GPs in small education groups: approx. 10 GPs in each group | After 1 and 2 years | Determine whether a QC-programme designed to promote rational GP prescribing succeeds in changing practice when added to audit and feed-back, academic detailing. | Meetings were monthly and group composition remained the same over time. | Control group: audit and feedback on prescription habits, academic detailing and educational bulletins. Intervention group: peer-led groups, monthly meetings. | Positive effect of the education strategy in groups compared to the combination of audit and feedback and academic detailing. | QCs on improving drug prescriptions in New Zealand. |
| Parker 2007[71] | USA (Hawaii) | Ran-domised controlled trial | PHC | 4 health-care facilities of similar size participated and were randomly assigned the local or the central QI approach | Duration about 2.5 years | Compare the participatory local approach with the central expert approach to QI in depression care. | Researchers allowed teams to design their own programmes. Local QI groups had a facilitator. | The QI teams followed guidance regarding team composition and process. The central expert approach used centrally organised teams of experts. | A hybrid model (central expertise and local participation) may be the most effective approach to maintain a high level of motivation. | QCs on Hawaii compared to centrally steered options. |
| Som-mers 2007[72] | USA (Califor-nia) | Survey and attendance rate | PHC | Researchers invited 30 sites, 11 (103 GPs) out of 14 sites who started continued with their meetings | 5 years | Introduce small-group meetings as means of managing clinical uncertainty. | A group member or an invited, external member facilitated discussions, searched for and appraised evidence and coordinated meeting logistics. | Reflection on and appraisal of one's own delivery of clinical care. Case-based discussion and reflection. | Most common themes: being with colleagues, the role of time in GP practice. Other common themes: acknowledging uncertainty, receiving validation. | Practice-Based Learning and Improvement in in California. |

**Supplemental material 7 Study characteristics**

| Author year | Country | Study design | Set-ting | Participants, professional background | Study duration | Objective and intervention setting | Facilitation and group dynamics | Didactic and QI technique | Outcome oriented data | Common characteristics of the cluster (kinship) |
|---|---|---|---|---|---|---|---|---|---|---|
| Murrihy 2009[73] | Australia | Before-and-after study | PHC | 6 groups of GPs (32 GPs) | 6 months | Improve GPs' skills and actual use of cognitive behaviour therapy. 8 two-hour sessions. | Expert-led small-group interactive learning, and ongoing discussion of patients. | Development of mentor-type relationships, the use of interactive learning and skills-based training, discussion of ongoing patients. | GPs' knowledge, skills in and actual use of cognitive behaviour therapy increased. | QCs in Australia. |
| **UPDATE December 2020** | | | | | | | | | | |
| Fisher 2013[74] | North East Ohio, USA | Before-and-after study and survey (qualitative data) | PHC | 78 participants in 20 practices/ groups; some groups were inter-professional | 1 year | The American Board of Medical Specialties' Performance and Practice initiated the project to support GPs in working in groups to improve practice. | A coach facilitated the process, led discussions, helped the team to recognise their skills, to identify the next steps and to address problems arising. | Physicians discussed their priorities for improvement, narrowed the topic, reflected on results of patient surveys and shared their view of 'best practice' using personal examples. | Introduction of QI tools into groups succeeded. Participants felt that the group activity encouraged collaboration with colleagues. | Practice-Based Learning and Improvement in the USA. |
| Francois 2013[75] | Isère, France | Survey | PHC | 16 groups, 132 GPs | Not applicable | Review the implementation of QCs by mapping the groups, describe the perspective of participants and study how these groups work. | Facilitators helped the groups to share experiences and to discuss difficult cases and medical errors. | Case discussions, audit charts to analyse prescription habits, interactive learning, reflective thinking and consensus-finding, local opinion leaders. | 6–10 GPs in each group, meetings lasted between 1 and 2.5 hrs, 6–10 meetings per year, participants had a high level of satisfaction. | Description of QC development in Isère, France. |
| Wilcock 2013[76] | England | Cluster randomised controlled trial | PHC | 11 practices using workshops, 12 practices usual care | 12 months | Test of a tailored educational intervention on the clinical management of | Facilitated small-group workshops with practice teams. | Adult learning approach to solving real-world problems, tailoring the learning need, using | The intervention did not alter the clinical management of patients with | QC-like intervention in England testing guideline adherence. |

**Supplemental material 7 Study characteristics**

| Author year | Country | Study design | Set-ting | Participants, professional background | Study duration | Objective and intervention setting | Facilitation and group dynamics | Didactic and QI technique | Outcome oriented data | Common characteristics of the cluster (kinship) |
|---|---|---|---|---|---|---|---|---|---|---|
| | | | | (NICE guidelines) | | people with dementia. | | workshops at the work place. | dementia. | |
| Andres 2015[77] | Germany | Focus group | PHC | 12 health-care professionals | Not applicable | Evaluation of 20 years' QC work | Maintaining autonomy, self-determination of topics and the process in QCs ensure the practical relevance of topics and emotional engagement of participants. | Case-based learning among peers in a facilitated group process is key in the QC process. | Measures to support QC-work: evidence-based information and trustworthy prescription patterns. | 20 years' experience of QCs in Germany |
| Dowling 2015[78] | Ireland | Survey | PHC | 96% of GPs participating in CME groups responded (1366), 146 groups | Not applicable | Investigate whether taking part in CME groups improves GPs' clinical knowledge. | A local, small-group setting provides live peer-group interaction, peer support and reflection on practice. | Face-to-face activities, multiple exposure, the use of multi-media and multiple education techniques. | 97% stated that they want to improve their clinical practice, 86.3% agreed that taking part in CME groups is key for this. | QCs in Ireland |
| Ver-bakel 2015[79] | The Nether-lands | A three-group cluster randomised controlled trial | PHC | 10 groups in each intervention group | 4 months | Assess the effect of two interventions on patient safety culture: a survey compared to adding a QC-like intervention compared to usual care. | Team-based reflection on personal practice data and team-based development of action plan. | Didactics were added to the experiential learning principles of Kolb, for example, concrete experience, reflection, conceptualisation, and experimentation. | Increased reporting of critical incidents | Dutch QC study on improving patient safety culture. |

**Supplemental material 7 Study characteristics**

| Author year | Country | Study design | Set-ting | Participants, professional background | Study duration | Objective and intervention setting | Facilitation and group dynamics | Didactic and QI technique | Outcome oriented data | Common characteristics of the cluster (kinship) |
|---|---|---|---|---|---|---|---|---|---|---|
| Mahl-knecht 2016[80] | Austria (Salzburg) and Italy (Tirol) | Before-and after-study | PHC | 20 GPs in regional QC groups (number of groups not mentioned) | 3 years | Assess whether quality can be improved by self-auditing, benchmarking and QCs. | Facilitated, regular group meetings | Critical self-reflection, audits and feedback, benchmarking. | The mean quality score increased significantly. | Austrian–Italian study using benchmarking in QCs. |
| Vervloet 2016[81] | The Nether-lands | Controlled before-and-after study | PHC | 4 groups (39 GPs) in the intervention and 4 groups (38 GPs) in the control group | 1 year | Evaluate the effect of a multifaceted, peer-group-based intervention aiming to reduce respiratory tract related antibiotic prescriptions. | A series of regular meetings between GPs and pharmacists in the same catchment area. | Communication skills training, including communication about delayed prescribing, quarterly feedback figures for GPs. | Guideline adherence increased. | Dutch QC study on improving drug prescription involving pharmacists. |
| Jäger 2013[82] | Germany | Protocol of a cluster randomised controlled trial | PHC | 10 QCs (40 GPs) | 6 months | To implement structured medication counselling, use of medication lists and medication reviews to avoid potentially inappropriate medication. | QC meetings every three months. | Development of individual concepts of change and their presentation at QC meetings. Posters and flyers for patients. Written feedback on individual practice patterns. | The degree of implementation of the three recommendations measured at patient level. | German QC study on improving drug prescription. |
| Jäger 2015[83] | Germany | Description of intervention | PHC | 12 GPs and 8 medical practice assistants from 8 practices participated in the workshop. | 6 months | Describe the content and delivery of the tailored intervention. | No further mention of QCs in the paper. | Workshops about structured medication counselling, use of medication lists and medication reviews to avoid potentially inappropriate medication. | The workshop seemed to improve participants' knowledge of medication management. | German QC study on improving drug prescription. |

**Supplemental material 7 Study characteristics**

| Author year | Country | Study design | Set-ting | Participants, professional background | Study duration | Objective and intervention setting | Facilitation and group dynamics | Didactic and QI technique | Outcome oriented data | Common characteristics of the cluster (kinship) |
|---|---|---|---|---|---|---|---|---|---|---|
| Jäger 2017[84] | Germany | A cluster randomised controlled trial | PHC | Intervention group: 10 GPs in 5 different QCs; control group: 11 GPs in 6 different QCs, | 6 months | As above in Jaeger 2013 | Not mentioned | Training for GPs and medical practice assistants, educational material for patients, individually developed action plans, written feedback on prescription patterns. | Little or no effect of the tailored programme on the combined primary outcome could be substantiated. Lack of statistical power to detect any effect. | |
| Jäger 2017[85] | Germany | Interviews | PHC | Analysis of 12 interviews, 21 question-naires, 120 documenta-tion forms. | Evalua-tion of 6 months' study | To evaluate the study Jaeger 2017 using various data sources. | Facilitation or group dynamics were not described as QCs were not used as planned. | Workshop-like atmosphere of one meeting. | Patients were not able to use the tablets provided. Participants suggested integrating the training into QCs. | |
| Ter Brugge 2017[86] | The Nether-lands | Mixed-methods design: question-naire about types of group meetings followed by interviews | PHC | 78 out of 128 GP supervisors filled out the questionnaire; 18 GP supervisors were interviewed | Not applicable | Examine different types of group meeting and explore the use of clinical research evidence. | Little discussion on clinical applicability of evidence. | Guidelines, local opinion leaders who lecture, consensus discussion. | QCs are the type of group meeting that occur most often in PHC. They seem to be more goal-oriented than learning-oriented. The agenda was heavily influenced by health insurance companies. | Dutch QC study on improving drug prescription involving pharmacists. |

**Supplemental material 7 Study characteristics**

| Author year | Country | Study design | Set-ting | Participants, professional background | Study duration | Objective and intervention setting | Facilitation and group dynamics | Didactic and QI technique | Outcome oriented data | Common characteristics of the cluster (kinship) |
|---|---|---|---|---|---|---|---|---|---|---|
| Trietsch 2017[87] | The Nether-lands | A cluster randomised controlled trial | PHC | 21 QCs (197 GPs) | 3 years | Test the effect of audit and feedback with peer review on GP' prescribing and test-ordering performance. | Facilitation by local opinion leaders (laboratory specialist or local pharmacist) who were trained in a three-hour meeting. The groups met twice for each topic. | Facilitators had written and digital evidence-based materials, individual feedback reports | The increase in total tests ordered was 3% in the intervention and 15% in the control group. The increase in prescriptions was 20% in the intervention and 66% in the control group. | Dutch QC study on improving test ordering and drug prescription. |
| Andres 2018[88] | Germany | Controlled before and after study | PHC | 48 GPs | 12 months | Test the effect of audit and feedback with peer review on quality indicators for coronary heart disease (CHD) | Classic German QC without further description | Individually presented 11 quality indicators for patients with CHD; feedback reports for each doctor's practice at two QC meetings | For three of these indicators the increase rates were higher than those in the Bavarian control group | German study of use of quality indicators in QCs |
| Binienda 2018[89] | USA (Ohio) | Survey | PHC | 126 GPs | Not applicable | To explore the research efforts of Practice Based Research Networks (PBRN) | Not applicable | Not applicable | PBRNs currently thrive on conducting research predominantly in quality improvement and practice transformation | QI in US |

**Supplemental material 7 Study characteristics**

| Author year | Country | Study design | Set-ting | Participants, professional background | Study duration | Objective and intervention setting | Facilitation and group dynamics | Didactic and QI technique | Outcome oriented data | Common characteristics of the cluster (kinship) |
|---|---|---|---|---|---|---|---|---|---|---|
| Kral 2018[90] | Czech Republic | Case study | PHC | GPs, not stated how many | 6 months | Use of quality circles as a support tool in the taking over of practices by young general practitioners. | 1st meeting, identification of problems; 2nd meeting, discussion of specific issues of starting to practice; 3rd meeting, analysis of the suggested measures and implementation; 4th meeting, evaluation. | Facilitated discussions | QC work offers a good platform for young GPs in starting their own practice. | QC pilot in the Czech Republic |
| Park 2018[91] | Scotland | Focus groups | PHC | GPs/Practice Nurses/Pharm acists | Not applicable | To determine how groups recruit new members and discern what are the important attributes of the new members. | Not applicable | Not applicable | 4 themes: group formation and purpose; group culture; experience of group members; professional socialisation. | Recruitment to PBSG in Scotland |
| Pedersen 2018[92] | Norway | Case series | PHC | 53 health care professionals PHC | 12 months | to investigate what is discussed when QCs work to complete an action form as part of an audit and feedback cycle. | Insight into their own and their colleagues' practices. | Discussion of results of the audit; identification of gaps between recommendations and local practice; choice of areas for improvement; addressing local barriers and enablers; evaluation. | Acting on audit and feedback provided an opportunity to discuss practice. | QC I Norway |

**Supplemental material 7 Study characteristics**

| Author year | Country | Study design | Set-ting | Participants, professional background | Study duration | Objective and intervention setting | Facilitation and group dynamics | Didactic and QI technique | Outcome oriented data | Common characteristics of the cluster (kinship) |
|---|---|---|---|---|---|---|---|---|---|---|
| Rogn-stad 2018[93] | Norway | Cluster-randomised controlled study | PHC | 80 CME groups; 7–8 GPs in each group located in the southern part of Norway | 6 months: meetings once a month; the study covered 3 meetings. | To undertake a multifaceted, educational intervention to improve GPs' prescribing practice for patients aged ⩾ 70. | See Rognstad 2013 | See Rognstad 2013 | Reduction of Potentially inappropriate prescriptions. | Norwegian QC studies on improving drug prescriptions |
| Rogn-stad 2018[94] | Norway | Cluster-randomised controlled study | PHC | 80 CME groups; 7–8 GPs in each group located in the southern part of Norway | 6 months: meetings once a month; the study covered 3 meetings. | To explore the characteristics of the GPs responding to QC intervention. | See Rognstad 2013 | See Rognstad 2013 | GPs with the lowest adherence to recommended practice at baseline improved their practice most. | Norwegian QC studies on improving drug prescriptions |
| Will-man 2018[95] | Scotland | Survey | PHC | Not known | Not applicable | To assess the educational impact of PBSGL. | Not applicable | Not applicable | PBSGL is an essential pillar for supporting all doctors in Defence Primary Healthcare. | Scottish PBSGL |

**Supplemental material 7 Study characteristics**

| Author year | Country | Study design | Set-ting | Participants, professional background | Study duration | Objective and intervention setting | Facilitation and group dynamics | Didactic and QI technique | Outcome oriented data | Common characteristics of the cluster (kinship) |
|---|---|---|---|---|---|---|---|---|---|---|
| Cunning ham 2019[96] | Scotland | Evaluation | PHC | Not applicable | Overview of 17 years | To increase clinical knowledge and to implement it. | Facilitated discussion case presentations; study of current evidence base; proposal of changes to practice. | Members are encouraged to make a commitment to change, to log these changes in a shared document, and to review changes with their colleagues. | 3,400 members drawn from GPs, GP nurses, pharmacists and other professions. | Scottish PBSGL overview |
| Dowling 2019[97] | Ireland | Survey | PHC | 1686 GPs answering the questionnaire | Not applicable | To examine whether local, accessible ongoing CME-SGL for rural GPs meets their educational needs. | Not applicable | Not applicable | 87% reported that their educational needs were fully or mostly met. | Irish CME groups |
| Martin 2019[98] | Switzer-land | Before and after study | PHC | 9 GPs | 2 years | Assess status of colorectal carcinoma screening and use of shared decision when choosing screening method. | Facilitated small group work according to Swiss standards. | data-driven Plan-Do-Study-Act cycles to implement changes in practice. | Through data-driven PDSA cycles and organisational changes, GPs implemented SDM tools in their daily routine. | Swiss QC on screening of colorectal carcinoma |

**Supplemental material 7 Study characteristics**

| Author year | Country | Study design | Set-ting | Participants, professional background | Study duration | Objective and intervention setting | Facilitation and group dynamics | Didactic and QI technique | Outcome oriented data | Common characteristics of the cluster (kinship) |
|---|---|---|---|---|---|---|---|---|---|---|
| Sieben-hofer 2019[99] | Germany | Cluster randomised controlled study | PHC | 52 general practices | 24 months | To examine whether case management reduces thromboembolic events and major bleeding events. | Training for healthcare assistants; information and quality circles for GPs; 24 months of case management. | Quality circles to discuss practical problems; case discussions. | The intervention appears to have positively influenced several process parameters under 'real-world conditions'. | German QCs on antithrombotic treatment |
| Armson 2020[100] | Canada | Mixed methods | PHC | 139 GPs | Not apppli-cable | To assessed feasibility and effectiveness of practice-based small-group learning in academic half days; question-naire and interviews. | Participants were divided into groups of 14-16 members to discuss 12 different module topics. | Presentation of clinical cases presented in educational modules and reflection on own clinical experiences; trained peer facilitator. | Feasible approach for half day learning sessions. | Canadian PBSGL |
| Dowling 2020 [101] | Ireland | Before and after study using mixed methods | PHC | 4 CME groups including 43 GPs | 6 months | To identify whether CME-small group learning increases knowledge and changes behaviour; questionnaires, prescribing audits and qualitative focus groups. | A two-hour teaching module on deprescribing in older patients was devised and implemented. | Needs assessment; four case studies and own examples; facilitated discussion. | Learning outcomes seemed achieved; 79.9% of cases were de-prescribed; sharing experiences helped them change practice | Irish CME groups |

**Supplemental material 7 Study characteristics**

| Author year | Country | Study design | Set-ting | Participants, professional background | Study duration | Objective and intervention setting | Facilitation and group dynamics | Didactic and QI technique | Outcome oriented data | Common characteristics of the cluster (kinship) |
|---|---|---|---|---|---|---|---|---|---|---|
| Mahl-knecht 2020[102] | Austria and Italy | Before and after study | PHC | 56 GPs | 2 years | To assess the changes in quality of life (QoL) and patient satisfaction of chronically ill patients in Tyrol and South Tyrol. | Not described | Intervention consisted of self-audit, benchmarking and QCs | The impact of the intervention was not significant within the intermediate time periods analysed in the study. | QCs in Tyrol (Austria and Italy) |
| Mercer 2020[103] | Scotland | Survey | PHC | 4371 GPs | Not applicable | To determine GPs' views on QCs. | QC participants were asked to what extent QCs were: 1) well organised; 2) friendly; 3) well facilitated; and 4) productive | Not applicable | 2456 responses were received from 4371 GPs (56.4%). QCs are in need of more support to improve quality of care | Scottish PBSGL |
| Plüss-Suard 2020[104] | Switzer-land | Before and after study | PHC | GPs, nurses and pharmacists | 6 Years | To describe antibacterial use in long-term care facilities and to investigate the determinants of use. | Improving the enforcement of clinical guidelines within long term care facilities prescribing practices. | Benchmarking, analysis of attitudes towards guidelines, building consensus and evaluation of results. | Antibacterial use decreased from 45.6 to 35.5 DDD per 1000 beds per day. | Swiss QC on drug prescription |

**Supplemental material 7 Study characteristics**

| Author year | Country | Study design | Set-ting | Participants, professional background | Study duration | Objective and intervention setting | Facilitation and group dynamics | Didactic and QI technique | Outcome oriented data | Common characteristics of the cluster (kinship) |
|---|---|---|---|---|---|---|---|---|---|---|
| Kamradt 2018[105] | Germany | Study protocol: three-armed cluster randomised trial compared to standard care | PHC | 193 practices | 3 years | To examine the change of the antibiotic prescription rate within three intervention arms and the comparison between the three intervention arms | Various social mechanisms influence the spread of new attitudes and behaviours | A: e-learning, QCs, data feedback<br>B: A plus in addition, feedback tailored for practice staff<br>C: A plus computerized support and multiprofessional QC. | Established indicators of the European Surveillance of Antimicrobial Consumption Network. Process evaluation: interviews. | German QC for rational antibiotic prescribing patterns. Effectiveness study is still pending. |
| Poss-Doering 2020[106] | Germany | Evaluation: interviews and surveys | PHC | 76 GPs and 80 medical assistants | Not applicable | To describe the individual and organizational factors affecting the uptake of this multi-faceted program using surveys and interviews | Not applicable | Not applicable | Highest uptake gave feedback reports, background information, e-learning modules and disease-specific QCs. | |
| Poss-Doering 2020[107] | Germany | Evaluation: interviews | PHC | GPs, medical assistants and stakeholder representa-tives | Not applicable | To explore factors and processes attributed to the network's contribution to improving antibiotic prescribing. | Not applicable | Not applicable | Professional peer exchange, social support and reassurance contributed to behaviour change. | |

**Supplemental material 7 Study characteristics**

| Author year | Country | Study design | Set-ting | Participants, professional background | Study duration | Objective and intervention setting | Facilitation and group dynamics | Didactic and QI technique | Outcome oriented data | Common characteristics of the cluster (kinship) |
|---|---|---|---|---|---|---|---|---|---|---|
| Stewart 2020[108] | Scotland | Evaluation: interviews | PHC | GPs, secondary care doctors | Not applicable | To identify the perceptions and experiences of participants in mixed groups of general practitioners and secondary care doctors | Not applicable | Not applicable | There was desire to improve working relationships; logistics of arranging further meetings seemed challenging. | Scottish PBSGL in mixed groups (GPs and secondary care doctors) |

**Supplemental material 7 Study characteristics**

**Supplemental material 7 Study characteristics**

**Supplemental material 7 Study characteristics**

**Supplemental material 7 Study characteristics**

**Supplemental material 7 Study characteristics**

**Supplemental material 7 Study characteristics**
