## Supplemental material 8 for "How and why do Quality Circles work for General Practitioners - a realist approach"

**Supplemental material 8: literature based programme theory including illustrative quotes and complete list of supporting quotes**

| |
|---|
| CMO configuration 1: **'participants know what to expect'**<br><br>If the introductory workshop conveys the principles of QI in PHC and the workings of QCs (social persuasion) (C), this will increase future participants' motivation to join QCs (O) because they learn what to expect and may feel that they are capable of meeting expectations (increase of self-efficacy) (M).<br><br>*Surveys have revealed wide-reaching gaps in information, some of which are the cause of misunderstandings and misjudgements. In particular, the working methods and objectives of medical quality circles are apparently insufficiently known. Better general information on this subject, which contracted doctors, in particular, expect from their KV [health insurance company], is therefore urgently needed. As examples from the Netherlands and Great Britain show, active information from the target group is a basic prerequisite for quality-enhancing measures in practice [translated from German][1].*<br><br>*Introduction of the model … is important for understanding and helps participants during the start of the process. It was also vital to have a common and shared understanding of the problem among participants. It is worthwhile taking the time for an agreement on shared guidelines …[2].*<br><br>*To deal with these issues [information overload], an initial, introductory session of 'Reflective Practice' needs to be included, where GPs' experience of previous prescribing management interventions can be aired, where safe 'rules of engagement' can be agreed, and the purpose of the 'reflective practice' intervention made explicit[3].* |
| CMO configuration 2: **'need for autonomy and obligation'**<br><br>If the administration at the national level or at the level of health insurance companies entrusts GPs with QI and autonomy (puts them in control of how do it) (C), then GPs may consider participating in QCs (O) because they feel they can take on the responsibility and make a difference (M).<br><br>*In the discussion with facilitators, the QC participants expressed their desire to be self-determined and work independently. For this reason, the Federal Association of Statutory Health Insurance Physicians (KBV) and the Associations of Statutory Health Insurance Physicians (KVen) have laid down the thematic and methodological autonomy of QCs as a prerequisite in their guidelines for quality-circle work and have committed themselves to supporting them [translated from German [4].* |
| CMO configuration 3: **'sharing similar needs'**<br><br>If the administration at the organisational level of QCs provides support (i.e. in training facilitators, data gathering, provision of evidence-based information), and the administration protects time and space and offers CME points and small financial incentives to QC participants (C), then the latter will meet in groups to exchange ideas (O) because GPs prefer learning in QCs (M); support generates positive expectations among participants (M) and GPs believe that QC meetings with their peers will be useful (M).<br><br>*External staff should organise QCs as facilitators have too little time to do this [translated from German][1].*<br><br>*The most-cited reason for joining Problem Based Small Group Learning (PBSGL) ……… their preferred learning style. 'I find my preferred method of learning to be in small groups and case*-study *discussion, so this programme seems ideal for my learning needs[5].*<br><br>*To ensure attendance in the future, the educational sessions need to be protected by the use of a paid locum, in the same way as other practice development work is now being supported[3].* |

**Supplemental material 8: literature based programme theory including illustrative quotes and complete list of supporting quotes**

| |
|---|
| CMO configuration 4: **'need for relatedness'**<br><br>If a steady group of members engages in socially enjoyable contact, led by a skilled facilitator who, e.g., introduces people to each other, opens discussions, clarifies and summarizes statements (C), then group members will get to know each other and norm rules that they are willing to follow and build safe environment based on trust (O), because members want to be among and to interact with equals (M).<br><br>*We estimate it took three to four sessions for the group to be comfortable with this process. Open discussions and debates then came more freely, and the group continued to gel*[6].<br><br>*Interestingly, the stage of storming, which is characterized by interpersonal hostility and conflict, was not evident in either group*[5].<br><br>*The role of the facilitator has been recognised. They need to be competent at many tasks including opening the discussion, clarifying, summarising, questioning and devising strategies to improve group function*[7].<br><br>*GPs regard the group as a place for social support, … growth in the professional role … for protection against burnout. Although … main purpose of small-group work is exchange … of knowledge, social aspects should not be neglected because they will increase the motivation to continue with meetings ….* [8].<br><br>*The success of group learning between GPs within a practice depends to a large extent on the quality of relationships within the group. Where individuals feel that their management decisions are under threat from colleagues with whose judgements they are not comfortable, discussion may be abruptly curtailed* [3]. |
| CMO configuration 5: **'need for autonomy and control'**<br><br>If the group members choose their own topics and facilitator (C), then they will feel they own the QC (O) because their need for autonomy - a feeling of being in control of their own behaviour - is satisfied (M).<br><br>*Tutors did not consider themselves as 'experts' but as 'one of them'. Being open about their background as GPs was an agreed-upon strategy, and tutors deliberately tried to avoid being perceived as experts. The tutors experienced that their own background was important for GPs' trust and acceptance*[9].<br><br>*The facilitator is selected by the group*[10].<br><br>*It is important for a learner to be in control of his or her learning process, to be motivated and to perceive meaningfulness*[11].<br><br>*… the rise of evidence-based medical guidelines probably decreases individual providers' autonomy. Physicians have raised similar concerns about threats to the autonomy of their profession …. … It is within this context, … declining perceived autonomy for … physicians, that we compare the participatory local and central expert approaches to QI*[12]. |
| CMO configuration 6: **'size of the group affects communication'**<br><br>If group size exceeds 15 (C), then interaction among group participants decreases (O) because participants cannot keep up with all participants and follow their conversations (M).<br><br>*All GPs participating in such peer groups, on average consisting of six to eight peers, located in southern Norway*[13].<br><br>*How can QCs be supported? (Table 2) Group sizes > 15 or < 5 - are problematic and participants need support [translated from German]*[14]. |

**Supplemental material 8: literature based programme theory including illustrative quotes and complete list of supporting quotes**

| |
|---|
| *The effect of the educational asthma programme was partly modified by the group size; prescribing behaviour for [asthma] exacerbations improved more in smaller groups. The group size varied from 4 to 13…. This result is … an optimal group size of 5 to 6 group members*[15]. |
| CMO configuration 7**: 'feeling safe and not vulnerable'**<br><br>If participants trust each other (C), then they can disclose how they work and also the holes in their knowledge (O), because they feel safe rather than vulnerable (M).<br><br>*In time, group members develop confidence and security in the group, rendering the disclosure of ignorance and 'blind spots of knowledge' easier. Group members could either use the whole group or parts of it to assess their own learning needs*[8]. |
| CMO configuration 8: **'need for competence and self-actualisation'**<br><br>If the facilitator supports the participants and encourages them to tell their stories and share their experiences in a safe environment, e.g. by encouraging interactive responses, through discussions and by summarising statements (C), then participants will become involved and share their positive experiences and failures (O) because they want to improve their professional competence (M), gain professional confidence (M) and fulfil their professional potentials (M).<br><br>*Subjects, topics and cases discussed in groups come from daily work and are highly relevant to practice. The small group will meet the demands of developing generalist knowledge as well as the expert role in general practice*[8].<br><br>*Small groups will have opportunities to discuss the 'art of medicine', founded upon context, anecdote, patient stories of illness and personal experiences. Accepting emotional responses being mirrored by other group members corresponds in some respects to the process in Balint groups*[8].<br><br>*Comparison with one's peers was important, as was the support, confidence and reassurance that some gained from being part of the group*[7].<br><br>*Exchanging experiences in QCs, GPs can work out and clarify the characteristics of general practice, which improves knowledge transfer [translated from German]*[16].<br><br>*'It [the role as a facilitator] gives you licence to play devil's advocate as well and challenge people a bit more whereas, if you were always doing that as just a group member, people might think you were just doing it to annoy them*[17]. |
| CMO configuration 9: **'previous knowledge is activated'**<br><br>If participants exchange case stories and experiences while actively listening to each other in the presence of a skilled facilitator in a safe environment (C), then they will share their knowledge by telling their own relevant stories (O) because the process activates knowledge they already possess (M).<br><br>*The use of a case-based format encourages activation of previous knowledge, allowing better retrieval of knowledge in the clinical setting…, particularly when it involves participation in small peer groups that foster trust, promote discussion of evidence relevant to real cases and provide feedback on performance*[10].<br><br>*During discussions at the level of relationships [case discussion], the exchange is more intense than in the exchange of pure facts; one's own behaviour is better analysed and suggestions arose for training in one's own practice [translated from German]*[18]. |

**Supplemental material 8: literature based programme theory including illustrative quotes and complete list of supporting quotes**

| |
|---|
| *Virtually everyone participates in presenting a case, asking for advice or clarification, or describing their practice patterns*[6]. |
| CMO configuration 10: **'immediate relevance for the practice'**<br><br>If QCs use the technique of experience-based learning (C), then knowledge becomes more relevant to GPs (O), because it relates to their everyday work and is therefore of immediate use (M).<br><br>*To better support PCPs (GPs) in managing uncertainty, more meeting time should be spent on the deliberate practice of blending evidence with experience (e.g. per-case, focused analysis of guidelines/relevance) and using case follow-up insights to 'reconstruct practice' for the individual patient while appreciating implications for the clinic/office*[19].<br><br>*There also must be some motivation for learning and change: this can be ensured if the issues discussed are derived from the learner's own clinical practice*[6].<br><br>*The decision to focus on clinical problems instead of tests was a good choice, since it allowed the feedback and group work to be linked to national evidence-based guidelines. GPs appreciated this approach, because it was also closely related to their everyday work routine*[20].<br><br>*By discussing specific cases, real problems in participants' everyday practice become the topic of discussion in QCs instead of designed problems. In systematic reconstructions [of patient situations], the experiences are made conscious, so that intuitively applied – implicit – mental guidelines can be made explicit [translated from German]*[21]. |
| CMO configuration 11: **'cognitive dissonance'**<br><br>If participants discuss and reflect on their work processes (e.g. based on trustworthy data or personal experiences) during a professionally facilitated exchange of positive experiences or failures (C), then they discover knowledge gaps and identify learning needs and relevant topics (O) because their own attitudes and behaviours may differ from their peers', creating cognitive dissonance (a negative emotional state triggered by conflicting perceptions) that makes them reconsider their own way of working (M).<br><br>*During the meetings, the treatment of these specific patient records was discussed, especially differences between what was prescribed according to the records and what was actually dispensed*[11].<br><br>*One of the key features of QCs is that working methods map the quality of care in one's own practice. First of all, this distinguishes QC work from further training in the classical style and second, it enables participants to identify real quality problems in their own practice [translated from German]*[14].<br><br>*The combination of the written simulated cases with actual prescribing allows the GPs to reflect on their decisions as well as the background for these decisions, and is in line with suggestions to make drug utilization studies closer to the reality of practice*[11].<br><br>*Through reflection, a gap between current practice and best practice is recognized. Distinguishing this gap presents an opportunity to identify learning objectives specific to the family practice setting*[10].<br><br>*Our results are in concordance with research that suggests that GPs may feel disappointment if their prescribing practice conflicts with their ideals*[9]. |
| CMO configuration 12: **'social learning'**<br><br>If the facilitator uses purposeful didactic techniques (e.g. brainstorming, contentious or consensus discussions, or role play) to keep the group active and to reward exploratory behaviour during reflection on the work |

process (C), then the group will create a learning environment that promotes knowledge exchange (O) because learning is a cognitive process in which participants observe and imitate their peers' behaviour to gain social approval (M).

*The participating GPs experienced the CME group meetings as an important arena for learning. They reported picking up good advice from others and learning practical alternatives …. GPs said their prescription data would not mirror all learning effects: 'The whole point is to reflect more …'* [9].

*Once the problem is acknowledged, one must learn and understand what caused the problem and how it can be solved. For this, elucidating and discussing the decision process underlying treatment decisions may be useful. To accept new information or practice recommendations, the credibility of the source is of importance*[15].

*Cooperative learning can increase flexibility and joy in medical action (everyone learns from everyone) [translated from German]*[16].

*Learning from and with colleagues is an important source of both new information and strategies for applying that information to practice*[10].

*Cognitive feedback is feedback on the decision process, i.e. why or how a decision is made and not on the decision itself, i.e. which decision is taken*[11].

CMO configuration 13: **'positive interdependence between health insurance companies and GPs'**

If physician network organisations require continuous QC activities (C), then QCs will negotiate priorities and design creative solutions (O) because the tension between autonomy and obligation spurs the group to act and negotiate together to reach a common goal (M).

*The physicians in the Rhine-Main network of physicians committed themselves to participating in QCs when they joined the contract. In QCs, they discuss prescription patterns for specific clinical situations and adapt guidelines to local conditions [translated from German]*[22].

*The participation of German GPs in QCs is mandatory in order to be part of government-funded disease management programmes (DMPs) or to be part of pilot projects with health insurance funds*[23].

CMO configuration 14: **'threat to professional autonomy'**

If GPs feel that the QC programme is only a top-down managerial intervention to reduce costs (C), then they will not be motivated and will not participate (O) because they feel unsafe and fear they lack autonomy in their clinical role (M).

*GPs and facilitators pointed to the difficulty of reaching consensus on a best buy…. Some found the term 'off-putting' because of its financial connotations. This suggests that some GPs may feel that their management decisions should be based on wider considerations than those of cost-effectiveness*[3].

*GPs were also unlikely to take part if they felt that the sessions would make them feel unsafe or if they felt that the sessions were yet another 'top-down' managerial intervention, where the main intention was to reduce prescribing costs*[3].

*The majority of respondents in both regions expected to benefit from participation in QCs but were unwilling to accept the risk that QI could be misused for control or cost reduction [translated from German]*[1].

**Supplemental material 8: literature based programme theory including illustrative quotes and complete list of supporting quotes**

| |
|---|
| CMO configuration 15: **'positive interdependence among group members'**<br><br>If participants maintain an atmosphere of trust in a learning environment that promotes the exchange of knowledge, assisted by facilitators who use professional techniques (e.g. contentious discussion, reaching consensus and role play) (C), then participants will adapt and generate new knowledge for local use (O) because they see themselves as similar, and so act and negotiate cooperatively to reach a common goal (M).<br><br>*The acquisition of new knowledge, skills, and approaches to bridge this gap follows. Often, however, access to new information alone is not sufficient. Reflection and discussion are necessary to help physicians 1) identify areas where current practice requires change and 2) develop strategies to integrate this new approach*[10].<br><br>*There was widespread agreement that the principal requisites for a good facilitator were experience and competence in small-group skills. One facilitator identified another skill: 'You've got to be able to hold the tension between comforting and challenging'*[7].<br><br>*The personal interaction and mutual influence between colleagues implicitly resulted in an individual or group contract*[24].<br><br>*Psychological research into group behaviour has produced an inventory of factors that influence conformity with group standards. Unanimity provides more pressure to conform, while privacy makes it easier not to*[25]. |
| CMO configuration 16: **'identifying and removing barriers to change'**<br><br>If participants, supported by skilled facilitators, address barriers to change (C), then they are more likely to implement the innovation (O) because participants help each other to develop strategies to identify and overcome these barriers (M).<br><br>*Barriers within doctors relate to competence, motivation and attitudes, and personal characteristics such as learning style, whereas barriers within practices exist as doctors do not work entirely independently*[26].<br><br>*Within the group, members endeavour to identify specific barriers to these practice changes and to formulate implementation strategies to facilitate desired changes*[10].<br><br>*The implementation of new knowledge is facilitated by expressing and discussing how to overcome obstacles to its acceptance*[25]. |
| CMO configuration 17: **'need for competence, autonomy and relatedness'**<br><br>If participants create new knowledge and plan an implementation strategy (C), then they feel satisfaction, responsibility and stewardship (O) because this fulfils their need for competence (being able to achieve specific objectives) (M), autonomy (a feeling of being in control of their own behaviour) (M), and relatedness (a sense of connection to a larger group) (M).<br><br>*The decentralised approach at a local, internal level includes participants gathering experience from daily practice and formulating a feasible consensus solution. The advantage of this method is that GPs are actively involved in this process and therefore motivated to implement the (newly) developed guidelines. In addition, the participants involved will be more likely to accept (new knowledge) and feel committed to implement it [translated from German]*[16].<br><br>*Potential advantages of the local approach: it promotes buy-in, maximizes fit to local culture and circumstances, maximizes the ability to work out the details associated with implementation, and produces a highly rewarding experience*[12]. |

**Supplemental material 8: literature based programme theory including illustrative quotes and complete list of supporting quotes**

CMO configuration 18: **'intention to change'**

If participants publicly announce their intention to change (C), then they are more likely to implement the change (O) because they and others in the group all think it is a good idea and believe they can carry it through (M).

*I was surprised to see how willing people were to reflect on their own behaviour and practice e... and constantly make comments like: 'Well, did I really do that? I surely have to pull myself together'. Very strong will, apparently, to make changes*[9].

*The third was the development of individual and group plans for change, to stimulate GPs to really put their plans into daily practice*[20].

*Groups can be more effective in accomplishing tasks, and publicly announcing behavioural changes results in more commitment than private change*[25].

*… draws on 'the theory of planned behaviour' and other studies that have identified the pre-requisites of successful behaviour change in general practice reviewed by Veninga et al. 2000*[3].

CMO configuration 19: **'testing new knowledge'**

If participants validate and test new knowledge in a QC, moderated by a skilled facilitator in a safe environment (C), then they feel confident putting that knowledge to use in everyday practice (O) because they have had the opportunity to practise and familiarise themselves with the innovation (M).

*Interactive approaches, however, can be effective, particularly when they involve participation in small peer groups that foster trust, promote discussion of evidence relevant to real cases, provide feedback on performance, and offer opportunities for practising newly learned skills*[10].

*Understanding application of new knowledge. The discussions helped members to consider translating evidence into practice: 'Sometimes you can read about things but are unable to see how to put it into practice and I feel PBSGL enables you to think how you can do that'*[5].

*Next, they examined empirical evidence concerning the validity of these solutions. To facilitate this process, teams had access to the large resource library that the research team had assembled*[12].

CMO configuration 20: **'gaining confidence in an innovation'**

If the group repeatedly practises implementing and adjusting to an innovation (C), then its members trust their own competence and turn the innovation into a habit (O) because successful outcomes increase their confidence in their abilities (M).

*One meeting may not be enough to actually change treatment, although that is the usual procedure in the peer review groups. Behavioural theories stress the importance of repetition, especially for changing routine behaviour*[2].

*In general, GPs were excited to find in the second year that they had indeed changed in accordance with their plans, and they were then usually more motivated to implement further changes*[20].

*These results demonstrate the need to look at repeating/reinforcing messages at 12–24-month intervals*[27].

*The constant feedback on progress achieved and the further possible improvements are other success factors*[28].

**Supplemental material 8: literature based programme theory including illustrative quotes and complete list of supporting quotes**

| |
|---|
| *The intervention comprised repeated feedback on prescribing routines and an intensive programme of educational small group sessions, as described by Bahrs et al. (2001)[29].* |
| CMO configuration 21: **'repetition priming and automaticity'**<br>If participants build a regular group and practise using QI tools (C), then they will successfully implement new knowledge into everyday practice (O) because responses improve with repetition: 'practice makes perfect' (M).<br><br>*Practitioners develop expertise when they move from their comfort zones to examine problems 'at the upper limit of the complexity they can handle'; they learn, and iteratively gain mastery through cycles of reflecting on practice, obtaining feedback, and adjusting performance[19].*<br><br>*The benefit from participation depended significantly on the frequency of the meetings. Successful projects might not only positively reinforce the introduction of continuous QI, but could also bring about a positive attitude to the other aspects of systematic and continuous quality improvement[30].*<br><br>*Real improvements to performance in daily care can only occur if there is an ongoing and regular quality circle process [translated from German][31].* |

In blue: changed wording

### CMO configurations across papers

#### ***Context mechanism outcome configuration 1: 'participants know what to expect'***

Improved wording: If the introductory workshop conveys the principles of QI in PHC and the workings of QCs (social persuasion) (C), this will increase future participants' motivation to join QCs (O) because they learn what to expect and may feel that they are capable of meeting expectations (increase of self-efficacy) (M).

*The introduction strategy included a meeting with all staff in which the model was explained, a manual on theoretical and practical backgrounds of the model; support in the use of the model and the start of a first improvement project; a one-day course on quality management ....[32].*

*This (small projects) seems to be in accordance with previous findings where improvement of the internal structure is often seen as the first step towards the full adoption of continuous quality improvement. It is sensible therefore to advise practices to start with this kind of improvement project[32].*

*Our findings stress the importance of starting CQI with small, easy-to-handle projects[30].*

*For CQI to be introduced successfully, a positive attitude toward CQI is required from all who will be working with it[30].*

*(They) learned how to organise the meetings, how to guide the members of a peer group through the steps of the quality circle, and how to deal with group processes[33].*

*Introduction of the model … is important for understanding and helps participants during the start of the process. It was also vital to have a common and shared understanding of the problem among participants[2].*

**Supplemental material 8: literature based programme theory including illustrative quotes and complete list of supporting quotes**

*In September 1992, 200 general practitioners and internists of a defined postal code area were contacted by the Kassel district office of the Kassenärztliche Vereinigung Hessen and invited to an information event [translated from German][34].*

*It might be better to provide targeted information in advance of the project at an information event. This would make it easier for potential participants to decide for or against participating in the project, since questions as well as fears and reservations can be clarified immediately [translated from German][31].*

*Surveys have revealed far-reaching gaps in information, some of which are the cause of misunderstandings and misjudgements. In particular, the working methods and objectives of medical quality circles are apparently insufficiently known. Better general information on this subject, which contracted doctors in particular, expect from their KV (health insurance company), is therefore urgently needed. As examples from the Netherlands and Great Britain show, active information from the target group is a basic prerequisite for quality-enhancing measures in practice [translated from German][1].*

*Introduction of the model … is important for understanding and helps participants during the start of the process. It was also vital to have a common and shared understanding of the problem among participants. It is worthwhile taking the time for an agreement on shared guidelines …….[2].*

*To deal with these issues (information overload), an initial, introductory session of 'Reflective Practice' needs to be included, where GPs' experience of previous prescribing management interventions can be aired, where safe 'rules of engagement' can be agreed, and the purpose of the 'reflective practice' intervention made explicit[3].*

*A more structured introductory meeting that would assess participants' learning needs, negotiate the future content of the small group meetings, seek agreement on learning agenda, dates, times and venues, establishing communication channels and explicitly discussing the educational rationale[35].*

#### *Context mechanism outcome configuration 2: 'need for autonomy and obligation''*

Improved wording: If the administration at the national level or at the level of health insurance companies entrusts GPs with QI and autonomy (puts them in control of how do it) (C), then GPs may consider participating in QCs (O) because they feel they can take on the responsibility and make a difference (M).

*Social Law Code has given new impetus to the obligation of the associations of statutory health insurance physicians to implement quality assurance measures. As early as 1991, the board of the Kassenärztliche Vereinigung decided to introduce nationwide quality circles as an instrument of quality assurance in outpatient care [translated from German][34].*

*In January 1993, the Association of Statutory Health Insurance Physicians in Southern Baden constituted an interdisciplinary working group with the aim of developing the organisational and conceptual framework for the establishment of quality circles in the Southern Baden region [translated from German][36].*

*The participants expressed their fears that participation in the quality circle could lead to possible regulation by KV or health insurance companies [translated from German][37].*

*The respondents are suspicious of an obligation for all physicians to participate in quality assurance measures. In Saxony-Anhalt in particular - as shown by the clear statements made by those surveyed - this scepticism is linked to the consideration that a commitment to quality assurance measures would be more acceptable if it also affected those colleagues who refrain from continuous medical education training [translated from German][1].*

**Supplemental material 8: literature based programme theory including illustrative quotes and complete list of supporting quotes**

*In 1993, the health structure law ('Gesundheitsstrukturgesetz') added more specific recommendations to the existing body of rules about quality assurance with the explicit aim to stimulate quality assurance programs (quality circles) in primary and hospital care[31].*

*The participation of German GPs in QCs is mandatory in order to be part of government-funded disease management programs (DMPs) or to be part of pilot projects with health insurance funds[23].*

*Furthermore, some differences between the regions could be observed. In region 1 the impact seemed highest, which may be explained by the activities of the Association of Statutory Health Insurance ('Kassenarztliche Vereinigung') in that region regarding continuing professional education[38].*

*Research evidence showed that budget constraints could reduce prescribing volume and costs (14 Sturm H 2007)[38].*

*In the discussion with facilitators, the QC participants expressed their desire to be self-determined and work independently. For this reason, the Federal Association of Statutory Health Insurance Physicians (KBV) and the Associations of Statutory Health Insurance Physicians (KVen) have laid down the thematic and methodological autonomy of QCs as a prerequisite in their guidelines for quality circle work and have committed themselves to supporting them [translated from German][4].*

#### *Context mechanism outcome configuration 3: 'sharing similar needs'*

If the administration at the organisational level of QCs provides administrative support (i.e. for training facilitators, data gathering, provision of evidence-based information), and the administration protects time and space, and offers CME points, and small financial incentives to QC participants (C), then they will meet in groups to exchange ideas (O) because GPs prefer learning in QCs (M); support generates positive expectations among participants (M) and GPs think QC meetings with their peers will be useful (M).

contextual features at the organisational level:

*With a restricted although realistic budget, facilitation should be set up as efficiently and effectively as possible[32].*

*We have a very busy schedule most of the time, leaving little or no time for extra work[30].*

*It was mainly a logistics problem. We have little room in practice[30].*

*We already had so many meetings and we have so many tasks to fulfil. I work in a health care centre[30].*

*I have often postponed things knowingly. Sometimes the bucket just overflowed[30].*

*Staying close to the needs and expectations of the practices could be a way to introduce continuous quality improvement more effectively[32].*

*We also found that the available time and possibilities to plan activities well were felt to be the most important barriers to using the CQI model[30].*

*When there are great obstacles to change (e.g. limited time, the need to acquire a new skill), the group might decide to set aside time to specifically address strategies for overcoming these barriers[10].*

#### Supplemental material 8: literature based programme theory including illustrative quotes and complete list of supporting quotes

*As a discussion platform, we developed special facilitator manuals according to a uniform didactic concept. It includes relevant clinical portraits (sleep disorders, back pain, upper abdominal pain, heart failure, etc.) in general practice. These materials provide the facilitators with guidance and make it possible to stimulate and supplement the problem-oriented discussion of the diagnostic and therapeutic procedure at critical points. ...In our opinion, the advantage of this approach is that it makes it easier to get started with concrete quality circle work and that quality circles can be implemented on a broad basis [translated from German][36].*

*External support should help with the administrative organisation of QCs, as this exceeds the time capacity of the facilitators [translated from German][1].*

*The majority of respondents (85%) [HB: 87.1%] want or even urgently demand support for quality circle work from their Association of Statutory Health Insurance. In Saxony-Anhalt, the vast majority of respondents want both organisational support (e.g. by making rooms available, making contacts and making those contacts available - "start-up on site" - and granting reimbursement of expenses) and content-related support (e.g. by providing materials, topic recommendations, arranging speakers). In Bremen, primarily organisational support is expected [translated from German][1].*

*The fact that all groups are led by recognised (i.e. trained) facilitators speaks for the existing structural quality. The high level of continuity and frequency of participation also suggests that structural conditions such as accessibility, suitable conference room and location, clear scheduling, etc. are in place [translated from German][14].*

*In some cases, the KVs took different approaches, for example by organising external facilitator training courses, developing special structure of QC meeting or supervision of facilitators [translated from German][4].*

*In addition, long-term maintenance of small groups implies a national support for CME in general practice with enough personnel and economic resources to assist all those GPs who have key roles in providing CME at the local level[8].*

*Participation, ..., does not come without costs. ..., it is time consuming, .... For clinicians, who often see patients continuously throughout the day, it can be especially difficult to find time to participate in QI efforts[12].*

*... substantial organizational resources .... ..., including tools that the QI teams could use to develop their programs and the costs of the local approach facilitator. ... HealthOrg covered some but not all of the time that participants spent outside of formal meetings, ...[12].*

*Staying close to the needs and expectations of the practices could be a way to introduce continuous quality improvement more effectively[32].*

*The peer groups met on a regular basis according to their needs[33].*

*...as a so called "clinical theme-course", which will give the participants important CME credits[13].*

*In Norway, specialists in general practice must renew their clinical specialty every five years. In this renewal process, participation in a number of peer CME group meetings are compulsory, in order to stimulate a continuously medical education and reflection[39].*

*CME group members earned CME points to renew their speciality[9].*

*General practitioners (GPs) favour learning environments such as reading journals, discussion with colleagues, and participation in quality circles[9].*

**Supplemental material 8: literature based programme theory including illustrative quotes and complete list of supporting quotes**

*GP specialists have to renew their specialty every 5 years. Recertification demands participation in a number of peer CME group meetings. Typically, a peer CME group comprises seven or eight GPs who set up their own educational programme for monthly evening meetings*[40].

*The strategy also fts in well with the work setting of many GPs in European and non-European countries, which are often characterised by small practices, relatively isolated settings and a desire for more contacts with peers*[20].

*The innovative, multifaceted strategy for improving test ordering behaviour was favourably evaluated by a large GP population. All local GP groups expressed a desire for continuation of the meetings after the experiment*[20].

*GPs appreciate the combination of individual feedback, discussions about guidelines and small group quality improvement meetings driven by peer influence*[20].

*Success rates of specific strategies seem to be strongly influenced by the extent to which they fit in with the local and organizational context and the physicians' day-today work routines*[24].

*The first success was the easy recruitment, with practice groups eager to participate in the trial*[24].

*…these groups of professionals practising in the same region meet regularly to discuss treatment, pharmacotherapy, and patient management*[25].

*We have also arranged for CME credits, needed to fulfil the educational requirements of ongoing licensure*[6].

*General practitioners can discuss topics relevant to day-to-day practice. They get access to a local expert … l. Since topics come out of their own and their peers' practices, and are discussed by the expert, it is more likely that perceived and unperceived needs will be addressed*[6].

*…that a small group format might be more attractive than other forms of CME, since this has been our experience*[6].

*This learning format may meet a need for practices that have protected learning time to enable them to use multi-professional group learning to its full advantage*[41].

*PBSGL enabled participants to compare their practice with that of their peers, and this was mentioned frequently as a very positive motivator in joining and continuing in the groups*[7].

*…with surprisingly few opportunities to gauge themselves and their practice against their peers, and they have been found to value this opportunity highly*[7].

*The most-cited reason for joining PBSGL … as the PBSGL format matched their preferred learning style. …. Keeping up-to-date in clinical practice was the second-most mentioned reason*[5].

*The most significant outcome did not come from the evaluative data collected during the research; rather that both groups are continuing to meet more than six months after the pilot finished*[42].

*PBSG enabled participants to compare their practice with that of their peers, and this was mentioned frequently as a very positive motivator in joining and continuing in the groups. This corroborates previous work which found this to be an enhancer for translating research into practice*[7].

*The reasons for participation varied and ranged from overcoming the lone fighter situation in the practice, defining the image of the family doctor, possibilities and limits, to searching for practical solutions to everyday treatment problems [translated from German]*[34].

**Supplemental material 8: literature based programme theory including illustrative quotes and complete list of supporting quotes**

*The most frequently mentioned motives for participating in quality circles were practical help and exchange of experience (57 and 58 mentions) [translated from German][21].*

*The vast majority of participants cite the collegial exchange of experience as the greatest motivating factor for working in a quality circle. The primary goal is to improve the collegial relationships. At the same time, the desire for more consensus in medical action and the improvement of skills in diagnostics and therapy is mentioned as a very important objective [translated from German][16].*

*The main motives for participating in quality circles were the expectation of practical help for one's own practice, inter-collegial exchange of experience, improvement of patient care and opportunities for self-reflection on one's own work as well as personal support. Competing time commitments and above all the fear of external controls were mentioned as obstacles to participation in quality circles [translated from German][1].*

*Quality assurance in outpatient care was considered necessary - even more so in Saxony-Anhalt than in Bremen [translated from German][1].*

*External staff should organise QCs as facilitators have too little time to do this [translated from German][1].*

*The summary makes it clear that the question of participation in a quality circle is primarily based on specific medical needs. Many physicians wish to receive practical assistance in their daily practice and wish to overcome the structurally dependent professional and emotional isolation through intercollegial exchange. The most important goal is therefore personal support [translated from German][1].*

*Overall, more than 86% of the participants were (very) satisfied with the work in the quality circle. In contrast, only 2.8% were dissatisfied and 0.4% very dissatisfied [translated from German][43].*

*For almost all participants (97.1 percent), the desire to analyse their own prescribing behaviour and to optimise it with the help of the prescription data evaluation of colleagues was at the top of the list. The exchange of experience with colleagues and the expansion and refreshing of knowledge regarding pharmacotherapy were also considered important [translated from German][44].*

*Data from older surveys showed that family physicians indicated colleagues most often as information sources, followed by journals and books…. The most important requirements for media in medical education as perceived by the participants were its relevancy for daily practice and dependability[23].*

*… we … predict that German general practitioners … favour the "classical" learning environments such as: journals, colleagues, and quality circles. journals and books. … exchange ideas and discuss actual trends with colleagues collegial and interactive rather than to meet experts …[23].*

*The second key area of expectation was with the promotion of collegial exchange: more than conventional further-training events, quality circles assumed that a special form of group work by doctors would be a way of overcoming isolation in the private practice [translated from German][1].*

*To ensure attendance in the future, the educational sessions need to be protected by the use of a paid locum, in the same way as other practice development work is now being supported[3].*

*The workshop was based on a provincial learning needs assessment and data from focus groups of family physicians from each of the provinces to ensure the curriculum material would meet the needs of physicians across Canada[45].*

*One of the strengths of the programme is its adaptation to the needs of GPs and pharmacists [translated from French][46].*

*GPs' participation in PPQC meetings is accredited by the association for their continuing education[28].*

*when asked, GPs also express a need for drug information/education that is academic and not promotional*[47].

*The participants were not offered any extra incentives, except for the education itself*[47].

*Doctors learn best when they recognise the need for learning and when learning is self-directed*[8].

*Many studies have shown that small group sessions are one of the most popular and stimulating CME activities practised by doctors*[8].

*The idea of problem-based and self-directed learning from everyday practice, closely linked to quality improvement, seemed to appeal to many Swedish GPs and the CME programme was successively accepted by the majority of them*[8].

*A meeting attendance fee was paid to the GPs, €70/hour for a plenary meeting (with the consultants), €45/hour for a quality circle meeting*[48].

*The brief qualitative responses indicated that participants chose to join the small groups mainly because …there is a better rapport between the individuals and one gains more than just attending a lecture*[35].

*The importance of a needs-identification process and the involvement of the programme user group in this process have been identified as crucial factors in the success of any effective learning programme*[27].

*Beginning in 2005, attendees received category-I CME credit*[19].

#### *Context mechanism outcome configuration 4: 'need for relatedness'*

Improved wording: If a steady group of members engages in socially enjoyable contact, led by a skilled facilitator who, e.g., introduces people to each other, opens discussions, clarifies and summarizes statements (C), then group members will get to know each other and norm rules that they are willing to follow and build safe environment based on trust (O), because members want to be among and to interact with equals (M).

*The groups were different ... we thought that a group is a group and all we have to do is to run the scheme ...and then I experienced that groups have their own cultures. These groups have existed for a while, which we probably have to consider ...*[9].

*Tutors did not consider themselves as "experts" but as "one of them"., Being open about their background as GPs was an agreed upon strategy, and tutors deliberately tried to avoid being perceived as experts: The tutors experienced that their own background was important for GPs' trust and acceptance*[9].

*Both tutors and GPs emphasised that a "good atmosphere" in the group, and "a sense of security" among group members was essential for an open and constructive discussion*[9].

*Membership in the peer review group has been stable over time because it is unusual for general practitioners (and their patients) to switch between groups*[2].

*CME has focused on disseminating information, but it has become increasingly clear that acquisition of knowledge is less important in changing physicians' behaviour than the social context of learning. Habit and custom, the beliefs of peers, and social norms are the major determinants*[49].

*Sessions are generally held in the evenings with a meal*[6].

**Supplemental material 8: literature based programme theory including illustrative quotes and complete list of supporting quotes**

*We estimate it took three to four sessions for the group to be comfortable with this process. Open discussions and debates then came more freely, and the group continued to gel*[6].

*The need to maintain the appearance of competence may be more compelling than the need to learn. Several strategies …. First, we tried to create as relaxed an atmosphere as possible. We arranged tables in a circle, removed all barriers …, and held the sessions with a meal*[6].

*The group and the interactive format are fun*[6].

*Initially, problems with group functioning were anticipated, but they are … uncommon. Groups of various compositions function effectively in this particular small group environment. … heterogeneous groups might provide broader practice experiences and greater variety in potential solutions to practice problems…*[10].

*Participants liked the inclusive nature of the small groups and appreciated the egalitarian quality of the interaction within them*[7]. *[No hierarchy]*

*It didn't matter where we came from; Skye, Wick or Brora. It soon became clear that we were all in the same learning position. And those in Inverness and Aberdeen didn't have all the answers*[7].

*When the expert comes in, learning stops. … The use of invited experts (invariably hospital-based consultants using a traditional didactic approach to learning) was seen as an anathema to adult learning and the small-group ethos*[7].

*Members of both groups described the meetings as relaxed, friendly and informal. The facilitators played a crucial role in creating the atmosphere: …, it would seem that the group members also contributed to the positive climate*[5].

*Reasonably quickly I relaxed. Everybody was keen to make it a success. The group opened up and there was a sense of calm. The positive atmosphere enabled members to be open about knowledge gaps and to ask questions*[5].

*The two groups appear to be at different stages of development. Group 1 seems to have developed a strong sense of cohesion quite quickly compared to Group 2*[5].

*Interestingly, the stage of storming, which is characterized by interpersonal hostility and conflict, was not evident in either group*[5].

*Norming reflects the development of group cohesion, openness and emotional support. The positive social dimension enabled the group to perform – that is, to focus on the task at hand, with resulting effectiveness*[5].

*To encourage GPs' engagement, all sessions took place over lunchtime, and a sandwich lunch was provided. GPs gained PGEA accreditation for their participation*[3].

*..., two practices have instituted a regular morning coffee break, which was described in positive terms as a discussion: "The indigestion .. has come up, and we have a coffee break and quite often discuss clinical things and some comments have come out about that"*[3].

*The success of group learning between GPs within a practice depends to a large extent on the quality of relationships within the group. Where individuals feel that their management decisions are under threat from colleagues with whose judgements, they are not comfortable, discussion may be abruptly curtailed*[3].

*Most participants stressed the benefits of the intervention for facilitating discussion, which was implicit in the design of the educational intervention. This seemed to counteract the convention of autonomous working practices by GPs, which can lead to professional isolation, even in partnerships*[3].

**Supplemental material 8: literature based programme theory including illustrative quotes and complete list of supporting quotes**

*(GPs) in non-academic settings have few safe and reliable forums where they can reflect and learn from the clinical dilemmas inherent in their work[19].*

*"Being with colleagues" … yield four subthemes: (1) gaining renewal through reflection, (2) obtaining others' perspectives, (3) developing collegial trust, and (4) learning specific information/skills …. Over half of the respondents commented on time issues related to participation; a third saw time constraints as deterring attendance[19].*

*They will facilitate the discussion …, based on the individual feedback reports, enabling participants to compare own prescription patterns …… This will probably trigger discussion …, aimed at critical reflection towards own prescription strategies for elderly patients and facilitating disclosure of areas where individual improvements may be desirable[39]. [Facilitation]*

*Facilitators …. trained …. over approximately 20 training hours. Facilitators provided … opportunity for all participants to ask questions …. We … encouraged participants to discuss their …practice patterns. The facilitator …redirected conversations that moved off topic, calmed the skeptics, and encouraged quieter participants to share their personal experiences[6]. [Facilitation]*

*In the mature group, the facilitator's major role is to introduce the expert to the group and the process, and to provide some closure at the end of the meeting[6].*

*… tasks of the facilitator are to focus discussion … to encourage the group to identify factors that ... hinder implementation of new knowledge … …. To successfully fulfil this role, facilitators ...establish a safe, supportive environment ... identify practice gaps and encourage the discussion of sensitive ... issues[10]. [Facilitation]*

*The role of the facilitator has been recognised. He/she needs to be competent at many tasks including opening the discussion, clarifying, summarising, questioning, and devising strategies to improve group function[7]. [Facilitation]*

*There was widespread agreement that the principal requisites for a good facilitator were experience and competence in small-group skills. One facilitator identified another skill: 'You've got to be able to hold the tension between comforting and challenging[7]. [Facilitation]*

*The facilitators were also skilled in encouraging participation[5]. [Facilitation]*

*'I think you need a facilitator certainly need it for the donkey work of the, arranging the meeting and making sure everybody has the module[17]. [Facilitation]*

*'It gives you licence to play devil's advocate as well and challenge people a bit more whereas if you were always doing that as just a group member, people might think you were just doing it to annoy them[17]. [Facilitation]*

*Participants considered that one-to-one mentorship with an experienced or established PBSGL facilitator would be very beneficial. This was also suggested as a method to encourage members of existing groups to train as facilitators[17]. [Facilitation]*

*Any anxieties that potential facilitators may feel, mainly the concern that a new group will be hard to form, or will be dysfunctional, – need to be discussed with potential facilitators before and during the initial training. Facilitators of such groups will need the most support …[17]. [Facilitation]*

*In countries using PBSGL, national networks provide training for facilitators and supporting material for the groups[10]. [Facilitation].*

*The facilitator is selected by the group[10]. [Facilitation]*

**Supplemental material 8: literature based programme theory including illustrative quotes and complete list of supporting quotes**

*The reasons for participation varied and ranged from overcoming the lone fighter situation in the practice, defining the image of the family doctor, possibilities and limits, to searching for practical solutions to everyday treatment problems [translated from German][34]. [Facilitation]*

*Our group prepared the facilitators for their task in two one-day training sessions. They had to conduct a model quality circle and critically discuss their role based on a pre-developed manual. In the second training course, we taught them important basic knowledge of group dynamics and basic didactic skills for their role as a facilitator [translated from German][36]. [Facilitation]*

*The vast majority of participants cite the collegial exchange of experience as the greatest motivating factor for working in a quality circle. The primary goal is to improve the collegial relationships. At the same time, the desire for more consensus in medical action and the improvement of skills in diagnostics and therapy is mentioned as a very important objective [translated from German][16].*

*The summary makes it clear that the question of participation in a quality circle is primarily based on specific medical needs. Many physicians wish to receive practical assistance in their daily practice and wish to overcome the structurally dependent professional and emotional isolation through intercollegial exchange. The most important goal is therefore personal support [translated from German][1].*

*1-2 doctors from each group took on the task of the facilitation. AQUA employees trained and supported them during the course of the project. They also prepared facilitation materials and provided organisational support [translated from German][43]. [Facilitation]*

*The groups were moderated by a primary care physician, who had had a 2-day training on moderation of quality circles and who received supervision in about two sessions per year. One session per 1 or 2 months was planned[38]. [Facilitation]*

*For this purpose (tutor system to support the moderators) 50 experienced facilitators were trained as quality circle tutors, who have been responsible for the training and further training of facilitators since 2001. In 2002 the KV Westfalen-Lippe followed this concept and trained 30 tutors. The encouraging experiences from both projects led to the KBV introducing the concept at a federal level in 2003. Since then it has trained 116 tutors [translated from German][4]. [Facilitation]*

*In order to support the facilitators in the design of circle meetings, the KBV has developed structured didactic handouts for circle work, so-called quality circle manuals.... The materials are to be understood as recommendations.... [translated from German][4]. [Facilitation]*

*The (facilitator) training usually lasts two days, i.e. between eight and 16 hours, usually twelve hours. ... In all twelve KVs, the trainers use the Quality Circle Handbook and their manuals. Further training for facilitators usually takes place in one day and lasts between three and ten hours [translated from German][4]. [Facilitation]*

*The CQC facilitators were local family physicians recruited and trained specifically to lead study meetings. They were chosen by the CQC steering committee for their skills in facilitating small-group activities, their known interest in chronic disease management, and their involvement in continuing professional development[45]. [Facilitation]*

*Before the meetings, train-the-trainer workshops were conducted to assist facilitators in their role as group leaders[45]. [Facilitation]*

*Facilitators were local family physicians recruited to lead and initiate discussion at study meetings and were chosen because of their skills in small group facilitation and involvement in continuing professional development and were selected by the study's steering committee[50]. [Facilitation]*

**Supplemental material 8: literature based programme theory including illustrative quotes and complete list of supporting quotes**

*… facilitation skills and aptitude were ... important … …. As one of the GPs commented …: "I think the person's much more important than their background." A facilitators' ability to manage a group successfully was central. … a good facilitator should "... whipping us into line"*[3]*. [Facilitation]*

*The ability of the facilitator to manage group discussions, … to create an atmosphere that was non-threatening and supportive. … willing to challenge the group when members colluded with one another to evade potentially contentious issues*[3]*. [Facilitation]*

*The respect of the group for the facilitator was crucial to the success of the intervention. Facilitators needed to be grounded in a sound knowledge of prescribed medicines, but also needed to have group facilitation skills*[3]*. [Facilitation]*

#### *Context mechanism outcome configuration 5: 'need for autonomy and control'*

If the group chooses its own topics and facilitator (C), then they will feel they own the QC (O) because their need for autonomy is satisfied (a feeling of being in control of their own behaviour) (M).

*Tutors did not consider themselves as "experts" but as "one of them". Being open about their background as GPs was an agreed-upon strategy, and tutors deliberately tried to avoid being perceived as experts. The tutors experienced that their own background was important for GPs' trust and acceptance*[9]*.*

*The extra benefits gained by using GPs instead of non-physicians as (facilitators) have also been reported in a Dutch study*[40]*.*

*A final limitation, caused by the study design is the fact that the peer groups did not have the opportunity to choose their own topics, …. After reading and discussing the content of the workbooks the peer-review groups defined self-selected change objectives*[33]*.*

*A bottom-up approach to CQI stands central, along with an active role for the practice team and the application of a clearly structured, stepwise problem-solving method to develop and implement the improvement plans*[51]*.*

*it is crucial to the model that practice teams formulate goals for improvement and attempt to achieve these goals in small scale*[32]*.*

*Reasons that were reported most often included "the subject chosen was felt to be a problem or a bottleneck in practice management", "the practice wanted to implement the national guidelines (on that specific topic)", and "the outcomes of the audit report"*[32]*.*

*As practices were free to select their own topics for improvement and set their own objectives, the fact that the intervention group met a significantly greater number of self-defined improvement objectives than the control group is an important finding*[51]*.*

*it consists of involving all staff, holding regular meetings on quality, designating a quality coordinator, and writing annual plans and reports on quality improvement*[32]*.*

*…. were willing to continue using the model, but were less positive about the quality cycle and preparing an annual report*[30]*.*

*(many physicians) …. felt that activities not directly related to practice work*[30]*.*

*The groups themselves generate topics for modules, with the subsequent module being authored by a GP*[10]*.*

**Supplemental material 8: literature based programme theory including illustrative quotes and complete list of supporting quotes**

*Each group decided the frequency, timing and location of the meetings at the first introductory meeting. Each group also decided their preferred method for module selection[41].*

*The facilitator is selected by the group[10].*

*The same publication points out that GPs – due to the lack of therapeutic consequences – do not seriously wish to diagnose the illness[52].*

*The group established common criteria for carrying out an inventory of needs using a standardised form of documentation of the QC process [translated from German][34].*

*This (negative) assessment (of QC work) could be an expression of resistance and reservations regarding the background of the project and gaining participants, and thus an implicit plea for voluntariness and self-determination as the most important characteristic of medical QC [translated from German][37].*

*The main focus of our analysis is on the characteristics of successful quality circle work that can be derived from theory, as they are also laid down in the above mentioned quality assurance guideline: group constancy and continuity, experience-based work on self-chosen topics, collegial group climate and goal orientation towards quality promotion in one's own practice [translated from German][14].*

*The participants of the circle determine the questions concerning the content themselves [translated from German][4].*

*It is important for a learner to be in control of his or her learning process, to be motivated, and to perceive meaningfulness[11].*

*At the beginning … GPs were induced to attend … with criticism. At first, … GPs participated somewhat reluctantly 'in order to avoid trouble', but over time most of them began to look forward to regular attendance and enjoyed … opportunity for an exchange … in a relaxed setting[53].*

*The rise of evidenced-based medical guidelines … decreases individual providers' autonomy. Physicians have raised similar concerns about threats to the autonomy of their profession …. It is within this context, … declining perceived autonomy for …. physicians, … we compare the participatory local and central expert approaches to QI[12].*

*Topic identification is collaborative, end-user driven and uses local data, literature review and input from small group members Groups are peer-led and membership is ...[27].*

#### *Context mechanism outcome configuration 6: 'size of the group affects communication'*

Improved wording: If group size exceeds 15 (C), then interaction among group participants decreases (O) because participants cannot keep up with all participants and follow their conversations (M).

*All GPs participating in such peer groups, on average consisting of six to eight peers, located in southern Norway[13].*

*The mean group size was 7.4 ± 2.7[20].*

*Our group usually has 12 to 15 participants, an expert, and a facilitator. We are primarily composed of generalists and family physicians, but regularly invite a pharmacist and a representative from the sponsoring pharmaceutical company[6].*

**Supplemental material 8: literature based programme theory including illustrative quotes and complete list of supporting quotes**

*Groups of 4 to 10 family physicians form a PBSG in their own communities, meeting for an average of 90 minutes once or twice a month at an agreed upon time and place, allowing time off for holidays and summer vacations*[10].

*How can QCs be supported? (Table 2) Group sizes > 15 or < 5 - are problematic and participants need support [translated from German]*[14].

*A maximum of 15 physicians in each geographical area were enrolled into each circle in the study*[50].

*Thus, it is a stable and voluntary group of five to eight doctors who meet about five times a year with a pharmacist, expert and facilitator, in a context of interdisciplinary continuing education [translated from French]*[46].

*A quality circle is a stable group of 3–10 GPs with … 1 trained pharmacist. Pharmacists volunteer as facilitators and are responsible for motivating local GPs to participate. They … organize the practical … (e.g., rooms, agenda) and get the prescribing profiles of the participating GPs)*[28].

*GPs had to join as groups; c) groups had to be pre-existing; d) the preferred group size was three to six*[11].

*The effect of the educational asthma programme was partly modified by the group size; prescribing behaviour for (asthma) exacerbations improved more in smaller groups. The group size varied from 4 to 13 ……. This result is ……. an optimal group size of 5 to 6 group members*[15].

*Although the optimum number of participants for quality circles is between eight and 10, when necessary, up to 16 per circle were allowed*[53].

*These are groups of ~10 GPs who meet monthly to discuss topics related to clinical practice. Group membership was constant and members of the same practice were grouped together where possible*[27].

#### ***Context mechanism outcome configuration 7: 'feeling safe and not vulnerable'***

Improved wording: If participants trust each other (C), then they can disclose how they work and also the holes in their knowledge (O), because they feel safe rather than vulnerable (M).

*I was surprised to see how willing people were to reflect on their own behaviour and practice ... and constantly comment like: "Well, did I really do that? I surely have to pull myself together". Very strong will, apparently, to make changes*[9].

*GPs generally experienced the CME group as a safe setting to present and discuss their feedback reports: It would have been more embarrassing if it had been in a large lecture hall or a large seminar*[9].

*A shared understanding of the complex decision-making involved in prescribing in general practice was reported by both GPs and tutors as essential for an open discussion in the CME groups*[9].

*GPs generally experienced the CME group as a safe setting to present and discuss their feedback reports*[9].

*After a while, it may become less needed, because participants may then feel more safe about discussing their own behaviour within the group as a whole*[20].

*…greater insights into and discussion of the physicians' own performance in a safe group of respected colleagues would be a powerful instrument to improve the quality of test ordering*[24].

**Supplemental material 8: literature based programme theory including illustrative quotes and complete list of supporting quotes**

*What have you gained from participating in this practice-based small group learning project? small group support: the group works effectively together and as time progressed, I was able to participate more effectively as my confidence grew*[41].

*…. particularly when they involve participation in small peer groups that foster trust, promote discussion of evidence relevant to real cases and provide feedback on performance*[10].

*In time, group members develop confidence and security in the group, rendering the disclosure of ignorance and ''blind spots of knowledge'' easier. Group members could either use the whole group or parts of it to assess their own learning needs*[8].

*It became evident that the only environment in which this intervention could flourish was one that was safe and interesting …* [3].

*GPs were also unlikely to take part if they felt that the sessions would make them feel unsafe*[3].

#### *Context mechanism outcome configuration 8: 'need for competence and self-actualisation'*

If the facilitator supports participants and encourages them to tell their stories and share their experiences in a safe environment, e.g., by encouraging interactive responses, through discussions and by summarising statements, (C) then participants will be involved and share their positive experiences and failures (O) because they want to improve their professional competence, (M), gain professional confidence (M) and fulfil their professional potential (M).

*…. that an improvement in prescription behaviour could be obtained in a group setting where the participants knew each other well and were used to discussing challenging topics related to their own clinical practices*[54].

*Both GPs and tutors experienced that sharing the experience of being a GP contributed to an open and constructive discussion*[9].

*Reflective thinking increased among GPs; they were able to reflect their individual prescription habits in the CME group. Inappropriate results could put some GPs in distress in front of the group (Frich et al., 2010)*[9].

*Another important topic of debate was how to deal with the frequent requests by patients to have inappropriate tests performed*[55].

*The decision to focus on clinical problems instead of tests was a good choice, since it allowed the feedback and group work to be linked to national evidence-based guidelines. GPs appreciated this approach, because it was also closely related to their everyday work routine*[20].

*There is some empirical evidence that participating in quality circles may increase GPs' job satisfaction*[20].

*Various members expressed a desire to keep up to date …. Others wanted to compare what they were doing with their peers, to confirm that they were practising safely …. Participants …stated that they wanted to be able to examine current evidence and to improve their critical appraisal skills*[7].

*The need to maintain the appearance of competence may be more compelling than the need to learn. Several strategies …. First, we tried to create as relaxed an atmosphere as possible. We arranged tables in a circle, removed all barriers …, and held the sessions with a meal*[6].

**Supplemental material 8: literature based programme theory including illustrative quotes and complete list of supporting quotes**

*The facilitator elicited interactive responses … with the aid of specific predetermined prompting questions and responses. The program participants resolved practice-based problems …. The best practices were determined by the group as a whole and conflict resolution was achieved with the mediation of the content expert, if required*[56].

*The cases were regarded as not only appropriate but also reflecting practical problems in office practice*[56].

*...the success of this format depends on availability of course material that reflects practice based clinical problems and on the important roles of specially trained facilitator*[56].

*The ability to change practice is enhanced if skills are endorsed by trusted colleagues and supported by published literature, and there is opportunity for practice and feedback*[6].

*The ability to change practice is enhanced if skills are endorsed by trusted colleagues and supported by published literature, and there is opportunity for practice and feedback*[6].

*An interactive small group can prompt moderately large changes in physician practice*[10].

*..comparison with one's peers was important, as was the support, confidence and reassurance that some gained from being part of the group*[7].

*I've gained more confidence because of spending time with these people. To go in [to PBSGL meetings], be with these fellow professionals, but it was completely calm, completely non-judgemental*[5].

*We recognize that many personal, professional, and social forces affect attendance at CME beyond the format itself*[6].

*Network (SIGN) on a variety of clinical and non-clinical topics. 'Modules are much better than SIGN guidelines because they are patient based and make you think about your own practice*[7].

*…discussion of personal stories might help participants tackle any doubts they may have on individual cases, and it might also enable attitudes to be highlighted and perhaps modified, through hearing the views and beliefs of others. …, the group members and the facilitator may …. offered each other educational support*[7].

*Comparison with one's peers was important, as was the support, confidence and reassurance that some gained from being part of the group*[7].

*Specific assistance and solutions for actual problems in their own practice are sought and willingly accepted. Finally, a decisive factor for the motivation to work in the case-oriented QZ is the emotional relief reported by all participants [translated from German]*[21].

*It became clear that one's own actions are influenced less by the appropriate clinical knowledge than by one's own experiences, attitudes, and interaction with patients [translated from German]*[34].

*Each participant described his or her own case of how a family doctor deals with their own sore throats or family members' complaints [translated from German]*[34].

*The possibility of overcoming isolation in one's own practice, a way out of isolation, as well as the experience that others have similar problems structurally and they are not different from anybody else, seems an emotional relief. Even more, the reawakening that medical action (e.g. active listening to blood pressure measurement) can be helpful and positive [translated from German]*[21].

*Exchanging experiences in QCs, GPs can work out and clarify the characteristics of the general practice, which improves knowledge transfer [translated from German]*[36].

#### Supplemental material 8: literature based programme theory including illustrative quotes and complete list of supporting quotes

*By working within a QC, I have received more emotional support for my daily practice. The QC work should offer help with disputes/arguments and emotional relief when, comes to, for instance, very expensive therapies [translated from German][37].*

*The basic message was that quality circles are necessary because they promote collegial cohesion more intensively than normal training events. It is extremely important for the individual to know that their colleagues share the same problems or experiences that he or she has [translated from German][18].*

*....it would be easier to conduct a conversation (e.g. when dealing with desired prescriptions); a topic that is always relevant for different indication areas and where many people seem to have benefited from the exchange of experiences and the group discussion. Probably, they felt strengthened by the support from colleagues and the enhanced self-image as GPs [translated from German][43].*

*....in the sense of a continuous, systematic, goal-oriented, facilitated exchange of experience on the basis of specific everyday actions in practice [translated from German][14].*

*At the first meeting the GPs discussed in groups how they diagnose the illness, and the underlying reasons they find important when deciding on treatment[57].*

*Subjects, topics and cases discussed in groups come from daily work and are highly relevant to practice. The small group will meet the demands of developing generalist knowledge as well as the expert role in general practice[8].*

*Small groups will have opportunities to discuss the ''art of medicine'', founded upon context, anecdote, patient stories of illness and personal experiences. Accepting emotional responses being mirrored by other group members corresponds in some respects to the process in Balint groups[8].*

*The group should act as a forum where its members can reflect freely upon all problems that bind them together in their profession[8].*

*…GPs regard the group as a place for social support, …., growth in the professional role …for protection against burnout. Although … main purpose of small group work is exchange …… of knowledge, social aspects should not be neglected because they will increase the motivation to continue with meetings[8].*

*The desire to be more competent and ''pride in performance'' are other key forces for change, while regulatory measures have little impact[8].*

*In addition, small group members have unique opportunities to discuss the way the individual patient experiences his or her illness through narratives, retold by the doctor[8].*

*The ways in which groups worked together in sessions seemed to be key to their success. …, group members sometimes seemed to strive to demonstrate their personal high standards of patient care. … the group challenged one such statement as unrealistic …[3].*

*Their expectations were mostly met as they found the time to ask questions and learn from both specialist and colleagues' opinions and knowledge. They found the time spent on clarifications, discussions and questions very useful[35].*

*Relevant factors identified in effective training initiatives include: the use of distributed practice techniques; the development of mentor-type relationships; the use of interactive learning and skills-based training, and the use of a format which enables doctors to discuss ongoing patients[58].*

**Supplemental material 8: literature based programme theory including illustrative quotes and complete list of supporting quotes**

*With the collaborative learning of residency training no longer available, clinicians often adopt idiosyncratic approaches when they encounter patient-care situations that cause them to question the limits of their own knowledge, … how to distinguish between their own knowledge limits and that of the medical canon—…, clinical uncertainty*[19].

*Social constructivist learning theorists, medical educators, and primary care researchers identify the problematic patient case as a powerful professional learning opportunity. Whether and how one decides to take on these problems in the "swampy lowlands" of practice become, according to Guest, decisions about "deliberate practice"*[19].

#### *Context mechanism outcome configuration 9: 'previous knowledge is activated'*

If participants exchange case stories and experiences while actively listening to each other in the presence of a skilled facilitator in a safe environment (C), then they will share their knowledge by telling their own relevant stories (O) because the process activates knowledge they already possess (M).

*...an important element is the focus on daily, clinical GP problems. In our study GPs preferred to talk about clinical problems and tests linked to these problems, rather than to discuss abstract phenomena like total test ordering volume or the ordering of specific tests*[20].

*The improvement strategy concentrated on 3 specific clinical topics (cardiovascular conditions, upper abdominal complaints, and lower abdominal complaints) and the tests used for these clinical problems, because it was believed that the physicians would prefer to discuss specific clinical topics rather than specific tests*[24].

*The use of a case-based format encourages activation of previous knowledge, allowing better retrieval of knowledge in the clinical setting………. particularly when it involves participation in small peer groups that foster trust, promote discussion of evidence relevant to real cases and provides feedback on performance*[10].

*11/2- to 2-hour discussion period follows, in which one or two of our GP learners will present a case from their practice on the topic*[6].

*Group members prepare cases from their office and present them in 3 to 4 minutes, to set the stage for discussion. …, we actively solicited group participation throughout the session. This encouragement was a major function of the facilitator early on*[6].

*Participants were encouraged to bring their own cases in relation to the topic. In this group, members were given tasks at the end of the meeting and fed back on these at the next meeting*[41].

*The theoretical basis for changing practice begins with the individual physician's experience of patient care*[10].

*PBSG modules are designed to engage family physicians "in learning activities that are self-directed and related to authentic practice problems.... The cases, linked with important information, are the keys to stimulating discussion around patient care issues*[10].

*The aim is not to solve the presented problems, rather the problems should act as a stimulus to encourage the group members to identify, discuss and address cases from their own experience too*[7].

*Virtually everyone participates in presenting a case, asking for advice or clarification, or describing their practice patterns*[6].

**Supplemental material 8: literature based programme theory including illustrative quotes and complete list of supporting quotes**

*During discussions at the level of relationships (case discussion), the exchange is more intense than in the exchange of pure facts; one's own behaviour is better analysed and suggestions for training in one's own practice came up [translated from German][18].*

*In each session, a colleague presented a difficult clinical case, which was discussed in the group according to a clearly structured manual, they sought solutions together and in the final phase, the group suggested a new treatment plan, which the presenting colleague had to try to implement in his practice [translated from German][18].*

*By dealing with actual clinical cases, real difficulties in the participants' everyday practice become the subject of discussion in the quality circle instead of constructed problems. In systematic reconstruction, participants make the experiences conscious, so that intuitively applied - implicit guidelines can be made explicit [translated from German][21].*

*Case discussions were by far the most popular agendas in groups[8].*

*Group work is built on sharing and improving ''collective'' knowledge and well-functioning groups provide this in an atmosphere of joy and curiosity[8].*

#### *Context mechanism outcome configuration 10: 'immediate relevance for the practice'*

Improved wording: If QCs use the technique of experience-based learning (C), then knowledge becomes more relevant to GPs (O), because it relates to their everyday work and is therefore of immediate use (M).

*...an important element is the focus on daily, clinical GP problems. In our study GPs preferred to talk about clinical problems and tests linked to these problems, rather than to discuss abstract phenomena like total test ordering volume or the ordering of specific tests[20].*

*The improvement strategy concentrated on 3 specific clinical topics (cardiovascular conditions, upper abdominal complaints, and lower abdominal complaints) and the tests used for these clinical problems, because it was believed that the physicians would prefer to discuss specific clinical topics rather than specific tests[24].*

*The use of a case-based format encourages activation of previous knowledge, allowing better retrieval of knowledge in the clinical setting…. particularly when it involves participation in small peer groups that foster trust, promote discussion of evidence relevant to real cases and provides feedback on performance[10].*

*11/2- to 2-hour discussion period follows, in which one or two of our GP learners will present a case from their practice on the topic[6].*

*Group members prepare cases from their office and present them in 3 to 4 minutes, to set the stage for discussion. …, we actively solicited group participation throughout the session. This encouragement was a major function of the facilitator early on[6].*

*Participants were encouraged to bring their own cases in relation to the topic. In this group, members were given tasks at the end of the meeting and fed back on these at the next meeting[41].*

*The theoretical basis for changing practice begins with the individual physician's experience of patient care[10].*

*PBSG modules are designed to engage family physicians "in learning activities that are self-directed and related to authentic practice problems.... The cases, linked with important information, are the keys to stimulating discussion around patient care issues[10].*

**Supplemental material 8: literature based programme theory including illustrative quotes and complete list of supporting quotes**

*The aim is not to solve the presented problems, rather the problems should act as a stimulus to encourage the group members to identify, discuss and address cases from their own experience too*[7].

*Virtually everyone participates in presenting a case, asking for advice or clarification, or describing their practice patterns*[6].

*During discussions at the level of relationships (case discussion), the exchange is more intense than in the exchange of pure facts; one's own behaviour is better analysed and suggestions for training in one's own practice came up [translated from German]*[18].

*In each session, a colleague presented a difficult clinical case, which was discussed in the group according to a clearly structured manual, they sought solutions together and in the final phase, the group suggested a new treatment plan, which the presenting colleague had to try to implement in his practice [translated from German]*[18].

*By dealing with actual clinical cases, real difficulties in the participants' everyday practice become the subject of discussion in the quality circle instead of constructed problems. In systematic reconstruction, participants make the experiences conscious, so that intuitively applied - implicit guidelines can be made explicit [translated from German]*[21].

*Case discussions were by far the most popular agendas in groups*[8].

*Group work is built on sharing and improving ''collective'' knowledge and well-functioning groups provide this in an atmosphere of joy and curiosity*[8].

#### *Context mechanism outcome configuration 11: 'cognitive dissonance'*

If participants discuss and reflect on their work processes (e.g., based on trustworthy data or personal experiences) during a professionally facilitated exchange of positive experiences or failures (C), then they discover knowledge gaps and identify learning needs and relevant topics (O) because their own attitudes and behaviours may differ from their peers', creating cognitive dissonance a negative emotional state triggered by conflicting perceptions that makes them reconsider their own way of working (M).

*The identification of suboptimal pharmacological treatments to be targeted in this study, was based on previous research and active reflection and discussions based on own clinical experience from general practice*[13].

*In the continuing medical education group setting, each participant was confronted with, and had to reflect on, the baseline report on their own prescription practice. We believe that this was a key component for obtaining improved prescription habits*[54].

*We consider the key element in our study to be "What happens to a general practitioner's prescribing behaviour when he or she reflects on his/her prescriptions?"*[54].

*Our intervention required general practitioners to expose their own antibiotic prescribing data in their continuing medical education group by using a structured pedagogical method, critically reflecting on the need for change together with an active listener*[54].

*Academic detailing involves educational outreach visits and incorporates external audit and supervision, and has a larger effect on prescribing than dissemination of educational materials, audit or feedback alone*[9].

*Peer group academic detailing was experienced as a suitable method to learn more about pharmacotherapy, though there were participants who argued that the scheme was time-consuming*[9].

#### Supplemental material 8: literature based programme theory including illustrative quotes and complete list of supporting quotes

*GPs' "hits" for inappropriate prescriptions in the elderly, or an unfavourable antibiotic prescription profile, was the starting point for group discussions at the second meeting. Tutors reported that GPs tried to justify and explain their practice*[9].

*Our results are in concordance with research that suggests that GPs may feel disappointment if their prescribing practice conflict with their ideals*[9].

*One important outcome for the GPs was an experience of being more reflective in decision-making about prescriptions*[9].

*The older … were silent, because they had a prescription profile … far from … recommended. The young … dominated the discussion, and they were much more familiar with the guidelines … … the old felt distress when disclosing their profiles ... have repeated their errors for … decades*[9].

*GPs were generally more embarrassed if they had hits they knew they should have avoided, such as prescribing flunitrazepam to elderly patients, compared to potentially harmful drug combinations that had not been highlighted in the recommendations*[9].

*The findings underscore that tutors have an important role in managing distress and contributing to an informal and relaxed atmosphere in peer academic detailing groups*[9].

*….and how to facilitate learning within a group setting*[40].

*Social interactions were used as an important motivator for change, as physicians learned how colleagues were handling test ordering problems and as they obtained information about the consequences of medical decision making in daily practice*[55].

*Personalized graphical feedback, including a comparison of each physician's own data with those of colleagues; dissemination of national, evidence-based guidelines, and regular meetings on quality improvement in small groups. The strategy focused on specific clinical problems and the diagnostic tests used for these problems*[55].

*The first was mutual personal feedback by peers, who worked in pairs at the start of the meeting. This was assumed to be a safe method of peer review*[20]

*A second important element is the fact that GPs are prepared to discuss personal, transparent data openly in a group of colleagues*[20].

*Compared with only disseminating comparative feedback reports to primary care physicians, the new strategy of involving peer interaction and social influence improved the physicians' test-ordering behavior. To be effective, feedback needs to be integrated in an interactive, educational environment*[24].

*90-minute standardized small-group quality improvement meeting about 2 weeks later at which one of the clinical problems was discussed based on the feedback reports and the guidelines … In these meetings social influence, which was an important vehicle to reach improvement on test ordering*[24].

*The second component was an interactive group education of national guidelines, to enable participants to relate their own and each other's test ordering behaviour with them*[20].

*The new strategy utilised peer influence among GPs, and gave GPs the opportunity to openly discuss their test ordering behaviour with colleagues*[20].

*They stated that this type of feedback definitely had added value, because comparison with colleagues made them more conscious of their own behaviour and motivated them to change. Their main criticism was the validity of the numbers of tests in the feedback and the absence of patient-related data*[20].

**Supplemental material 8: literature based programme theory including illustrative quotes and complete list of supporting quotes**

*Participants were shown the overall data on prescribing of antidepressants in the past year to illustrate that most anticholinergic antidepressants are prescribed to people aged over 60…. During the second visit a graph was provided showing personal performance*[25].

*…all doctors received a summary of their group's guidelines by mail, and two months after the intervention they received the results of the baseline measurement (see outcome variables) to reinforce the consensus reached*[2].

*Of the 40 GPs that reported having received individual feedback, 37 rated it as useful*[2].

*The purpose is to enable the transfer of evidence into practice through the use of facilitated small groups, using presented cases to encourage reflection on individual practice*[7].

*…it provides an opportunity to measure one's practice against that of one's peers. Direct, extended interaction with a local recognized...* [6].

*It is a relaxed, enjoyable evening in a friendly environment. …. This exchange allows for clarifications and redirections, leading to learning for both GP and expert. It is also an opportunity for the expert to learn of the tremendous competence that exists within GP practice*[6].

*Of greatest importance to GPs is the opportunity to measure their current practice patterns against that of their peers*[6].

*Individualized feedback with specific recommendations, especially when combined with education, generally have been more effective than single intervention*[59].

*One objective of the PBSG program is to encourage physician members to reflect on their individual practices and identify any gaps between current practice and the best available evidence. This is accomplished through discussion of real-life medical and patient problems in small groups of peers*[10].

*Through reflection, a gap between current practice and best practice is recognized. Distinguishing this gap presents an opportunity to identify learning objectives specific to the family practice setting*[10].

*Physicians who received feedback about personal prescribing or who used the PBSG process to discuss hypertension were more likely to change their prescribing than physicians in PBSG who reviewed a different condition. When feedback about personal prescribing was combined with the PBSG process, the effect … was even greater*[10].

*One of the key features of QCs is that working methods map the quality of care in one's own practice. First of all, this distinguishes QC work from further training in the classical style and second, it enables participants to identify real quality problems in their own practice [Translated from German]*[14].

*A systematic procedure …: data in the feedback report were studied …, reasons for variation were discussed, … prices of drugs, evidence underlying drug treatment was considered, typical patient cases from practice were analysed and finally objectives for improvement were formulated, and specific plans for improvement were made*[38].

*With regard to taking a practical assessment of their own actions, some participants feared that they would only be burdened with additional work, but on the other hand they were also very curious to see what we were doing [translated from German]*[34].

**Supplemental material 8: literature based programme theory including illustrative quotes and complete list of supporting quotes**

*A case-related approach offers the opportunity to confront learned normative expertise and concrete actual action in one's own practice......By comparing one's own perceptions and the viewpoints of other circle members, as well as by confronting assumed and real actions, e.g. by analysing video recordings, individual and collective defence strategies can become conscious. The deviations from one's own normative expertise and thus the problem of the implementation of existing theoretical knowledge into everyday practice become accessible for analysis [translated from German][21].*

*A basic problem of continuing medical education is the well-known mismatch between individual, existing specialist knowledge and putting this knowledge into everyday practice......Quality circles as a form of QI, which among other things also serves the purpose of continuing medical education, can improve this situation [translated from German][21].*

*On the basis of documentation of one's own activities in daily practice (e.g., in the form of index card evaluations, video recordings, EDP extracts, documents that one has created oneself, etc.), it is possible to learn from your own actions [translated from German][21].*

*They (the modules) are didactically structured in a way that allow comparisons of systematic processes with the actual procedures in practice. This stimulates the participants and the principle of cooperative learning can be realised.... The participants should identify, name and document deviations from their medical actions. This also includes checking whether documentation and data material is available [translated from German][16].*

*The aim of the project was to make doctors' own prescription behaviour transparent and to highlight problem areas [translated from German][37].*

*Presentation of these predefined guidelines was meant to encourage the participating doctors to assess their own performance and to foster discussion and refinement of the moderator-manuals[31]*

*GPs see QCs on pharmacotherapy as a sensible and useful measure for optimizing their own prescription methods. They regard part 1 of the prescription mirror as the most important instrument of the quality circle work (i.e.: the feedback of one's own, specially prepared prescription data with the possibility to compare oneself with colleagues of the project group as well as a GP control group without intervention) [translated from German][43].*

*Quality circles comprise a practice-base strategy to improve professional performance, which is based on meetings in small groups of health professionals, provision of evidence-based information, written feedback on professional performance and exchange of best practices in improving patient care[38].*

*The intervention comprised of quality circles of primary care physicians, including repeated feedback on prescribing patterns … nine small group sessions in which the feedback, guidelines on appropriate prescribing and exchange of best practices in changing performance were discussed…... The report included evidence-based information on prescribing in targeted conditions[38].*

*What sources of information were used? Still considered a "classic", the oral case presentation was by far the most frequently used source of information in 56.2% (=15,313 meetings). Other methods such as index cards, note sheets, data from electronic medical records were used significantly less frequently (20%) [Translated from German][4].*

*Following the training meeting, CQC members collected baseline data on patients from their practices using the CQC-form to ascertain how they currently diagnosed and treated osteoporosis…. These profiles, displayed graphically with brief text summaries, permitted anonymous comparisons of individual circle members' practices[45].*

**Supplemental material 8: literature based programme theory including illustrative quotes and complete list of supporting quotes**

*Physician profiles were displayed graphically with a brief text summary. The profiles permitted anonymous comparisons of individual circle member data with their peers in their circle and with all the participating physicians in the project*[50].

*Based on the questionnaire responses, physician profiles were generated that showed how individual physicians treated patients …. The profiles permitted anonymous comparisons of individual circle member data with their peers … and with all the participating physicians …. The physicians' profiles were than compared to the Osteoporosis Canada guidelines*[60].

*The PPQC process includes a combination of several elements (e.g., local networking, feedback, interdisciplinary continuing education) that facilitate changes in prescribing practice GPs*[28].

*The pharmacist compares individual prescribing habits with treatment recommendations (clinical guidelines) and with the most up-to-date information on the efficacy of the medication [translated from French]*[46].

*The GPs were asked to bring copies of the records for these patients to the meeting. During the meetings, the treatment of these specific patients was discussed, especially differences between what was prescribed according to the records and what was actually dispensed*[11].

*The combination of the written simulated cases with actual prescribing allows the GPs to reflect on their decisions as well as the background for these decisions, and is in line with suggestions to make drug utilization studies closer to the reality of practice*[11].

*.. their perception of a gap between their current knowledge and skills and those needed. … Cognitive feedback is feedback on the decision process, i.e., why or how a decision is made and not on the decision itself, i.e., which decision is taken (outcome)*[11].

*.. feedback regarding the written simulated case. ...feedback on actual decisions taken ..., the extent of use of the information factors ... and the agreement on decisions between individual members within the group*[11].

*Feedback was given on actual decisions… and factors taken into account when these decisions were made—so-called cognitive feedback—in our case, clinical judgment analysis (CJA)*[47].

*When looking at knowledge and attitudes, the largest improvements were indeed seen when the baseline performance was low*[26].

*… overview of the recommendations given in the guidelines. The major component … was to discuss individualized feedback on the decision process underlying treatment decisions, …. Using series of 18 case vignettes, factors triggering specific treatment decisions were identified …. The case vignettes were constructed to represent real patients ….*[15].

*Doctors may, during this active process, discover the consequences of new knowledge in relation to their own behaviour*[57].

*… to introduce independent information about polymedications, …. During these meetings, public health consultants also provided feedback information about prescribing patterns and cost. Quality circles (two in each area) met every 6 weeks and some GPs were trained and given documentation*[48].

*The current programme was multifaceted and included expert input, voluntary feedback, peer review, and specific recommendations for changes: all features generally associated with the successful implementation of changes in general practice*[48].

**Supplemental material 8: literature based programme theory including illustrative quotes and complete list of supporting quotes**

*As GPs had previously found its provision of diagnostic and therapeutic guidelines feasible and effective, the steering group .. prepared educational material …. In addition to prescribing behaviour, costs of drugs prescribed and the use of generics in primary care, … were addressed*[53].

*They supplied statistics on all drugs prescribed by GPs under contract and the costs involved, so that the intervention provided a repeated written feedback to participants on their personal prescribing behaviour*[53].

*Most surprising was the reaction to the presence of these texts (clinical evidence) during the educational sessions. The researcher observed that GPs seized on the books with gusto when the facilitator brought them into view*[3].

#### *Context mechanism outcome configuration 12: 'social learning'*

If the facilitator uses purposeful didactic techniques (e.g., brainstorming, contentious or consensus discussions, or role play) to keep the group active and to reward exploratory behaviour during reflection on the work process (C), then the group will create a learning environment that promotes knowledge exchange (O) because learning is a cognitive process in which participants observe and imitate their peers' behaviour to gain social approval (M).

*The identification of suboptimal practice is, however, only the first step for quality improvements. Several educational strategies have been used to improve doctors' clinical practice, but substantial effects are only rarely reported*[13].

*… found evidence that educational intervention consisting of passive dissemination of clinical practice guidelines had little or no effect on practice. This corresponds with later reports … More active strategies, like educational outreach visits and multifaceted interventions, are more effective, but require more resources*[39].

*The elements of the intervention are discussions within the peer group, collection of individual prescription data, audit based on individual feedback reports, as well as a one-day regional work-shop*[39].

*The participating GPs experienced the CME group meetings as an important arena for learning. They reported picking up good advice from others and learning practical alternatives …. GPs said their prescription data would not mirror all learning effects: 'The whole point is to reflect more, …… '*[9].

*Peer group academic detailing was experienced as a suitable method to learn more about pharmacotherapy, though there were participants who argued that the scheme was time-consuming*[9].

*The participating GPs experienced the CME group meetings as an important arena for learning. They reported picking up good advice from others and learning practical alternatives …. GPs said their prescription data would not mirror all learning effects: 'The whole point is to reflect more, …… '*[9].

*GPs said that the feedback on their prescription profile motivated them for reflection, learning, and change. Critical reflections on own strategies help change attitude and behaviour*[9].

*…. facilitated the discussion within the CME group, where each GP exposed their own prescription patterns as presented in his or her report and potentials for improvements were discussed within the group*[40].

*The systematic approach of the quality cycle was used reasonably well, although practices did have some difficulties in gathering data and evaluating progress in the improvement projects*[32].

*An intensive small group education and peer review programme, which combined various strategies, was proved to influence aspects of knowledge, skills, opinions, and the presence of equipment according to the guidelines but on its own no significant influence on the provided care*[30].

**Supplemental material 8: literature based programme theory including illustrative quotes and complete list of supporting quotes**

*It should be noted that a new skill is required for the recommended technique. It is possible that many midwives have not as yet learnt these skills. Small group CQI is not sufficient for the teaching of new techniques*[33].

*we found that small peer group CQI had a positive effect on changing clinical practice when no new skills had to be learnt, when the recommendations were considered to have more advantages than disadvantages, and when there was no 'ceiling effect' at baseline*[33].

*… of a strategy that combines a traditional feedback strategy with a multifaceted strategy, including feedback, dissemination of and group education on evidence-based guidelines, and small group quality improvement meetings in a local primary care physicians' group, using social influence as an important motivator for change*[61].

*A multifaceted strategy combining comparative feedback on tests ordered, group education on guidelines, and small group quality improvement meetings in a local GP group, with social influence as an important motivator for change, was expected to offer good prospects*[20].

*Compared with only disseminating comparative feedback reports to primary care physicians, the new strategy of involving peer interaction and social influence improved the physicians' test-ordering behavior. To be effective, feedback needs to be integrated in an interactive, educational environment*[24].

*At these meetings, test-ordering behavior and changes in routines were discussed, using social influence and peer influence as important motivators for change. Social influence from respected colleagues or opinion leaders seems to have a greater effect on practice routines than do traditional medical education activities …*[24].

*Many test-ordering problems that physicians encounter in everyday practice, such as demands for tests by patients and changing guidelines, can be discussed and may be solved in an open and respectful discussion among colleagues*[24].

*Our intervention—which included a group education meeting with a consensus procedure and communication skills training*[2].

*...various strategies for implementing the guidelines were used: lectures, role playing, skills training, peer review of performance, group consensus discussions, and problem solving of hypothetical situations involving patients. The group education and review was done in two small groups …….) and was supervised by an experienced GP*[62].

*An intensive small group education and peer review programme, which combined various strategies, was proved to influence aspects of knowledge, skills, opinions, and the presence of equipment according to the guidelines*[62].

*Either the facilitator or the expert is asked to recommend one or two relevant articles to follow up the discussion. The expert has frequently selected an article in advance, from knowledge of frequently asked questions in prior learning environments*[6].

*The group selects topics, directs the agenda, points out inappropriate comments or inappropriate practices in a constructive manner, and also leads group members back on topic. The conversation is free-flowing and highly interactive*[6].

*The group encourages other points of view to establish practice norms. This allows individual GPs to see where they may deviate from usual standards of care*[6].

*What have you gained from participating in this practice-based small group learning project – learning from colleagues: the group discussion allowed us to share our experience and management of various problems*[41].

**Supplemental material 8: literature based programme theory including illustrative quotes and complete list of supporting quotes**

*Group discussion allows for sharing of experiences and of thoughts about strategies for implementing practice changes and about overcoming anticipated barriers (Armson et al., 2007)[10].*

*Learning from and with colleagues is an important source of both new information and strategies for applying that information to practice[10].*

*Most GPs and PNs valued learning together. Several GPs from Group 1 said that they were consistently satisfied with the learning that took place in their group[5].*

*I think there is a mutual keenness to learn from each other[5].*

*A ''mutual keenness'' to learn from and about each other emerges as a crucial ingredient for learners to feel that their learning needs were being met given the multi-professional context[5].*

*'Learning from colleagues' (three comments) 'Has been very constructive and helpful[42].*

*….to work through cases together and massively furthered my learning.' 'Really useful (secondly) discussion with colleagues/peers regarding management of conditions in real practice[42].*

*This should not be done schematically according to a fixed schedule, but rather with the help of various methods that reflect the reality of everyday practice (e.g., case discussions, file card analysis, documentation with a study character, video, etc.). They can also be used in parallel [translated from German][34].*

*At the same time, our concept leaves room for case-related and problem-oriented learning using our own patient examples from practice [translated from German][36].*

*They (the modules) are didactically structured in a way that allows comparison of theoretical approaches with the actual procedure in practice and so the principle of cooperative learning can be realised [translated from German][16].*

*The principle of cooperative learning in the quality circle (everyone learns from everyone else) leads to increased flexibility and more pleasure in practice [translated from German][16].*

*Almost 73 % of the participants thus confirm that the intercollegial exchange in the Pharmacotherapy Quality Circle - as in all other quality circles (10-12) - can be regarded as one of the fundamental mechanisms of action [translated from German][43].*

*They particularly appreciate the opportunity to compare their own prescribing behaviour with that of their colleagues and to discuss it in the familiar setting of a small group. In addition to this intrinsic principle of quality circles of intercollegial, equal and non-hierarchical exchange in a familiar group (of so-called "peers"), the feedback of one's own prescription data with the possibility of comparing oneself with other colleagues and to measuring one's own progress in the context of a before-and-after comparison (evaluation) contributes significantly to the success of the project [translated from German][43].*

*Especially the expectation of a successful collegial exchange of experiences, which was most frequently mentioned at the beginning of the project, seems to have been fulfilled: The vast majority of participants emphasised that they had received helpful tips from colleagues who had helped them to implement changes [translated from German][44].*

*Regular ... reflection on common practice with other colleagues. … individual feedback, …. discussed in the group under the guidance of a moderator … benchmark activities… The core element of these circles is the conjoint discussion of evidence-… and management of patients on the basis of prescribing data….[63].*

**Supplemental material 8: literature based programme theory including illustrative quotes and complete list of supporting quotes**

*… impact of physicians' views on the use of performance feedback, indicators and price comparisons. This set of views reflects both a willingness to reflect critically on one's professional performance and a positive attitude regarding the ideal of evidence-based medicine*[38].

*… an important component of the improvement strategy is an individual learning activity: reading and reflecting on the written feedback reports. This is consistent with insights from educational research, which showed that …. learning activity is an important predictor of the effectiveness of any educational programme for professionals*[38].

*It involved practice audits, feedback on performance by peers …, interactive discussion of evidence, small-group educational workshops led by … facilitators and supported by local osteoporosis specialists, diagnosis and treatment reminders (CQC-forms), and making personal plans for improving clinical management of osteoporosis in accordance with the OC 2002 guidelines*[45].

*The educational intervention consisted of eight key components: 1) audit and feedback, …; 2) interactive small group discussions …; 3) use of opinion leaders …; 4) reminders, …; 5) multi-professional collaboration and community building …*[50].

*Our educational intervention consisted of eight key components and consisted of 1) audit and feedback, 2) interactive small group discussions 3) use of opinion leaders, 4) remainders, 5) multiprofessional collaboration with osteoporosis specialists, 6) nominal financial reimbursement to circle members, 7) patient medicated interventions 8) and educational material*[60].

*The key elements are local networking; feedback of comparative and detailed data regarding costs, drug choice, and volume of medical prescriptions; as well as interdisciplinary continuing education adapted to primary care needs*[28].

*(GPs)... depend less on factual knowledge than on their capacity to reflect in action, to be in control of ... learning process, to be motivated, and to perceive meaningfulness; ... it is beneficial ... when the social climate is supportive …*[11].

*The intervention comprised several elements, … the provision of individual feedback on the series of simulated cases … and on actual prescribing, use of outreach visits, use of peer group discussions, and use of existing guidelines. …, the sessions were especially tailored for each group*[11].

*Cognitive feedback is feedback on the decision process, i.e., why or how a decision is made and not on the decision itself, i.e., which decision is taken (outcome)*[11].

*The combination of the written simulated cases with actual prescribing allows the GPs to reflect on their decisions as well as the background for these decisions, and is in line with suggestions to make drug utilization studies closer to the reality of practice*[11].

*For UTI the usually high use of the non-recommended drugs was stressed and was related to the ... high use in the simulated cases and the cues that triggered these decisions… the identity of the individual doctor was disclosed at the request of the participating GPs*[11].

*Learning methods that have proven effective include interactive and problem-solving exercises combined with feedback on performance. Combined strategies that deal with different types of barriers seem to be more effective than single separate strategies*[26].

*Once the problem is acknowledged, one must learn and understand what caused the problem and how it can be solved. For this, elucidating and discussing the decision process underlying treatment decisions may be useful. To accept new information or practice recommendations the credibility of the source is of importance*[15].

**Supplemental material 8: literature based programme theory including illustrative quotes and complete list of supporting quotes**

*Problem based learning, …, places the emphasis on the learner's own initiative to discover problems and how to improve. By discussion in peer review groups the individual doctor's self-efficacy, defined as one's ability to organise and execute a course of action required to produce given results, is substantially increased*[57].

*Although academic knowledge is important, the fundament for professional development is reflection of one's own practice or, as Schön stated, ''reflection-in-action''. Until now, these aspects of learning have been mostly neglected, …*[8].

*The strengths of small CME groups are principally that learning is self-directed and based on relevant problems and ''reflection-on-action'', a pedagogic prerequisite for effective learning*[8].

*…, hardly any participants failed to contribute to the group discussions. … they thought that for a meaningful comparison of prescription costs such data must be correlated with morbidity and disorder distribution among patients. Every opportunity was taken to discuss various clinical aspects of patient management and pharmacotherapy*[53].

*…, CME should involve the learner actively and as we know from the protocols that there were hardly any participants who did not contribute to the discussions, we can say that our qualitative data support the general notion that quality circles are an appropriate CME format for practising physicians*[53].

*A defensive attitude of the GP, for instance, been linked to overprescribing, Quality improvement requires a reflective attitude of one's own knowledge and performance*[64].

*There is, however, adequate evidence that merely distributing a guideline without any additional intervention does not have an effect on prescribing behaviour*[64].

*Participants and facilitators saw the strength of the small groups as facilitating the learning of practical skills (e.g. through the use of role play*[35].

*Discussion is based on evidence-based topic notes prepared for each leader as well as individual prescribing and laboratory data related to the topic that is provided to each GP. Although the education groups cover all aspects of clinical practice…* [27].

#### *Context mechanism outcome configuration 13: 'interdependence between health insurance companies and GPs'*

If physician network organisations require continuous QC activities (C), then QCs will negotiate priorities and design creative solutions (O) because the tension between autonomy and obligation spurs the group to act and negotiate together to reach a common goal (M).

*The physicians in the Rhine-Main network of physicians committed themselves to participating in QCs when they joined the contract. In QCs, they discuss prescription patterns for specific clinical situations and adapt (guidelines) to local conditions [translated from German]*[22].

*The participation of German GPs in QCs is mandatory in order to be part of government-funded disease management programmes (DMPs) or to be part of pilot projects with health insurance funds*[23].

*The principle of 'quality circles' is now also used for clearly defined quality promotion purposes: The Associations of Statutory Health Insurance Physicians (KVs) of Hesse, Saxony-Anhalt and Lower Saxony started off with structured QC programmes to demand rational pharmacotherapy. In the meantime, QCs have become a requirement in numerous contracts (for disease management, family doctor-centred care, etc.) [translated from German]*[14].

**Supplemental material 8: literature based programme theory including illustrative quotes and complete list of supporting quotes**

*The QCPs were designed as a measure of quality assurance in pharmacotherapy and, as doctors were expected to encounter various problems in educating their patients in the use of generics, to offer them a forum for discussing these with their peers[53].*

*…. Specifically, we suggest that centrally organized experts make the strategic decisions about best practices based on evidence but local site staff members make tactical decisions about how best to implement the plan based on what fits local circumstances, needs, and cultures[12].*

*… interrelationships that exist among a particular organization's technologies, tasks, goals, stakeholder characteristics, and environment …. Participation provides one of the best methods for obtaining valuable information about .. local conditions[12]*

#### *Context mechanism outcome configuration 14: 'threat to professional autonomy'*

If GPs feel that the QC programme is only a top-down managerial intervention to reduce costs (C), then they will not be motivated and will not participate (O) because they feel unsafe and think they lack autonomy in their clinical role (M).

*In general, efforts are being made to improve practice performance by developing guidelines. Guidelines are intended to help general practitioners to tailor the care of individual patients to generally accepted scientific findings. However, guidelines are not sufficiently implemented. The reason for this is probably the lack of practicability and low relevance of the guidelines for family doctors. In addition, they give general practitioners too little room for their own medical decisions [translated from German][65].*

*… much pressure about their prescribing budgets that they participated … as an attempt to appease their prescribing adviser. This resulted in poor attendance and a reluctance to participate …. This defeated the notion of reaching and establishing a consensus that was 'owned' by the practice as a whole[3].*

*…., it emerged that it could be difficult for GPs to match top-down initiatives with everyday practice. The difficulty, which is another form of pressure, was expressed well by one GP who explained in the interview[3].*

*The structure of each session demanded a firm commitment ... to a common management strategy for …, we found that GPs were reluctant to do this. This reluctance appeared to arise from a sense of threat to their perceived need for clinical autonomy —... [3].*

*The concept of clinical autonomy is highly valued and it has been argued that in British general practice, prescribing is the principal battleground on which the cause of clinical autonomy is being defended[3].*

*An understanding of what GPs mean by clinical autonomy and how it affects their ability to reach explicit consensus on clinical management decisions is crucial if practice prescribing is to become more cost-effective. Many GPs perceive guidance on cost-effectiveness … as an intrusion on their professional independence[3].*

*… GPs and facilitators pointed to the difficulty of reaching consensus on a best buy, ……. Some found the term 'off-putting' because of its financial connotations. This suggests that some GPs may feel that their management decisions should be based on wider considerations than those of cost-effectiveness[3].*

*GPs were also unlikely to take part if they felt that the sessions would make them feel unsafe or if they felt that the sessions were yet another 'top-down' managerial intervention, where the main intention was to reduce prescribing costs[3].*

*The majority of respondents in both regions expected to benefit from participation in QCs, but they were unwilling to accept the risk that QI could be misused for control or cost reduction [translated from German][1].*

**Supplemental material 8: literature based programme theory including illustrative quotes and complete list of supporting quotes**

*In the discussion with facilitators, the QC participants' claim to be able to work in a self-determined and independent manner became apparent. For this reason, the National Association of Statutory Health Insurance Physicians (KBV) and the Associations of Statutory Health Insurance Physicians (KVs) have laid down the thematic and methodological autonomy of the circles as indispensable in their guidelines for quality circle work and have committed themselves to supporting them [translated from German][4].*

*Physicians have raised similar concerns about threats to the autonomy of their profession ... It is within this context, a time of declining perceived autonomy for individual physicians, that we compare the participatory local and central expert approaches to QI[12].*

#### *Context mechanism outcome configuration 15: 'interdependence among group members'*

If participants maintain a learning environment based on trust that promotes knowledge exchange, assisted by facilitators who use professional techniques (e.g., contentious discussion, reaching consensus, and role play), (C), then participants will adapt and generate new knowledge for local use (O) because they see themselves as similar, and so act and negotiate cooperatively to achieve a common goal (M).

*… that combining information from a peer detailer with reflection on one's own need for change together with trusted colleagues would improve prescribing patterns[54].*

*Balancing interests and concerns is an essential aspect of GPs' work[9].*

*The physicians discussed their feedback data, and if it appeared that a physician clearly ordered fewer tests than his/her colleagues, he/she made plans for ordering more tests[61].*

*interactive group education in which national guidelines were related to the individual physician's actual test-ordering behavior and an effort to reach a group consensus on the optimal test-ordering behaviour[24].*

*The personal interaction and mutual influence between colleagues implicitly resulted in an individual or group contract[24].*

*Psychological research into group behaviour has produced an inventory of factors that influence conformity with group standards. Unanimity provides more pressure to conform, while privacy makes it easier not to[25].*

*In presenting the evidence we used relative and absolute effects of antibiotics by means of the numbers needed to treat and the numbers needed to treat to harm. This discussion resulted in group consensus about indication and first choice antibiotics per disease[2].*

*…a learner-directed agenda of topics, presentation of information by trusted peers or local experts, and opportunity for practice and feedback. If the information comes from several sources—…—the perception of need for and the durability of change are enhanced[6].*

*Interactive approaches, however, can be effective, particularly when they involve participation in small peer groups that foster trust, promote discussion of evidence relevant to real cases, provide feedback on performance, and offer opportunities for practising newly learned skills[10].*

*It is known that small groups can encourage active participation and deep learning as well as learning of group skills and the ability to express new ideas[7].*

*The acquisition of new knowledge, skills, and approaches to bridge this gap follows. Often, however, access to new information alone is not sufficient. Reflection and discussion are necessary to help physicians 1) identify areas where current practice requires change and 2) develop strategies to integrate this new approach[10].*

**Supplemental material 8: literature based programme theory including illustrative quotes and complete list of supporting quotes**

*There was widespread agreement that the principal requisites for a good facilitator were experience and competence in small-group skills. One facilitator identified another skill: 'You've got to be able to hold the tension between comforting and challenging*[7].

*The decentralised approach at the local, internal level consists of collecting available knowledge from the everyday practice of the medical participants and formulating a workable consensus from this. The advantage of this method is that the physicians are actively involved in this process and are more motivated to implement the developed guidelines. In addition, this results in a stronger commitment and acceptance by the participants [translated from German]*[16].

*The programme for the meetings was based on principles of quality improvement, which implied that a systematic procedure was followed: themes were selected, objectives were formulated, plans for improvement were made and implemented and changes were evaluated*[29].

*In more than 90% of the meetings, new health care aspects could be identified according to the facilitators' assessment [translated from German]*[14].

*Facilitators ensure that participants not only focused on a specific topic, but also focus on their own actions in their own practices and that they identify blind spots in their daily work. Approximately 90% of the methods used are certified as having been able to reveal previously unknown aspects of care [translated from German]*[14].

*Discussions concerning the progress made by incorporating strategies identified in the prior phases of the project were shared among the group. Based on the major findings from the profiles, members discussed additional measures that should be implemented in their practices to increase alignment with the 2002 guidelines*[50].

*An analysis of prescription attitudes in comparison with scientific and economic data and the search for alternatives in the drug market is then run by each PPQC to build its own consensus. An annual assessment is conducted for facilitating the continuing improvement of the process*[28].

*.. over time those GPs who, at first, were reluctant to prescribe generics changed their attitude, …. After 2 years of QCP participation ..., GPs confirmed that the prescribing of generics, where appropriate, had for them become common practice and that their efforts and the various discussions… had helped*[53].

*The reluctance of GPs to appear in agreement with one another does not mean that discussions are pointless or ineffective. For example, as we found in an earlier study, the process of sharing different management strategies for a particular clinical problem may result in marked changes in prescribing behaviour*[3].

#### *Context mechanism outcome configuration 16: 'identifying and removing barriers to change'*

If participants, supported by skilled facilitators, address barriers to change (C), then they are more likely to implement the innovation (O) because participants help each other develop strategies to identify and overcome these barriers (M).

*Therefore, it is recommended to address potential barriers to change when tailoring an intervention targeting change in medical performance*[39].

*it appears to be essential that throughout the implementation personal obstacles are addressed*[30].

*The implementation of new knowledge is facilitated by expressing and discussing how to overcome obstacles to its acceptance*[25].

**Supplemental material 8: literature based programme theory including illustrative quotes and complete list of supporting quotes**

*The barrier most often mentioned for changing the CHF treatment was related to perceived difficulties with changing treatment initiated by a specialist[2].*

*Cranney also identified some barriers to translating evidence into practice including: doubts about the applicability of data to particular patients, against attitudes and the absence of an educational mentor[7]. [Trustworthy data]*

*.. study feedback to individual doctors ... ... The recommendations needed to be reformulated to enable a quality assessment of patient treatment to be judged from prescription feedback. Such quality criteria were developed during group discussions between doctors participating in the study[57]. [Trustworthy data]*

*Within the group, members endeavour to identify specific barriers to these practice changes and to formulate implementation strategies to facilitate desired changes[10].*

*The acquisition of new knowledge, skills, and approaches to bridge this gap follows. Often, however, access to new information alone is not sufficient. Reflection and discussion are necessary to help physicians 1) identify areas where current practice requires change and 2) develop strategies to integrate this new[10].*

*Commonality and differences between local practices. Some participants commented that listening to ''how peers work'' was a benefit: Finding out what everybody is doing locally . . . it makes you think 'would that be better?'[5].*

*… an educational workshop, and facilitators led small group discussions that identify barriers to the management of osteoporosis and strategies to improve patient care, family physicians demonstrated greater odds of administering osteoporosis therapy appropriately over a two-year period[60].*

*Back at the practice, the difficulty is to apply the consensus reached in the group, while considering the particular situation of each patient[46].*

*Barriers within doctors relate to competence, motivation and attitudes, and personal characteristics such as learning style, whereas barriers within practices exist as doctors do not work entirely independently[26].*

*These results make clear that, although in the educational program a lot of attention was paid to overcome barriers within GPs, barriers within practice setting may not have been sufficiently addressed, preventing the correct implementation of the recommendations concerning asthma maintenance treatment in practice[15].*

*Once a doctor has accepted a new practice and has the intention to change, there still may be several barriers within the practice setting that prevent the actual implementation in practice. Discussing problems encountered in everyday practice may help to overcome such barriers to implementation[15].*

#### ***Context mechanism outcome configuration 17: 'need for competence, autonomy and relatedness'***

If participants create new knowledge and plan an implementation strategy (C), then they feel satisfaction, responsibility and stewardship (O) because this fulfils their need for competence (being able to achieve specific objectives) (M), autonomy (a feeling of being in control of their own behaviour) (M), and relatedness (a sense of connection to a larger group) (M).

**Supplemental material 8: literature based programme theory including illustrative quotes and complete list of supporting quotes**

*The decentralised approach at a local, internal level includes participants gathering experience from daily practice and formulating a feasible consensus solution. The advantage of this method is that GPs are actively involved in this process and therefore motivated to implement the (newly) developed guidelines. In addition, participants involved will be more likely to accept (new knowledge) and feel committed to implement it [translated from German][16].*

*Potential advantages of the local approach: it promotes buy-in, maximizes fit to local culture and circumstances, maximizes the ability to work out the details associated with implementation, and produces a highly rewarding experience[12].*

*… new knowledge is … facilitated through the "... working with it, discussing it, and connecting it with what is … known…. because physicians … generate … 1 question for every 2 patients … the opportunity to explore these questions in …groups can stimulate … ideas for future change[10].*

*… working on projects… is … of great advantage. Everyone is involved ...has to prepare something for the next meeting. The structure of the quality cycle committed us to make all steps …. You don't cling to ideas but …come to changes. Evaluation … is … important ….[30].*

*A higher appreciation of the quality of the group discussion led to more effect of the intervention on the treatment of asthma exacerbations and on the duration of treatment prescribed for uncomplicated urinary tract infections[15].*

*The quality of the group discussion as evaluated by the participants seems to be an important predictor of successful educational group meetings[15].*

*When studying how physicians learn and change their medical practice, disposing, enabling, and forcing factors can be identified. These are a mix of professional factors, such as the desire for competence, social factors such as working climate, and personal factors such as curiosity[11].*

*International and national guidelines are more difficult to implement than local or internally developed guidelines[26].*

*Only in The Netherlands, national guidelines were developed by GPs and intended primarily for their use. This guideline initiative has been quite successful and highly accepted, because it is initiated and "owned" by the GPs themselves[26].*

*… In addition to the pragmatic benefits of the local approach, participants also mentioned one psychological advantage: intrinsic reward. …the local approach might be rewarding because it promotes team camaraderie…. "It [the local approach] is more creative and it's fun ... I enjoyed it"[12].*

*Not surprisingly, they personally relished the level of participation that the local approach affords. It is possible that … high level of enthusiasm permeated the entire team. In fact, every person on this team reported enjoying the opportunity to participate at a high level on this project[12].*

*…potential advantages of the local approach: it promotes buy-in, maximizes fit to local culture and circumstances, maximizes the ability to work out the details associated with implementation, and produces a highly rewarding experience[12].*

*Whereas the effectiveness of many PBLI methods is unknown, social interaction, a key element in some PBLI approaches, appears to increase physician satisfaction with learning and improve certain practice and patient outcomes[19].*

#### *Context mechanism outcome configuration 18: 'intention to change'*

**Supplemental material 8: literature based programme theory including illustrative quotes and complete list of supporting quotes**

If participants publicly announce their intention to change (C), then they are more likely to implement the change (O) because they and others in the group both think it is a good idea and believe they can carry it through (M).

*It is crucial to the model that practice teams formulate goals for improvement and attempt to achieve these goals in small scale*[32].

*The third was the development of individual and group plans for change, to stimulate GPs to really put their plans into daily practice*[20].

*An example of such an individual commitment was, 'I will order fewer haemoglobin tests, because I realise that this test does not give much information in patients with vague complaints'*[20].

*Plans at group level were also made, e.g., the plan to use the …brochure to inform patients …, or … to follow the national guideline on delaying testing in patients with vague complaints. All results show that the quality circles were an essential element in the improvement strategy*[20].

*The strategy gives physicians an opportunity to discuss their test-ordering performance with colleagues on the basis of actual performance data, making the participants feel more committed to the agreements*[24].

*Groups can be more effective in accomplishing tasks… and publicly announcing behavioural changes results in more commitment than privately announced change*[25].

*In most groups, there had been a discussion of the optimal treatment, as well as of barriers to change treatment in line with the recommendations of the guidelines. The idea was that by sharing experiences and learning from peers, possible solutions to perceived barriers might be offered*[2].

*…a structured tool for promoting reflection on the topic discussed at the group meeting and for identifying plans for practice change. The commitment to change section of the log sheet appears*[10].

*… participants stated that they had applied some learning to their practice. They reported a general increase in awareness of conditions and also confidence in treating them*[7].

*I was surprised to see how willing people were to reflect on their own behaviour and practice... and constantly make comments like: "Well, did I really do that? I surely have to pull myself together". Very strong will, apparently, to make changes*[9].

*The third was the development of individual and group plans for change, to stimulate GPs to really put their plans into daily practice*[61].

*Groups can be more effective in accomplishing tasks, and publicly announcing behavioural changes results in more commitment than private change*[25].

*…. draws on 'the theory of planned behaviour' and other studies that have identified the pre-requisites of successful behaviour change in general practice reviewed by Veninga et al.2000*[3].

*Discussions in the QCs are often lively and then lead to the determination of a consensus that everyone is committed to implementing in the best possible way [translated from French]*[46].

*The discussions within PPQCs are often lively and end in the determination of a common consensus that everyone makes a commitment to apply to the best of his or her ability*[28].

*Theories of adult learning stress the importance of motivation; the doctors must see the need and be willing to change their behavior to increase their professional competence*[26].

**Supplemental material 8: literature based programme theory including illustrative quotes and complete list of supporting quotes**

#### *Context mechanism outcome configuration 19: 'testing new knowledge'*

If participants validate and test new knowledge in a QC, moderated by a skilled facilitator in a safe environment (C), then they feel confident putting that knowledge to use in everyday practice (O) because they have had the opportunity to practise and familiarise themselves with the innovation (M).

*Interactive approaches, however, can be effective, particularly when they involve participation in small peer groups that foster trust, promote discussion of evidence relevant to real cases, provide feedback on performance, and offer opportunities for practising newly learned skills*[10].

*Understanding application of new knowledge. The discussions helped members to consider translating evidence into practice: Sometimes you can read about things but are unable to see how to put it into practice and I feel PBSGL enables you to think how you can do that*[5].

*Innovative solutions to clinical problems can be shared, and nonstandard methods are highlighted in a nonthreatening way*[6].

*…, in some situations, evidence may not exist or local experts may disagree with the evidence. The facilitator can help by reinforcing the tenets of evidence-based medicine, by selecting methodologically sound overviews …, and by asking the expert to address any evidence that exists for the recommendations made*[6].

*This means, for example, that the group is currently working on a new topic, while, analogous to steps d and e, checks are made whether changes have taken place in the doctor's actions (or in the actions of the entire practice team) with regard to the previous topic [translated from German]*[34].

*Next, they examined empirical evidence concerning the validity of these solutions. To facilitate this process, teams had access to the large resource library that the research team had assembled*[12].

#### *Context mechanism outcome configuration 20: 'gaining confidence in an innovation'*

If the group repeatedly practices implementing and adjusting to an innovation (C), then they trust their own competence and turn the innovation into a habit (O) because successful outcomes increase their confidence in their abilities (M).

*A cyclic process … is used which leads project teams through the improvement projects. This means that after having chosen a subject that requires attention, the team sets specific targets for the project, analyses the actual performance on the subject, makes and introduces plans for change, and evaluates progress*[32].

*One meeting may not be enough to actually change treatment, although that is the usual procedure in the peer review groups. Behavioural theories stress the importance of repetition, especially for changing routine behaviour*[2].

*Six months after the intervention, general practitioners again received feedback on their prescribing behaviour, based on insurance claims data comparing the period after the intervention (March to May 2001) with the same period before the intervention (March to May 2000)*[2].

*In general, GPs were excited to find in the second year that they had indeed changed in accordance with their plans, and they were then usually more motivated to implement further changes*[61].

*The intervention comprised repeated feedback on prescribing routines and an intensive programme of educational small group sessions, as described by Bahrs et al. (2001)*[29].

**Supplemental material 8: literature based programme theory including illustrative quotes and complete list of supporting quotes**

*Suitable data illustrate everyday practice. Participants formulate and discuss possibilities for improvements within the collegial framework of the quality circle and implement these in a further step in their own practice. Renewed data collection then allows them to observe effects of the implemented measures and gain confidence. The results are input for a new discussion in the quality circle [translated from German][44].*

*The analysis that is carried out each year secures change, as they give the pharmacist the means to maintain motivation: each doctor receives detailed feedback on their successes and the progress still to be made in relation to a control group (doctors working without particular collaboration with pharmacists) and in relation to the good results of other colleagues [translated from French][46].*

*The constant feedback on progress achieved and the further possible improvements are other success factors[28].*

*The evaluations of the GPs' prescriptions are performed every year to provide concrete feedback and a source of motivation … to change prescription attitudes[28].*

*Doctors who were accustomed to discussing their prescribing in peer groups changed their behavior more as a result of such (iterating) peer group meetings than doctors who are not used to this approach[26].*

*In contrast to our pragmatic study, the interventions in most trials consist of multiple sessions on the same topic supervised by a researcher or an expert, a situation usually very different from real life[64].*

*These results demonstrate the need to look at repeating/reinforcing messages at 12–24-month intervals[27].*

#### *Context mechanism outcome configuration 21: 'repetition priming and automaticity'*

If participants build a steady group and practice using QI tools (C), then they will successfully implement new knowledge into everyday practice (O) because responses improve with repetition: 'practice makes perfect' (M).

*This favours change: Having regular practice meetings on quality improvement with all staff [30].*

*Successful projects might not only positively reinforce the introduction of CQI, but could also bring about a positive attitude to the other aspects of systematic and continuous quality improvement[30].*

*Regular meetings with the practice team was selected as a topic for improvement by several of the practices[51].*

*Finally, for the same reason we were unable to assess possible learning effects, which could mean that quality activities may become less time-consuming over time, even if the approach is directed to other clinical problems[61].*

*This schedule was repeated a year later, using the same three clinical problems, to assess whether a GP or GP group had implemented the plans for change and to initiate further improvements. This iterative aspect was another important feature of the strategy[20].*

*In general, GPs were excited to find in the second year that they had indeed changed in accordance with their plans, and they were then usually more motivated to implement further changes[20].*

*Our strategy also seems worthwhile because small-group quality improvement meetings can help build a local practice group focusing on quality improvement[24].*

*However, other studies have shown that repeated interventions are needed for sustained behavioural changes[25].*

**Supplemental material 8: literature based programme theory including illustrative quotes and complete list of supporting quotes**

*…one meeting may not be enough to actually change treatment, although that is the usual procedure in the peer review groups. Behavioural theories stress the importance of repetition, especially for changing routine behaviour*[2].

*The benefit from participation depended significantly on the frequency of the meetings. Real improvements to performance in daily care can only occur if there is an ongoing and regular quality circle process*[31].

*The benefit from participation depended significantly on the frequency of the meetings. Successful projects might not only positively reinforce the introduction of continuous QI, but could also bring about a positive attitude to the other aspects of systematic and continuous quality improvement*[30].

*The intervention comprised repeated feedback on prescribing routines and an intensive programme of educational small group sessions, as described by Bahrs et al. (2001)*[29].

*Assuming a straightforward dose–response relationship, it was expected that the groups were most effective when physicians participated in most sessions. Stronger effects were also expected, if the groups comprised of physicians who had more experience with learning in small peer groups …*[38].

*The quality circles (N=1,241) documented an average of 22 meetings (mean value: 21.96) (range: by definition min. 4, max. 127 meetings [translated from German]*[14].

*The higher the attendance rate and the more experienced the GPs in a group were, the shorter the courses were prescribed for UTI after the intervention*[15].

*In principle, material learnt in brief training workshops decays quickly over time, whereas repetition on many occasions ensures greater retention*[58].

*Practitioners develop expertise when they move from their comfort zones to examine problems "at the upper limit of the complexity they can handle;" they learn, and iteratively gain mastery through cycles of reflecting on practice, obtaining feedback, and adjusting performance*[19].
