## Supplemental material 9 for "How and why do Quality Circles work for General Practitioners - a realist approach"

**Supplemental material 9 revised programme theory including illustrative quotes and complete list of supporting quotes**

| |
|---|
| CMO configuration 1: **'participants know what to expect'**<br><br>If the introductory workshop teaches the principles of QI in PHC and illustrates how QCs work (C), then potential members may be more willing to join QCs (O) because they know what to expect and feel that they can meet expectations(M).<br><br>*So, I think that everything should be well organised and planned, and that you really have to understand what it's all about – as far as you can understand it without having gone through the process at least once [translated from Swiss German] (2).* |
| CMO configuration 2: **'need for autonomy and obligation'**<br><br>If the administration at the national level or at the level of health insurance companies entrusts GPs with QI and autonomy (so they can decide how to implement it) (C), then GPs might participate in QCs (O) because they feel they can take on the responsibility and make a difference (M).<br><br>*Yes, of course, these are specific drivers [to meet], and after all it [being member of a physician network] is like a commitment to participate in it [QC]… at the same as you may fear limitation in your professional autonomy etc… but there are rules and, if everybody sticks to these rules, it [QC] will work [translated from Swiss German] (3).* |
| <u>New</u> CMO configuration at the organisational level: **'feeling they have a say'**<br><br>If an organisation, (e.g. a physician network organisation) has a decentralised policy that encourages the use of local knowledge (C), then the QC takes on tasks (O) because members feel that they have a say in QI in their practice (M).<br><br>*I was asking myself what autonomy is and what it actually means ... so decentralised organisation means accepting local decisions and having a flexible administration. So not simply stubborn and rigid, but adaptable to local decisions, which are then taken by the QC, for example … [translated from Swiss German] (2).*<br><br>*So, if I want to promote and do QCs, then it's in the nature of the QC that different groups come up with different solutions. And you have to be able to live with them and you also have to be able to endorse them as an organisation, otherwise the QC instrument wouldn't make sense [translated from Swiss German] (1).* |
| CMO configuration 3: **'sharing similar needs'**<br><br>If the administration at the organisational level of QCs provides support (i.e. in training facilitators, data gathering, provision of evidence-based information), and the administration protects time and space and offers CME points and small financial incentives to QC participants (C), then the latter will meet in groups to exchange ideas (O) because GPs prefer learning in QCs (M); support generates positive expectations among participants (M), and GPs think QC meetings with their peers will be useful (M).<br><br>*Yes, this kind of network allows high organisational and professional autonomy ... and this is a good start for QCs [translated from Swiss German] (2).*<br><br>*For me, protected time is more than just the aspect of time; it relates to work intensity before and after the meeting ... and not just that the telephone does not ring. I think you should not be disturbed ... and you have to plan that [translated from Swiss German] (3).* |

**Supplemental material 9 revised programme theory including illustrative quotes and complete list of supporting quotes**

| |
|---|
| CMO configuration 4: **'need for relatedness'**<br><br>If a regular group of members engages in socially enjoyable contact, led by a skilled facilitator who, e.g. introduces people to each other, opens discussions, and clarifies and summarises statements (C), then group members will get to know each other and decide on rules that they are willing to follow, building a safe environment based on trust (O) because members want to be among and to interact with equals (M).<br><br>*I think it is important that we define the basic arrangements, like time, place, etc. and the procedure; that makes us feel safe. And respect and standards must be established and discussed, again and again. If this is done over and over again, there is a possibility to improve everything and give the participants the opportunity to express themselves; I think this is important [translated from Swiss German] (6).*<br><br>*With a glass of wine … it works best ... The satisfaction in QCs, this concerns my own role, or my emotional situation, because I am basically more satisfied if I have a fulfilling QC with my colleagues [translated from Swiss German] (3).* |
| CMO configuration 5: **'need for autonomy and control'**<br><br>If the group chooses its own topics and facilitator (C), then its members will feel they own the QC (O) because their need for autonomy - a feeling of being in control of their own behaviour - is satisfied (M).<br><br>*So, I think it is important that they are always the same participants and that the place and time frame are clear, and the distribution of tasks. This simply promotes trust in QCs and, on the basis of trust, you can have discussions between one colleague and another. And it is important that the QC can choose both the facilitator and the methods they want to work with [translated from Swiss German] (4b).* |
| CMO configuration 6: **'size of the group affects communication'**<br><br>If the group size exceeds 15 (C), then interaction among group participants decreases (O) because participants cannot keep up with each other and follow all conversations (M).<br><br>*When the group became too big, there were no interactions among participants any more. They all became listeners, and that was the main reason why we decided to split off and start a new QC [translated from Swiss German] (5).* |
| CMO configuration 7: **'feeling safe and not vulnerable'**<br><br>If participants trust each other (C), then they can describe how they work and admit what they do not know (O), because they feel safe rather than vulnerable (M).<br><br>*So, it takes a lot of courage to talk about a [clinical] case, especially if it didn't go really well. And if someone does tell such a story, then he is a role model, and others may have the courage to do the same and they gain trust on the one hand and, on the other hand, a certain feeling of togetherness [translated from Swiss German] (1).* |
| CMO configuration 8: **'need for competence and self-actualisation'**<br><br>If the facilitator supports participants and encourages them to tell their stories and share their experiences in a safe environment, e.g. by encouraging interactive responses, through discussions and by summarising statements (C), then participants will become involved and share their positive experiences and failures (O) because they want to improve their professional competence (M), gain professional confidence (M) and fulfil their professional potential (M). |

**Supplemental material 9 revised programme theory including illustrative quotes and complete list of supporting quotes**

| |
|---|
| *It's about your professionality [as a doctor] and in a case discussion you gain knowledge and learn a lot. Doing that, the doctors are totally focused on their work. (4b). [adds:] that's why I have the feeling that such case discussions are an important basis for the process ... if they are all peers, and everyone has experienced similar situations before, they can empower each other [translated from Swiss German] (4a).* |
| CMO configuration 9: **'previous knowledge is activated'**<br>If participants exchange case stories and experiences while actively listening to each other in the presence of a skilled facilitator in a safe environment (C), then they will share their knowledge by telling their own relevant stories (O) because the process activates knowledge they already possess (M).<br>*We discuss clinical cases, we show where we made decisions and mistakes ... and then we try to work out the principles, how to go on ... that's an important point, that you can relate difficult situations and the others support you and tell their stories [translated from Swiss German] (6).* |
| CMO configuration 10: '**immediate relevance for the practice'**<br>If QCs use the technique of experience-based learning (C), then knowledge becomes more relevant to GPs (O) because it relates to their everyday work and they can use it immediately (M).<br>*In the last QC, the topic was euthanasia … and I could use it immediately and was able to apply it. Discussing current cases … I often realise that the problem is actually also about my patients and that the topic helps in my everyday life [translated from Swiss German] (5).* |
| CMO configuration 11: **'cognitive dissonance'**<br>If participants discuss and reflect on their work processes (e.g. based on trustworthy data or personal experiences) during a professionally facilitated exchange of positive experiences or failures (C), then they discover knowledge gaps and identify learning needs and relevant topics (O) because their own attitudes and behaviours may differ from their peers', creating cognitive dissonance (a negative emotional state triggered by conflicting perceptions) that makes them reconsider their own way of working (M).<br>*Prescription data ... that would be very helpful if I had access to prescription patterns by simple means, or laboratory tests, just go and have a look [translated from Swiss German] (2).* |
| CMO configuration 12: **'social learning'**<br>If the facilitator uses purposeful didactic techniques (e.g. brainstorming, contentious or consensus discussions, or role-play) to keep the group active and to reward exploratory behaviour during reflection on the work process (C), then the group will create a learning environment that promotes knowledge exchange (O) because learning is a cognitive process in which participants observe and imitate their peers' behaviour to gain social approval (M).<br>*I think it has a lot to do with interactive learning; thanks to the support of evidence-based information, we are all at the same level of knowledge – right? Afterwards, we combine that with case discussions and that takes us further ... an important step ... [translated from Swiss German] (5).* |
| New CMO configuration at the group level: **'variety of characters stimulates reflection – cognitive dissonance'**<br>If members of the group have individual character traits and describe different professional experiences but accept each other's views (C), then they can learn from each other (O) because individual attitudes and behaviours will contrast with the knowledge of their peers and cause cognitive dissonance that makes them reflect on their way of working (M). |

**Supplemental material 9 revised programme theory including illustrative quotes and complete list of supporting quotes**

| |
|---|
| *I can't remember if we've talked about this before. But there's one point missing (in the programme theory) and that's the story that the group should have a say about who joins in. If the group is too different and everyone pulls in a completely different direction, it becomes difficult in QCs but it gets difficult as well if differences between participants are too small [translated from Swiss German] (2).*<br><br>*But I think GPs are generally people who want to get together and improve something; so, they are somehow a selection, but a varied selection, otherwise it doesn't work either ... [translated from Swiss German] (5).*<br><br>*Social learning is not without conflict but you have to be able to talk about different views without jumping down each other's throats... and talk about different views ... and that's sometimes a tightrope walk: how hard can I challenge someone – or do I have to treat someone gently? But I also think it depends on the person concerned – if it's someone who's anxious, I'll approach them rather gently as a facilitator [translated from Swiss German] (2).* |
| New CMO configuration at the individual level: **'threat to self-image – strong cognitive dissonance'**<br><br>If individuals feel too strong a cognitive dissonance when integrating new knowledge (C), then they may disrupt group dynamics and halt the QC process (O) because their self-image is threatened and they fear the loss of their professional identity (M).<br><br>*There are exceptions of, of individuals who cannot accept anything in any way and they are self-seeking and cannot learn anything new, who then, even if they participate, are not able to understand anything or to change practice [translated from Swiss German] (4a).*<br><br>*And of course, with time, you trust each other. And you open up, and then I have to find out if people fit in or not; unfortunately, there are a few people who don't fit in. Then decisions have to be made, i.e. they have to leave, if they can't deal with the group or accept something new. Otherwise QCs can't work [translated from Swiss German] (6).*<br><br>*If there is a disrupting feature in it, in the group, that hinders the group from norming, then I have to deal with it. For example, if someone always withdraws, then I have to ask the individual, in front of the group or maybe better in private [translated from Swiss German] (2).* |
| CMO configuration 13: **'positive interdependence between health insurance companies (or administration at the national level) and GPs'**<br><br>If physician network organisations require continuous QC activities (C), then QCs will negotiate priorities and design creative solutions (O) because the tension between autonomy and obligation spurs the group to act and negotiate together to reach a common goal (M).<br><br>*That is certainly a driver to improve quality, and the physician network or the responsible organisation must have the resources to apply these results and at the same time support the QCs, for example with scientific knowledge [translated from Swiss German] (2).* |
| CMO configuration 14: **'threat to professional autonomy'**<br><br>If GPs feel that the QC programme is only a top-down managerial intervention to reduce costs (C), then they will not be motivated and will not participate (O) because they feel unsafe and fear they lack autonomy in their clinical role (M).<br><br>*No data* |
| CMO configuration 15: **'positive interdependence among group members'**<br><br>If participants maintain a learning environment based on trust that promotes the exchange of knowledge, assisted by facilitators who use professional techniques (e.g. contentious discussion, reaching consensus and |

role play) (C), then participants will adapt and generate new knowledge for local use (O) because they see themselves as similar, and so act and negotiate cooperatively to achieve a common goal (M).

*The reason why the group drafts such recommendations and continues to work on them is, I really think, so that the group says, 'okay' we need to discuss how to put it together and see if we can use it, and if a facilitator skilfully tackles that and steers the process in a good direction so the group participates, and then the group really does it and creates something new, they get the feeling that we've done it ourselves now [translated from Swiss German] (3).*

---

CMO configuration 16: **'identifying and removing barriers to change'**

If participants, supported by skilled facilitators, address barriers to change (C), then they are more likely to implement the innovation (O) because participants help each other to develop strategies to identify and overcome these barriers (M).

*But I don't think barriers are a big problem, because you hardly ever put up relevant barriers if you are there, participating in the QC, and address and solve them during the process [translated from Swiss German] (2*).

---

CMO configuration 17: **'need for competence, autonomy and relatedness'**

If participants create new knowledge and plan an implementation strategy (C), then they feel satisfaction, responsibility and stewardship (O) because this fulfils their need for competence (being able to achieve specific objectives) (M), autonomy (a feeling of being in control of their own behaviour) (M) and relatedness (a sense of connection to a larger group) (M).

*And you're satisfied because you were involved, and so you can relate to the content [of the innovation], and that's probably why it works so well; it was well done and I can identify with it ... and support it ... and you feel like you developed it or helped develop it [translated from Swiss German] (4a).*

---

CMO configuration 18: **'intention to change'**

If participants publicly announce their intention to change (C), then they are more likely to implement the change (O) because they and others in the group all think it is a good idea and believe they can carry it through (M).

*And that gives me a feeling of significance and satisfaction in QCs, when I've done something like this. And also when I intend to use it [new knowledge] ... the facilitator helps me if he takes me through the PDCA cycle. In this phase, the facilitator is very important, important for planning, because he helps me when I have to plan; that is, when the members or at least one of them has decided to change behaviour [translated from Swiss German] (2).*

---

CMO configuration 19: **'testing new knowledge'**

If participants validate and test new knowledge in a QC, moderated by a skilled facilitator in a safe environment (C), then they feel confident putting that knowledge to use in everyday practice (O) because they have had the opportunity to practise and familiarise themselves with the innovation (M).

*No data*

---

CMO configuration 20: **'gaining confidence in an innovation'**

If the group repeatedly practises implementing and adjusting to an innovation (C), then its members trust their own competence and turn the innovation into a habit (O) because successful outcomes increase their confidence in their abilities (M).

**Supplemental material 9 revised programme theory including illustrative quotes and complete list of supporting quotes**

| |
|---|
| *But to recognise whether the process actually leads to a change in my own behaviour, I have to have an outside indicator that shows it to me, time and again. In principle, this means that I have to have an external objective assessment of what I am doing and whether anything is changing – and this helps to change my behaviour and to gain confidence in the innovation [translated from Swiss German] (1).* |
| CMO configuration 21: **'repetition priming and automaticity'**<br>If participants build a regular group and practice using QI tools (C), then they will successfully implement new knowledge into everyday practice (O) because responses improve with repetition: 'practice makes perfect' (M).<br><br>*It is a fact that you use the same techniques over and over again, in terms of methodology – and don't even notice it ... so, even if you don't notice it, it helps to improve quality in PHC, that's for sure, that's absolutely right [translated from Swiss German](3).* |

### Quotes of interviews in Phase III

**Sources:**

(1) S.S. GP and manager, CEO of Santémed network of primary health care centres owned by health insurance company
(2) O.R., GP, tutor in MediX Bern, a doctor-owned network of primary health care centres in Bern
(3) M.J., GP and manager, facilitator and member of Sanacare's management board, a network of primary health care centres owned by health insurance companies
(4) V.M (4a). + E.K. (4b), social scientists, representatives of the professional body Swiss Association of Medicine
(5) O.S., GP, facilitator of a QC belonging to MediS Zürich, a doctor-owned network of primary health care centres, Professor at the Institute of General Practice in Zürich
(6) S.K., GP, executive for General Practice at the hospital in Aarau

#### *Preconditions*

Context mechanism outcome configuration 1: **'participants know what to expect'**

If the introductory workshop conveys the principles of QI in PHC and the workings of QCs (social persuasion) (C), this will increase future participants' motivation to join QCs (O) because they learn what to expect and may feel that they are capable of meeting expectations (increase of self-efficacy) (M).

*Well, if it's set up that way, I'm sure it'll work. [translated from Swiss German] (3).*

*So, I think that everything should be well organised and planned, and that you really have to understand what it's all about – as far as you can understand it without having gone through the process at least once [translated from Swiss German] (2).*

**Supplemental material 9 revised programme theory including illustrative quotes and complete list of supporting quotes**

Context mechanism outcome configuration 2: **'need for autonomy'**

If the administration at the national level or at the level of health insurance companies entrusts GPs with QI and autonomy (puts them in control of how do it) (C), then GPs may consider participating in QCs (O) because they feel they can take on the responsibility and make a difference (M).

*You actually buy your network participation or membership through QC attendance. This is sort of obstructive because it results in a kind of obligation for the participants. But the opportunities that such a network offers with different QCs reduces the problem because you have different choices. You find people who you may want to work with in your area. This kind of organisation may fit your needs [translated from Swiss German] (5).*

*Yes, of course, these are specific drivers to meet, and after all it being member of a physician network is like a commitment to participate in QC…at the same as you may fear limitation in your professional autonomy etc… but there are rules and if everybody sticks to these rules will work [translated from Swiss German] (3).*

*The physician networks are very open as to QCs. They do not set any rules on what is done or dealt with in the QCs, there is simply a requirement that participants must conduct QCs [translated from Swiss German] (2).*

In order to account for the interviews data, I developed an additional context mechanism outcome configuration at the organisational level: '**feeling of having a say'**

If an organisation, (e.g., a physician network organisation) has a decentralised policy that encourages use of local knowledge (C), then the QC takes on tasks (O) because they feel that they have a say in QI in their practice (M).

*I was asking myself what autonomy is and what it actually means...so decentralised organisation means accepting local decisions and having a flexible administration. So not simply stubborn and rigid, but adaptable to local decisions, which are then taken by the QC, for example [translated from Swiss German] (2).*

*So, if I want to promote and do QCs, then it's in the nature of the QC that different groups come up with different solutions. And you have to be able to live with them and you also have to be able to endorse them as an organisation, otherwise the QC instrument wouldn't make sense [translated from Swiss German] (1).*

*For this reason, it is certainly beneficial if QCs are as decentralized as possible and adapted to local conditions; but I also think that depending on the network and organisation, certain formal requirements must be in place. And if someone joins this network, then they also accept the conditions to a certain extent [translated from Swiss German] (5).*

*If you recognize positive things that you can perhaps implement well within a short time, or sometimes slowly, then of course you have to; if you don't do that, you nip further initiatives in the bud, because then no further good suggestions will come if nothing is ever implemented. People lose motivation [translated from Swiss German] (3).*

*I think that if something like this is developed by a QC now, then there should also be appreciation from the organisation. [translated from Swiss German] (3).*

*If we want to discuss something and talk about it, about a vitamin or whatever, it doesn't matter, then the organisation should support it. It should be interested that we get prescription data to objectify our behaviour and also to show any progress that may result from the whole discussion. Or whatever, but I believe that the organisation should make this possible; that is important [translated from Swiss German] (3).*

**Supplemental material 9 revised programme theory including illustrative quotes and complete list of supporting quotes**

*As I have described before, I believe we should have certain freedom to implement new things in an organisation. At the same time, it should be clear what is important to the organisation. Organisations should also provide knowledge and create a kind of platform for knowledge exchange [translated from Swiss German] (3).*

#### *Establishing the group*

Context mechanism outcome configuration 3: **'sharing similar needs'**

If the administration at the organisational level of QCs provides administrative support (i.e. training of facilitators, data gathering and provision of evidence-based information), protected time and space, CME points, and small financial incentives to QC participants (C), then they will meet to exchange ideas (O) because QCs are GPs' preferred learning style (M), support generates positive expectations among participants (M), and GPs think QC meetings with their peers will be useful (M).

*Yes, such a network allows high organisational and professional autonomy...and this is a good start for QCs [translated from Swiss German] (2).*

*For me this (protected time) is more than just the aspect of time, it relates to the amount of the work before and after the meeting....and not just that no telephone rings. I think you should not be disturbed...and you have to plan that [translated from Swiss German] (3).*

*It benefits the participants a lot, that's no question. I just ask myself how do we manage to take away 1½ to 2 hours from the working day without increasing the amount of the work? Basically, this is probably an organizational problem. In an ideal world, we should have time to discuss things with each other and form a basis for identification, GP-based, mutual trust, a discussion of sensitive issues, and trying something new together [translated from Swiss German] (3).*

*The time in the QC, that is the time during which no phone calls should come in, and it should also have no influence on the amount of the work that day. It is a little easier with the internal staff, by that I mean the participants from here. Those with a longer journey have more difficulties [translated from Swiss German] (2).*

*I believe that 1½ hours should be enough for a QC according to our experience. It is extremely important that you are not disturbed. And the amount of work on the same day must not increase because of the QC. After all, this is an integral part of the work and also of the contract with the network [translated from Swiss German] (4b).*

*If I do it this way, it's probably easier later. For example, I have to know how much money is available from the network; this is important if the implementation of the ideas also involves costs [translated from Swiss German] (4a).*

*So really a fee to participate or a fee for the facilitator did not exist. And the further education points were not a big issue either. But now that we have that, a small fee and CME points is certainly beneficial. [translated from Swiss German] (2).*

*So, I think if participants know that there is adequate compensation for the time they spend, then it is an important complement to intrinsic motivation. And naturally also the CME points. [translated from Swiss German] (5).*

**Supplemental material 9 revised programme theory including illustrative quotes and complete list of supporting quotes**

*Well, I just think it's very different how a QC forms. There are people at the end of their career who are almost burnt out and want to exchange ideas, in the sense of improving their well-being. And at the other end of the spectrum, there are newcomers or beginners who are at the beginning of their GP career and are intrinsically highly motivated; they simply want to exchange experiences and clinical information [translated from Swiss German] (5).*

*Sometimes the organisation or network is more important. In a network all participants have to take part in QCs because it is a standard. Or you take part because you become simply more and more interested. Sometimes it starts as an obligation and becomes an intrinsic motivation] (4b).*

*Well, for me, if I look at it that way, there doesn't have to be a cause. I think the motivation to participate has to be there before, otherwise you wouldn't take part in QCs. But I think the logistical and practical support is important. First of all, the intrinsic motivation has to exist; obligation is an extra driver, as is administrative support; all that is important. [translated from Swiss German] (5).*

*So, I have the feeling that the incentives that are mentioned do not matter so much, but the philosophy and the attitude of the doctor. I believe that if he is committed to his profession, then a little money will simply speed up this process a little. [translated from Swiss German] (3).*

*I think it takes a basic motivation to understand the purpose of QCs and to invest 1½ hours to improve over and over again. I believe it takes a certain amount of reflection and openness to continue developing [translated from Swiss German] (3).*

*It simply needs something more, everything we have already discussed, but also something else; it is a kind of commitment to the profession and you want to feel comfortable in your position. [translated from Swiss German] (3).*

*I think intrinsic motivation is the basic requirement but it mostly takes an outside force to get different GPs to come together and actually take the time to do this [translated from Swiss German] (1).*

*Peer consulting is certainly a motivating factor for participating in QCs, but the networks create the structure to make it possible. Smaller monetary values only play a minor role, I think. [translated from Swiss German] (4b).*

*Personally, I would be motivated if I knew that someone with a lot of experience was facilitating the QC. But also, the subject-specific aspects are part of it, the peers know what they are talking about [translated from Swiss German] (4a).*

*In the beginning, we had no organizational help. In the course of time this has changed to the extent that two more QC participants joined the network because it is required that they participate, but it helps with organising them [translated from Swiss German] (2).*

*Yes, that they help to organize a room, that they help to take minutes, that they help us to develop our own initiative. You should also help us with the design of the content like evidence-based material [translated from Swiss German] (2).*

*So, it is important that there are rooms available that are easily accessible for everyone in terms of time. Up to now it has been over lunch. I think it is beneficial if there is some kind of catering available because most people go straight to the office afterwards. I think this logistical aspect is important [translated from Swiss German] (5).*

*There are certainly different needs in terms of location. Some people appreciate being able to do their QCs in the practice, others prefer to meet privately. Others may simply want a meeting room. We have sensed a variety of needs. Some really want to stay in their practice environment and others choose a completely different location [translated from Swiss German] (4b).*

**Supplemental material 9 revised programme theory including illustrative quotes and complete list of supporting quotes**

Context mechanism outcome configuration 4: **'need for relatedness'**

If a steady group of members engages in socially enjoyable contact, led by a skilled facilitator who, e.g., introduces people to each other, opens discussions, clarifies and summarizes statements (C), then group members will get to know each other and norm rules that they are willing to follow and build safe environment based on trust (O), because members want to be among and to interact with equals (M).

*I think it is important that we define the basic arrangements, like time, place, etc., and the procedure; that makes us feel safe. And respect and standards must be established and discussed, and again and again. If this is done over and over again, there is also the possibility of improving everything, and gives the participants the opportunity to express themselves; I think this is important [translated from Swiss German] (6).*

*There are important cornerstones such as time, duration, division of tasks and also the process of a QC; it is also important to determine norms of communication with each other regarding feedback and communication culture. These two things are actually prerequisites for trust and security as a basis for open communication. I have never experienced stormy times, as often described in group development. I think you have to come back to this and discuss together what the rules of the game are. And if it doesn't work out that way, you can simply ask: where is the problem [translated from Swiss German] (2)?*

*The QC takes place in an open circle and with a facilitator who opens the discussion. He clarifies statements and summarizes. The activities at the beginning usually consist of case discussions. The session should be as open as possible at the beginning and everyone who has something on their mind should have their say [translated from Swiss German] /2).*

*In existing QCs, I think it would be good if the rules of the game were reasonably clearly defined [translated from Swiss German] (2).*

*The facilitators spend a relatively large amount of time determining the timing of the sessions so that it is convenient for everyone. I think this is a rather delicate process in which the group does not always reach consensus immediately. But with time, this will become part of the process. It's relatively important that you do it together and no one gets left behind [translated from Swiss German] (2).*

*By the way, I always put the basic points up for discussion, also with regard to respect and communication culture. That works somewhat differently in every QC. Individual groups have, how shall I put it, their own character [translated from Swiss German] (2).*

*It's important to discuss respectfully how to deal with each other, etc.; this is not only important for the development of the group, but also for what comes later in the QC in terms of the QI process. Sometimes small groups of people form who like each other from the past and this is not always helpful [translated from Swiss German] (5).*

*It was enormously important to me that the general conditions were made clear: when do we meet? where do we meet? how often? Then it was important for me that this wasn't a one-man show, where the facilitator didn't just do all the work, but that he had the competence to lead the group and distribute the tasks [translated from Swiss German] (5).*

*I think it also has to do with the fact that you get to know people better over time. You know what they are like, and if they don't always get along so well with each other, I can still ask them to be respectful to each other in the specific case [translated from Swiss German] (5).*

**Supplemental material 9 revised programme theory including illustrative quotes and complete list of supporting quotes**

*With a glass of wine… it works best...The satisfaction in QCs, this concerns my own role, or it concerns my emotional situation, because I am basically more satisfied if I have had a fulfilling QC with my colleagues [translated from Swiss German] (3)*

*You have to set the rules at the beginning and if the rules are there, it works; and if the rules are not there, then some people don't know what to do and how to do it, and then it doesn't work. And from this I can only confirm: you need the rules [translated from Swiss German] (1).*

*Confrontation rarely happens, but when it does, it's usually not bad because it means that someone really .... that the group is really engaged and can participate in 'confrontation' at all. If someone is not involved, they will never get upset. And the moment someone gets upset, it means that they, the ones, who bring the topic into the group are extremely interested in what they do.... So, when it happens it is a very good step that leads to improvement. [translated from Swiss German] (1).*

*Actually, two things: one is that there is the framework we discussed before, and the other is that confidential things remain confidential. So that it really stays within the framework in which it's articulated and doesn't leave the room. [translated from Swiss German] (1).*

*This brings me to another aspect: because patient cases are discussed, it is very important that confidentiality and patient confidentiality are guaranteed. So, there must be a protected setting. I also have the feeling that this must be made very clear and that everyone in the circle agrees with it [translated from Swiss German] (4b).*

*I have the feeling that you have to do it as in a project team: in a new committee, you first agree on the joint procedure and record it in detail. In the session, they agree on how to communicate and how to organise things, and everyone has to say something about it or at least nod briefly [translated from Swiss German] (4a).*

Context mechanism outcome configuration 5: **'need for autonomy and control'**

If the group chooses its own topics and facilitator (C), then they will feel they own the QC (O), because this satisfies their need for autonomy (a feeling of being in control of one' s own behaviour) (M).

*So, I think it is important that they are always the same participants and that the place and time frame are clear, and the distribution of tasks. This simply promotes trust in QCs and on the basis of trust you can have discussions, from colleague to colleague. And that the QC can choose both facilitator and the methods they want to work with [translated from Swiss German] (4b).*

*Regarding autonomy, we are free to decide when, where and how long the QCs will last. In pilot tests, however, the duration has levelled off at 1½ hours. The network imposes certain conditions regarding certification. These require that we have to do a certain number of QCs per year and that we regularly review some topics but are free to choose the content [translated from Swiss German] (2).*

Context mechanism outcome configuration 6: **'size of the group affects communication'**

If group size exceeds 15 (C), then interaction among group participants decreases (O) because participants cannot keep up with all participants and follow their conversations (M).

*When the group became too big, there were no interactions among participants anymore. They all became listeners, and that was the main reason why we decided to split off and start a new QC [translated from Swiss German] (5).*

**Supplemental material 9 revised programme theory including illustrative quotes and complete list of supporting quotes**

*The principle is that less technology helps and that the group sits in a circle and thus communicates better [translated from Swiss German] (5).*

#### *Learning environment*

Context mechanism outcome configuration 7: '**feeling safe and not vulnerable'**

If participants trust each other (C), then they can disclose how they work and also the holes in their knowledge (O), because they feel safe rather than vulnerable (M).

*So, it takes a lot of courage to talk about a [clinical] case, especially if it didn't go really well. And if someone does tell the story, then he is a role model, and others may have the courage to do the same and they gain trust on the one hand and, on the other hand, a certain feeling of togetherness [translated from Swiss German] (1).*

*It benefits the participants a lot, that's no question. I just ask myself how do we manage to take away 1½ to 2 hours from the working day without increasing the amount of work? Basically, this is probably an organizational problem. In an ideal world, we should have time to discuss things with each other and form a basis for identification, GP based, mutual trust, a discussion of sensitive issues, and trying something new together [translated from Swiss German] (3).*

*This is consistent and fits, I think this is exactly how the QC reveals knowledge gaps [translated from Swiss German] (3).*

*The QC takes place in an open circle and with a facilitator who opens the discussion. He clarifies statements and summarizes. The activities at the beginning usually consist of case discussions. The session should be as open as possible at the beginning and everyone who has something on their mind should have their say [translated from Swiss German] (2).*

*I have the feeling that QCs work and the chance of this increases if the QC takes place in a good atmosphere. Certainly, mutual trust is extremely important in the interaction. In the group, everyone has their own knowledge, which they make available to the group and thus increase the competence of the group [translated from Swiss German] (4a).*

*It would certainly be a good basis for discussion if you really had facts and not just the feeling of how you work with your patients, then you really have facts. But an atmosphere of trust is necessary so participants can discuss their data in the QC [translated from Swiss German] (4a).*

Context mechanism outcome configuration 8: **'need for competence and self-actualisation'**

If the facilitator supports participants and encourages them to tell their stories and share their experiences in a safe environment by, e.g., encouraging interactive responses, discussions, and summarizing statements, (C) then participants will be involved and share their experiences and failures (O), because they want to improve their competency (a sense of self-efficacy to achieve specific objectives) (M) and gain professional confidence (M), and achieve professional self-actualisation (M).

*It's about your professionality as a doctor and in a case discussion you gain knowledge and learn a lot. Doing that, the doctors are totally focused on their work (4b) adds: that's why I have the feeling that such case discussions are an important basis for the process...if they are all peers, and everyone has experienced similar situations before, they can empower each other [translated from Swiss German] (4a and b).*

**Supplemental material 9 revised programme theory including illustrative quotes and complete list of supporting quotes**

*And the case discussions are a very good way of getting into the subject. I often do it this way. At the beginning, however, we always start with a general opening, when everyone who has something very urgent to recount can ask questions, especially if she needs an answer for her clinical work [translated from Swiss German] (5).*

*This part of the programme theory is certainly correct. I do believe that QCs can relieve worries and fears, maybe even help you get rid of worries because you can discuss difficult things and share experiences that are tough [translated from Swiss German] (3).*

*The active participation in discussions of cases and personal experiences gives the feeling of group cohesion and a kind of understanding culture. There is also a feeling of openness and mutual support, and trust increases, a kind of collegiality. In this atmosphere, a collegial influence is possible and medical problems that are important to people are discussed [translated from Swiss German] (2).*

*This part of the programme theory also corresponds to my experience. I think the participants realize in these case discussions that they partly have the same problems. And this creates the feeling of belonging together as GPs and being in the same boat [translated from Swiss German] (5).*

*So, I think in the first part of the process, facilitators are very important because they stimulate people to actively participate. Facilitation is certainly important, but also the opportunity to discuss cases that are important to participants, that is basics [translated from Swiss German] (5).*

Context mechanism outcome configuration 9: **'previous knowledge is activated'**

If participants exchange case stories and experiences while they actively listen to each other in the presence of a skilled facilitator in a safe environment (C), then they will share their knowledge by telling their own relevant stories (O), because the process activates knowledge they already possess (M).

*We discuss clinical cases, we show where we make decision and mistakes... and then we try to work out the principles, how to go on... that's an important point, that you can relate difficult situations and the others support you and tell their stories [translated from Swiss German] (6).*

*So, in that setting, I think, you need your own case studies because otherwise, if someone doesn't talk about their own cases, then it's just another lesson in school and so the emotional involvement will be much, much lower and the interesting thing is, really, when someone gives something with heart and soul. And that lifeblood is then what motivates the others to participate [translated from Swiss German] (1).*

*Case discussions are important for the group. They are also exciting. We had chosen a broad topic in the QC that I facilitate. The topic of choice we made was actually an impossible one. We wanted to work on a guideline and then realized that it was completely useless. I then told the participants that they should simply take their own real-life examples for discussion, and that works well! [translated from Swiss German] (2).*

*Yes, exactly! You simply have to be open for any case vignettes that come from practice. They then form the basis for the topic that participants choose. For example, if someone asks you how to detect these 'damn' food allergies, what serological test to do. Then as a facilitator you have to get to the topic, so that everybody can understand and deepen their knowledge [translated from Swiss German] (5).*

*As a facilitator, I help the group to choose the relevant topics from the case discussions, which are then covered separately in a QC [translated from Swiss German] (5).*

**Supplemental material 9 revised programme theory including illustrative quotes and complete list of supporting quotes**

Context mechanism outcome configuration 10: **'immediate relevance for the practice'**

If QCs use the technique of experience-based learning (C), then knowledge becomes more relevant to GPs (O), because it relates to their everyday work and is therefore of immediate use (M).

*In the last QC, the topic was euthanasia… and I had immediate use of something and was able to apply it. Discussing current cases…. I often realize that the problem is actually also about my patients and that the topic helps me in my everyday life [translated from Swiss German] (5).*

*It took some time before the discussion got going. At the end I asked them how they wanted to continue. The group was unanimous that they wanted to look at cases and then decide how to proceed based on the cases [translated from Swiss German] (2).*

*If the facilitator succeeds in actually uncovering the path of decision making, i.e. what the patient says and the doctor does with this... if he succeeds in working it out and communicating it in an emotional and lively way, then I think that something very important will happen. If the QC is like reading a textbook, then it won't help anyone. But if the group succeeds in tracing the path of a decision-making process and working out the critical aspects or pitfalls within the framework of this decision-making process, then the person who tells the case is personally affected. And in the moment when that person is personally affected and can express this, the rest of the group is usually also addressed and personally affected. And these are then the moments I recall, at least that's how I feel, and I will do that automatically, when I meet the next patient, and benefit from the discussion a great deal [translated from Swiss German] (1).*

Context mechanism outcome configuration 11: **'cognitive dissonance'**

If participants discuss and reflect on their work processes (e.g. based on trustworthy data or exchange of personal experiences) during a professionally facilitated exchange of positive experiences or failures (C), then they discover knowledge gaps and identify learning needs and relevant topics (O) because their own attitudes and behaviours may contrast with their peers' knowledge, causing cognitive dissonance that makes them reconsider their way of working (M).

*Prescription data.... that would be very helpful if I had access to prescription patterns by simple means, or laboratory tests, just go and have a look [translated from Swiss German] (2).*

*This seems familiar to me. I think we are in a special situation in our physician network, where we actively work with guidelines [translated from Swiss German] (5).*

*Our physician network collects statistical evaluations on a practical level, where they present us with the results and benchmarks. Of course, confidence-building measures play an important role in this process so that the group can discuss the results together. I don't think that this requires so much skill from the facilitator, because the statistics are stimulating enough and everybody wonders why he is where he is, statistically [translated from Swiss German] (5).*

*I don't think statistics cause fear but motivate people to change their behaviour and for example prescribe more generics if a patient requires, for example, a proton pump inhibitor. [translated from Swiss German] (5).*

*For example, when I look at all my referrals, I can't recall why I wrote them. Then, when I discuss this with my colleagues, my colleagues can explain ninety nine percent of the time a plausible reason for the referral and I can learn from these discussions. [translated from Swiss German] (5).*

**Supplemental material 9 revised programme theory including illustrative quotes and complete list of supporting quotes**

*Yes, when seven to ten different opinions have been expressed, then the group should use evidence-based information. They just have to look up what is going on and revise their views. Sooner or later you always arrive back at guidelines. [translated from Swiss German] (3).*

*Yes, somehow participants must recognize and bridge the gaps that appear in the discussion. The QC is a good place to reflect on whether something is correct or not, or whether you have handled a situation correctly or not [translated from Swiss German] (3).*

*It is important that the group first comes to an issue, where there is some dissent, isn't it? [translated from Swiss German] (3).*

*Yeah, this is the part where the process moves into the QI. We don't just sit there and discuss cases; we reflect on them. I think about what I have heard and wonder whether I understood my colleagues correctly, it's all about self-reflection. I ask myself, is that true, do I do really do it that way, and why, and how could I do it better? Afterwards, the group then moves to the personal level of action, where quality in everyday practice improves. [translated from Swiss German] (4a).*

*I have the feeling that participants realize that someone else may be right, even if they don't admit it right away. I have the feeling that on the journey home or at home, or later at some point, especially facing similar cases, they think of the discussion. Yes, and then you do not automatically do what you have always done, but you start considering other possibilities for solving the problem. Then, participants may actually change their attitude and behaviour or consult a colleague or take a look in the books. [translated from Swiss German] (4a).*

*I believe it would be important to have data and to know what is going on in order to reflect on what we are doing and to improve the quality of our work [translated from Swiss German] (4a).*

*For example, if there is a topic that concerns a certain medication, then I hardly know how often I prescribed the drug in question. Then it's good if I can see in reality what I'm really doing. On the basis of routine data that can easily be compiled, GPs could find out how often they prescribe something, also compared to other colleagues in Switzerland [translated from Swiss German] (4a).*

*It would be good if we had prescription data on any clinical topic; you should be able to see what the reality is in practice on any topic. For example, we have done a feasibility study about drug interactions. Based on the data, GPs could recognise where the most common problems were and take action. [translated from Swiss German] (6).*

#### Context mechanism outcome configuration 12: **'social learning'**

If the facilitator uses purposeful didactic techniques (e.g., brainstorming, contentious or consensus discussions, or role play) to keep the group active and to reward exploratory behaviour during reflection of the work process (C), then the group will create a learning environment that promotes knowledge exchange (O) because learning is a cognitive process in which participants observe and imitate their peers' behaviour to gain social approval (M).

*I think it has a lot to do with interactive learning; thanks to the support of evidence-based information, we are all at the same level of knowledge - right? Afterwards we combine that with case discussions and that takes us further....an important step [translated from Swiss German] (5).*

*It is important in this phase as a moderator to actively request opinions from the group and not simply throw general questions at the group [translated from Swiss German] (2).*

**Supplemental material 9 revised programme theory including illustrative quotes and complete list of supporting quotes**

*I think this exchange of knowledge in the group needs great trust and a safe climate. It's about confidence in dealing with discussions, in dealing with colleagues, so that they can show weaknesses. The others might think it's a funny story at first, but often they recognize themselves in it. The facilitator needs to ensure that the participants maintain respect and tolerance. After all, there are no false statements. A facilitator needs excellent training to be able to convey this feeling. And the training should be professional [translated from Swiss German] (3).*

*What helps me least, how shall I put it, are the quantifiable areas that can be measured as outcome. What helps me most is the path that leads to a decision. And measuring results afterwards is a tool for estimating roughly where I am moving to, but it has little to do with me as a person. But more importantly, I can develop professionally in a QC, I can learn how to take a decision and to make the process of decision making transparent [translated from Swiss German] (1).*

*If there is, perhaps, a need for clarification in the group. If some people say that they always do it for this or that reason and others reply that they judge the problem quite differently and therefore, they do it this way. It helps with evidence-based information and data that show how people really work. [translated from Swiss German] (4a)*

*So, whenever we look at our data, we start being distracted by exceptions and we lose the overall context, i.e. what is actually at stake. And when you have a sheet of numbers in front of you and you don't know how they came about; it becomes difficult to interpret. Misconceptions quickly happen and I have my doubts that if you just focus on a few small exceptions, that you see how GPs really work. Numbers should only be looked at in connection with case discussions [translated from Swiss German] (4b).*

In order to account for the interviews data, I developed an additional context mechanism outcome configuration: '**Variety of characters stimulates reflection – stimulating cognitive dissonance'**

If members of the groups have individual character traits and describe differing professional experiences but accept each other's views (C), then they can learn from each other (O) because own attitudes and behaviours will contrast with their peers' knowledge and cause cognitive dissonance that makes them reflect on their way of working (M).

*Social learning is not without conflict, but you have to be able to talk about different views without chopping each other's hand off......and talk about different views...and that's sometimes a tightrope walk: how hard can I challenge someone - or do I have to treat someone gently? But I also think it depends on the person it concerns - if it's someone who's anxious, I'll approach them rather gently as a facilitator [translated from Swiss German] (2).*

*I can't remember if we've talked about this before. But there's one point missing, in the programme theory, and that's the idea that the group should have a say about who joins in. If the group is too different and everyone pulls in a completely different direction, it becomes difficult in QCs, but it gets difficult as well if the differences between participants are too small [translated from Swiss German] (2).*

*But I think GPs are generally people who want to get together and improve something.; so, they are somehow a selection, but a varied selection, otherwise it doesn't work either [translated from Swiss German] (5).*

**Supplemental material 9 revised programme theory including illustrative quotes and complete list of supporting quotes**

In order to account for the interviews data, I developed an additional context mechanism outcome configuration: **'threat to self-image' to account for the interviews data.**

If individuals cannot cope and integrate new knowledge (C), then they disrupt group dynamics and the QC process halts (O) because new knowledge threatens their self-image and they feel at risk of losing their professional identity (M)

*There are exceptions of, of individuals who cannot accept anything in anyway and; they are self-seeking and cannot learn anything new, who then, even if they participate, are not able to understand anything or to change practice [translated from Swiss German] (4a).*

*And of course, with time, you trust each other. And you open up, and then you have to find out if people fit in or not; unfortunately, there are a few people who don't fit in. Then decisions have to be made, i.e. they have to leave; if they can't deal with the group or accept something new. Otherwise QCs can't work [translated from Swiss German] (6).*

*If there is a disrupting feature in the group that hinders the group from norming, then I have to deal with it. For example, if someone always withdraws, then I have to ask the individual, in front of the group or maybe better in private [translated from Swiss German] (2).*

#### *Adapting, creating and testing new knowledge*

Context mechanism outcome configuration 13: **'interdependence between insurers / physician network organisations / national administrations and GPs'**

If national administrations require continuous QC activities (C), then QCs will negotiate priorities and design creative solutions(O) because the tension between autonomy and obligation spurs the group to act and negotiate together to reach a common goal (M).

*I also think intrinsic motivation is an important point for someone to participate in a QC. But especially in our network, it is not only intrinsic motivation, it is also a requirement [translated from Swiss German] (5).*

*That is certainly a driver for improving quality and the physician network or the responsible organisation must then have resources to apply these results and at the same time support the QCs, for example with scientific knowledge [translated from Swiss German] (2).*

*I think I can sign up to these points of programme theory. In our case it is our network that forms and maintains the organization in the background, that supports the QC, but also requires participation [translated from Swiss German] (5).*

*I completely agree with the programme theory here and I think it points the finger at an important point. I could give you many examples to confirm this. You have to demand change to a certain degree [translated from Swiss German] (2).*

*It is the balance between autonomy and then still being able to demand something, even if it is mainly at the organisational level. As an incentive, small financial compensations for the participants come to mind [translated from Swiss German] (2).*

*The question is then, as many participants then of course may think, okay, I would like to devote myself to an important topic. But these topics should then also be important for the whole organisation and that may then have a stimulating effect [translated from Swiss German] (3).*

**Supplemental material 9 revised programme theory including illustrative quotes and complete list of supporting quotes**

Context mechanism outcome configuration 14: **'threat to professional autonomy'**

If GPs feel that the QC programme is only a top-down managerial intervention to reduce costs (C), then they will not be motivated and not participate (O) because they feel unsafe and lack autonomy in their clinical role (M).

*no data*

Context mechanism outcome configuration 15: **'interdependence among group members'**

If participants maintain a trusting learning environment that promotes knowledge exchange, assisted by facilitators who use professional techniques (e.g., contentious discussion, reaching consensus, and role play), (C), then participants will adapt and generate new knowledge for local use (O) because they see themselves as similar, and act and negotiate cooperatively to reach a common goal (M).

*I was just thinking about the means to challenge the QC. Basically, I think you can stimulate a discussion, even a controversial one. Then the group debates, thinks in a circular fashion, uses role play and they stimulate one other to find new solutions. [translated from Swiss German] (5).*

*At the beginning of a QC, there is hardly any consensus in a group regarding therapy. But then, if you work on it in circles and look at it from different perspectives, and are motivated to improve, then there is an incentive within the group to align [translated from Swiss German] (5).*

*The reason why the group drafts such recommendations and continues to work on them is, I really think so, that the group says, 'okay' <u>we</u> need to discuss how to put it together and see if <u>we</u> can use it, and if a facilitator skilfully tackles that and steers the process in a good direction so the group participates, and then the group really does it and creates something new, they get the feeling that we've done it ourselves now [translated from Swiss German] (3).*

*Sometimes you don't understand it right from the start. But then you can say, okay, then I'll just try with a few patients for the next two or three months. Especially if something's a common problem. Then I can take a closer look at it again in the QC and slowly change my behaviour bit by bit [translated from Swiss German] (4a).*

*I can also well imagine the following happening, and it does in our case: someone reads an article and wants to discuss it in the group, because something new is recommended, something that should be changed in practice. But before I change something, I first want to know what my peers think about it. And I often see that it is only then that something new is actually introduced [translated from Swiss German] (4a).*

Context mechanism outcome configuration 16: **'identifying and removing barriers to change'**

If participants, supported by skilled facilitators, address barriers to change (C), then they are more likely to implement the innovation (O), because participants help each other develop strategies to identify and overcome these barriers (M).

*It is useful to identify barriers in advance, as for example in drug prescription projects. It increases the willingness to change. For example, there was a lot of resistance to the discussion of antidepressants, because most colleagues simply didn't want to change anything. It was good that we had discussed this in advance [translated from Swiss German] (5).*

**Supplemental material 9 revised programme theory including illustrative quotes and complete list of supporting quotes**

*Innovations create opposition and nothing else is true. I think that's clear. And sometimes I have to bring it up, so that it becomes clear even before problems arise. We must then remain consistent and stick to our goal, even if we talk a lot about these barriers [translated from Swiss German] (2).*

*But I don't think barriers are a big problem, because you hardly ever put up relevant barriers if you are there, participating in the QC, and addressing and solving them during the process [translated from Swiss German] (2).*

Context mechanism outcome configuration 17: **'need for competence**, **autonomy and relatedness'**

If participants create new knowledge and plan an implementations strategy (C), then they feel satisfaction, responsibility and stewardship (O), because this fulfils their need for competence (a sense of self-efficacy to achieve specific objectives) (M), autonomy (a feeling of being in control of one' s own behaviour) (M), and relatedness (a sense of connection to a larger group) (M).

*We also have the feeling of togetherness when we go to the literature and read and discuss guidelines, but then agree that it is not always possible to follow them in all situations. We then somehow deviate somewhat from evidence-based medicine, but only for the good of the patient. We then feel on equal footing with the specialists, we work almost guideline compliant, and that creates professional identity [translated from Swiss German] (5).*

*It is important that the group jointly works through the pdca cycle. And then the facilitator - it doesn't always have to be just the facilitator, it can also be a participant - should make sure that you measure how it is and then again later, when everyone is changing in a similar way. Basically, this is the task of the facilitator, but the group acts as a regulating element for the individual, so that everyone participates, in the development and in the change of behaviour. And when the group has made progress, it feels good [translated from Swiss German] (3).*

*And you're satisfied because you were involved, and so you can relate to the content of the innovation, and that's probably why it works so well; it was well done and I can identify myself with it...and support it...and you feel like you developed it or helped develop it [translated from Swiss German] (4a).*

Context mechanism outcome configuration 18: **'intention to change'**

If participants publicly announce their intention to change (C), then they are more likely to implement the change (O) because they and others in the group both think it is a good idea and they believe they can carry it through (M).

*And that gives me the feeling of significance and satisfaction in QCs, when I've done something like this. And when I also intend to use [new knowledge... the facilitator helps me if he carries me through the PDCA cycle. In this phase, the facilitator is very important, important for planning, because he helps me when I have to plan; that is, when the members or at least one of them has decided to change behaviour [translated from Swiss German] (2).*

#### *Repeating the process*

Context mechanism outcome configuration 19: 'testing new knowledge'

If participants validate and test new knowledge in a QC moderated by a skilled facilitator in a safe environment (C), then they feel safe to put that knowledge to use in everyday practice (O) because they have had the opportunity to practise and familiarise themselves with the innovation (M).

**Supplemental material 9 revised programme theory including illustrative quotes and complete list of supporting quotes**

*No data*

Context mechanism outcome configuration 20: **'gaining confidence in an innovation'**

If the group iteratively practices implementing and coping with an innovation (C), then they trust their own competence and turn the innovation into a habit (O) because successful outcomes build up confidence in their abilities (M).

*Is a sustainable change taking place? I think yes, but just, if I am completely honest, I have to tell you that I cannot always prove it. I just think that if it were documented, i.e. if the process in the QC were well documented, the change would be more sustainable. And when you see that it works, it is even more motivating and shows that the QC is a good thing [translated from Swiss German] (2).*

*But to recognise whether the process actually leads to a change in my own behaviour, I have to have an outside indicator that shows it to me, time and again. In principle, this means that I have to have an external objective assessment of what I am doing and whether anything is changing - and this helps to change my behaviour and to gain confidence in [translated from Swiss German] (1).*

*Right. Because that is the ultimate question: if I wanted to change something, then something should change and if nothing has changed, then it is somewhere on the way between wanting it and being stuck; but if I see that it starts working, it is very motivating and I continue [translated from Swiss German] (1).*

Context mechanism outcome configuration 21: '**'repetition priming and automaticity'**

If participants build a steady group and practice using QI tools (C), then they will successfully implement new knowledge into everyday practice (O) because successful responses increase with repetition: 'practice makes perfect' (M).

*I facilitate a QC there and notice that the motivation has rather increased over the years because the participants have realized what benefits they get from it. But in the beginning people were extremely critical and passive. [translated from Swiss German] (2).*

*That is totally exciting, when you see that the participants choose the same or similar topics, but they deal with them in a very different way and they go deeper into the topic [translated from Swiss German] (2).*

*It is a fact that you use the same techniques, over and over again, in terms of methodology - and don't even notice it...so, even if you don't notice it, it helps [to improve quality in PHC], that's for sure, that's absolutely right [translated from Swiss German](3).*

*But a lot of things are just as I have just described, and it is effectively the case that local autonomy is important for the implementation of innovations. The organisations must regard this as important, and the new knowledge must be available on a platform, so that it can be exchanged again and again, and looked at until it is established [translated from Swiss German] (3).*

*Well, I have had varying experiences; one is that you have to think in great detail and do the same procedure over and over again. But if, over a longer period of time, this same process has become established, and the thinking within the group becomes more similar, or they become, how should I put it, part of this, this, this basic cycle, then sometimes you can skip elements and make fairly rapid progress. It is a question of routine [translated from Swiss German] (1).*

**Supplemental material 9 revised programme theory including illustrative quotes and complete list of supporting quotes**

*I can also see that it takes a long time and that it takes two or three QC sessions before an improvement is achieved. Therefore, you have to keep addressing the problem and thus initiate a repetitive process [translated from Swiss German] (4b).*

### Results of the focus group sessions

#### *Focus group: summary of agreement / disagreement*

**Conference Workshop: April 24th**

Participants: 21

Quality Circles: what works for whom and why

Adrian Rohrbasser / Sharon Mickan / Janet Harris

I. Read the summary statements

II. Think about your own groups while considering the statements

III. Consider the ones that are different in your group

IV. Add your comments to the statements

1 Statement

If an invitation to a meeting with general information on QCs addressing GP needs is issued and a facilitator, preferably a GP, introduces QCs and explains how they work

then the meeting takes place and people become familiar with the theoretical background of the QC model

provided that the venue is close to the working place and time convenient and/or CME Points are offered.

Agree ☐

Disagree ☒ / unsure Why? Please explain below:

4 out of 21 disagree or are unsure

Too many invitations: invitation has to be personal (not further explained although facilitator asked)

Facilitator needs a reward because of "more work"

2) Statement

If a GP, as part of the team, is introduced into the group as a facilitator

then QCs are more relevant to the participants and a sense of ownership is created

**Supplemental material 9 revised programme theory including illustrative quotes and complete list of supporting quotes**

provided that the GP has completed 2 days' training in facilitating structured small group work

Agree ☐

Disagree ☒ /unsure Why? Please explain below:

5 out of 21 disagree or are unsure

Denmark: Flat democratic structure; all members have facilitator training and therefore they have an understanding of the role of a facilitator; role of facilitator rotates.

People feel safe when there is equality about facilitation

Swiss participant: the group needs a leader and training in leadership, continuous and updated training in leadership.

Germany: Facilitation needs qualification and training and financial compensation. Financial resources to train facilitators are important.

Facilitator should be a GP but other professionals may have the skills. Facilitators need respect and understanding for GP environment.

The leader must be accepted by the group – formal or informal -. Training in leadership should be part of GPs' training

Uncertain whether 2 days are enough: description of hours and content are needed.

Spain: content is more important than hours or days!

Talented Facilitators need to be able to motivate - clear leadership is required.

Shared facilitation is possible

Separate group leader and facilitator are also possible: separate group leader and separate facilitator during the meeting.

3) Statement

If objectives, both for the team as a whole and for individual team members are discussed, including the venue, duration and frequency of meetings, as well as individual tasks, such as writing the minutes then the group forms and develops a basic level of trust

provided that the facilitator introduces people to each other, opens discussions, clarifies and summarizes statements in a group arranged in a circle without barriers, such as screens.

**Supplemental material 9 revised programme theory including illustrative quotes and complete list of supporting quotes**

Additional context features: professional and administrative autonomy are needed as well as rooms and equipment.

Agree ☐

Disagree ☒ / unsure Why? Please explain below:

2 out of 21

4) Statement

If social norms are discussed, for instance punctuality and how people interact with each other

and if further tasks within the group are talked over

then group bonding develops and the level of trust increases

provided that professional and organisational autonomy are granted

Agree ☐

Disagree ☒ /unsure Why? Please explain below:

1 out of 21

5) Statement

If people can present their clinical cases and share their experiences

then people will be satisfied with the group process

provided that the facilitator ensures interactive responses and respects each member's contributions.

Additional context features: organizational barriers and excessive demands for the meetings have to be avoided.

Protected time is offered: the meeting takes place during working hours and participants are freed from clinical duties during the sessions.

Agree ☐

Disagree ☒ / unsure Why? Please explain below:

7 out of 21

It has to take place during working hours

Confidential atmosphere and mutual respect for each other

Sharing experiences is enough the beginning

**Supplemental  material 9 revised programme theory including illustrative quotes and complete list of supporting quotes**

After 2- or 3-years people become more expert in the process of QCs

People need a clear agenda (facilitator tried to clarify what that meant – could not be clearly explained)

Shared responsibility for the content is necessary; the whole group has to set the agenda and prepare the content

Each session is slightly different

Delegate ownership for different sessions (???)

Moderator has to ensure that the group works

You have to clarify roles and responsibilities

6) Statement

If the group discusses case reports and experiences

then people will participate actively and relate to the group

provided that the facilitator elicits open communication

Agree ☐

Disagree ☒ / unsure Why? Please explain below:

4 out of 21

Cases are accepted as common agendas

Safe environment and mutual respect for each other

Leader starts to show vulnerability and others will follow (leadership or facilitator = role model)

It is better to have big questions about new laws, policy and practice; a single case is not enough; the issue should affect all people

Sharing emotionally difficult cases: if difficult for one, then difficult for others: helping each other.

For instance, Critical Incidence feels like relief for others and is therefore seen as important.

Need to have mutual goal, how to define a purpose. Leaders and members have to define the goal.

You should negotiate the topic with all members

Needs of the group change over time

Democratic process

7) Statement

**Supplemental material 9 revised programme theory including illustrative quotes and complete list of supporting quotes**

If the group discusses difficult experiences that burden participants

then others will appreciate their professional role and group identity increases

provided that confidentiality is guaranteed within the group and granted even at an organizational level

Agree ☐

Disagree ☒ Why? Please explain below:

1 out of 21

8) Statement

If clinical cases are presented and different opinions discussed

then interactive learning and personal reflection on action take place

provided that the facilitator involves all QC members with an appropriate balance between comfort and challenge, depending on what level of trust the group has reached.

Agree ☐

Disagree ☒ Why? Please explain below:

1 out of 21

9) Statement

If the group contemplates clinical cases and mirrors their practice by looking at diagnostic patterns or prescription habits and discusses emerging topics in the light of evidence-based information

And/Or

If the group reflects on video consultations or results of patient satisfaction surveys

And/Or

If the group meets up with a local opinion leader and reflects on clinical scenarios

then people increase their understanding and gain insight into gaps in knowledge

provided that the facilitator establishes and maintains a learning environment and acts cautiously when addressing performance.

Additional context features: the management supports QCs to improve performance by allowing autonomy while setting expectations. Access to scientific knowledge, practice guidelines, quality indicators, a performance management system and an electronic medical record are provided to mirror current practice.

**Supplemental material 9 revised programme theory including illustrative quotes and complete list of supporting quotes**

Agree ☐

Disagree ☒ Why? Please explain below:

3 out of 21

It might increase

Need to look and analyse

Feedback is important to each other

Measurement and data triggers discussion

Data is an important tool

Need to learn about data / skills to analyse and interpret / trust data

Data is supported by AQA institute for discussion in QCs – it took a long time to build trust and confidence

After QC: people have to move towards agreement, consensus

10) Statement

If the facilitator uses different techniques to reflect practice, such as brain-storming, followed by contentious discussions and reaching a consensus, professional reprocessing of patient situations and role play, raising awareness of emotions, and purposeful use of local experts

then new knowledge is created

provided that the management accepts that acknowledging unstated beliefs about innovations is essential for creating new knowledge.

Additional context features: the management values QC contributions to the organization. Local adjustments of knowhow are allowed in order to customize knowledge and organizational pluralism is accepted.

Agree ☐

Disagree ☒ Why? Please explain below:

4 out of 21

Management: level above QCs

Change techniques to reflect gradually - group develops and learns different ways

You should stick to the same technique

**Supplemental  material 9 revised programme theory including illustrative quotes and complete list of supporting quotes**

Mature group will change ways of working more easily; choose technique to fit the problem

Check if right technique is used after the session

11) Statement

If participants in the group compare their experiences and current practice with each other and with evidence-based information using different facilitating techniques like contentious discussion and reaching a consensus

then new knowledge is validated and corroborated

provided that the management agrees on shared decisions when it comes to the use of new knowledge and accepts a certain degree of diversity in the organization.

Agree ☐

Disagree ☒ Why? Please explain below:

1 out of 21

12) Statement

If the facilitator helps the group or the individual to make a binding plan of what they want to change in the light of new knowledge

then participants make a commitment to change

provided that the management agrees on shared decisions when it comes to the use of that new knowledge.

Agree ☐

Disagree ☒ Why? Please explain below:

3 out of 21

Continuous reflection

As knowledge increases plans evolve

Plan also needs a follow up PDCA

Facilitate change

Binding plan – make participants aware of, engage, participate from start creates ownership

Nothing changes – need social, financial incentives to change

Long term commitment

**Supplemental material 9 revised programme theory including illustrative quotes and complete list of supporting quotes**

13) Statement

If the group discusses factors that hinder or foster implementation of new knowledge or change of practice and carefully evaluates adjustments to local conditions

then people use that new knowledge at their working place

provided that the management supports social processes that form and circulate knowledge and accepts a certain degree of diversity in the organization

Agree ☐

Disagree ☒ Why? Please explain below:

1 out of 21

14) Statement

If the group compiles detailed information about how to follow progressive goals and continuously evaluate new knowledge

then new knowledge is systematically used and evaluated in the working place

provided that the management supports social processes that form and circulate knowledge and assists in carrying out the tasks

Agree ☐

Disagree ☒ Why? Please explain below:

4 out of 21

Not everyone will follow

Doing a little bit better is always good

Small change is achievable

Group may need different targets

Need monitoring feedback

Reflections

Differences in funded and free QCs

Nothing about financial support

**Conference Workshop April 25th**

12 Participants

**Supplemental material 9 revised programme theory including illustrative quotes and complete list of supporting quotes**

Quality Circles: what works for whom and why

Adrian Rohrbasser / Sharon Mickan / Janet Harris

I. Read the summary statements

II. Think about your own groups while considering the statements

III. Consider the ones that are different in your group

IV. Add your comments to the statements

1) Statement

If an invitation to a meeting with general information on QCs addressing GP needs is issued and a facilitator, preferably a GP, introduces QCs and explains how they work

then the meeting takes place and people become familiar with the theoretical background of the QC model

provided that the venue is close to the working place and time convenient and/or CME Points are offered.

Agree ☐

Disagree x Why? Please explain below:

4 out of 12

Denmark: don't wend out invitations (what other ways of contacting and offering training – Facilitator tries to investigate – no answer) – no CME points should be offered

CME points are important, but not the most important reason to attend

A sent invitation is not enough – "it does not make sure that a person will attend"; link invitations to an existing system. Organisational structure makes the difference, for instance networks.

Use external motivator, for instance money to get there = trigger similar to CME points.

Need information to be informed about QC (facilitator asks for further explanations without success)

Convenient location is important

2) Statement

If a GP, as part of the team, is introduced into the group as a facilitator

then QCs are more relevant to the participants and a sense of ownership is created

provided that the GP has completed 2 days' training in facilitating structured small group work

**Supplemental material 9 revised programme theory including illustrative quotes and complete list of supporting quotes**

Agree ☐

Disagree 4 Why? Please explain below:

4 out of 12

Facilitators don’t have to be GPs

On order to receive CMEs, there has to be a moderator (=facilitator)

In Norway, training takes two years to become a facilitator and three years’ of GP facilitation (not confirmed by other members und not confirmed in the literature)

Bottom up system in QCs creates natural leaders

The facilitator has to be part of the group: flat hierarchical structure creates sense of ownership

Members of the group offers to be facilitator

Not all groups have a facilitator; all are equal (Denmark)

Another Danish participant: the group leader is the administrative secretary, teacher and facilitator comparable to the role of a CEO

In Denmark, all participants attend moderator courses and can lead....

3) Statement

If objectives, both for the team as a whole and for individual team members are discussed, including the venue, duration and frequency of meetings, as well as individual tasks, such as writing the minutes then the group forms and develops a basic level of trust

provided that the facilitator introduces people to each other, opens discussions, clarifies and summarizes statements in a group arranged in a circle without barriers, such as screens.

Additional context features: professional and administrative autonomy are needed as well as rooms and equipment.

Agree ☐

Disagree x Why? Please explain below:

5 out of 12 disagree

You should not have any barriers like screens at all, they are distracters and should be removed

Physical arrangement of people is important, there has to be enough space

Rearrange classroom to interaction

**Supplemental material 9 revised programme theory including illustrative quotes and complete list of supporting quotes**

4) Statement

If social norms are discussed, for instance punctuality and how people interact with each other

and if further tasks within the group are talked over

then group bonding develops and the level of trust increases

provided that professional and organisational autonomy are granted

Agree ☐

Disagree x Why? Please explain below:

3 out of 12

Start with clinical discussions before you set norms!

Talk about basic rules at the first meeting

Address these topics – like social norms – after the group has been together – need trust to have a discussion.

1 sheet of paper = rules for the group = pass out in the 1st QC – these rules can be changed; after 3 – 4 meetings set a discussion to confirm the rules

As group norms grow, social control increases

If you discuss which rules the group has, it improves trust.

5) Statement

If people can present their clinical cases and share their experiences

then people will be satisfied with the group process

provided that the facilitator ensures interactive responses and respects each member's contributions.

Additional context features: organizational barriers and excessive demands for the meetings have to be avoided.

Protected time is offered: the meeting takes place during working hours and participants are freed from clinical duties during the sessions.

Agree ☐

Disagree x Why? Please explain below:

8 out of 12 disagree

Meeting doesn't not necessarily have to take place during working hours, really not necessary

**Supplemental material 9 revised programme theory including illustrative quotes and complete list of supporting quotes**

It should be out of working hours evening or during lunch time

The group should decide on this

6) Statement

If the group discusses case reports and experiences

then people will participate actively and relate to the group

provided that the facilitator elicits open communication

Agree ☐

Disagree x Why? Please explain below:

1 out of 12

7) Statement

If the group discusses difficult experiences that burden participants

then others will appreciate their professional role and group identity increases

provided that confidentiality is guaranteed within the group and granted even at an organizational level

Agree ☐

Disagree x Why? Please explain below:

1 out of 12

8) Statement

If clinical cases are presented and different opinions discussed

then interactive learning and personal reflection on action take place

provided that the facilitator involves all QC members with an appropriate balance between comfort and challenge, depending on what level of trust the group has reached.

Agree ☐

Disagree x Why? Please explain below:

1 out of 12

9) Statement

If the group contemplates clinical cases and mirrors their practice by looking at diagnostic patterns or prescription habits and discusses emerging topics in the light of evidence-based information

**Supplemental  material 9 revised programme theory including illustrative quotes and complete list of supporting quotes**

And/Or

If the group reflects on video consultations or results of patient satisfaction surveys

And/Or

If the group meets up with a local opinion leader and reflects on clinical scenarios

then people increase their understanding and gain insight into gaps in knowledge

provided that the facilitator establishes and maintains a learning environment and acts cautiously when addressing performance.

Additional context features: the management supports QCs to improve performance by allowing autonomy while setting expectations. Access to scientific knowledge, practice guidelines, quality indicators, a performance management system and an electronic medical record are provided to mirror current practice.

Agree ☐

Disagree x Why? Please explain below:

3 out of 12

Management is the level above the GP level (explanation ADR) Wrong word? Authority was proposed, accountability, government

Denmark: GPs are the managers! (facilitator points out that these GPs play a different role then – as mangers – even if it is the same person: for instance – QC during working hours – managerial decision even though it is taken be a GP in the role of the manager...)

Danish participant: There are different levels above the GP: medical association – they teach, supervise and facilitate

There is a different impact if a local opinion leader leads the QC

When there is no support, the group manages itself

10) Statement

If the facilitator uses different techniques to reflect practice, such as brain-storming, followed by contentious discussions and reaching a consensus, professional reprocessing of patient situations and role play, raising awareness of emotions, and purposeful use of local experts

**Supplemental  material 9 revised programme theory including illustrative quotes and complete list of supporting quotes**

then new knowledge is created

provided that the management accepts that acknowledging unstated beliefs about innovations is essential for creating new knowledge.

Additional context features:  the management values QC contributions to the organization.  Local adjustments of knowhow are allowed in order to customize knowledge and organizational pluralism is accepted.

Agree   x

Disagree ☐       Why? Please explain below:

11)       Statement

If   participants in the group compare their experiences and current practice with each other and with evidence-based information using different facilitating techniques like contentious discussion and reaching a consensus

then new knowledge is validated and corroborated

provided that the management agrees on shared decisions when it comes to the use of new knowledge and accepts a certain degree of diversity in the organization.

Agree   x

Disagre e          Why? Please explain below:

12)       Statement

If the facilitator helps the group or the individual to make a binding plan of what they want to change in the light of new knowledge

then participants make a commitment to change

provided that the management agrees on shared decisions when it comes to the use of that new knowledge.

Agree   x

Disagree ☐       Why? Please explain below:

**Supplemental material 9 revised programme theory including illustrative quotes and complete list of supporting quotes**

13) Statement

If the group discusses factors that hinder or foster implementation of new knowledge or change of practice and carefully evaluates adjustments to local conditions

then people use that new knowledge at their working place

provided that the management supports social processes that form and circulate knowledge and accepts a certain degree of diversity in the organization

Agree x

Disagree ☐ Why? Please explain below:

14) Statement

If the group compiles detailed information about how to follow progressive goals and continuously evaluate new knowledge

then new knowledge is systematically used and evaluated in the working place

provided that the management supports social processes that form and circulate knowledge and assists in carrying out the tasks

Agree ☐

Disagree x Why? Please explain below:

3 out of 12

Denmark: the results (data) should not be discussed in the group

Too optimistic, the group won’t change as much as we (?) hope

Clarification of compile: minimal documentation about how to proceed (?)

Pre survey in each QC for each practice; post survey and qualitative data could show change

#### *Workshops in Fischingen: Summary of participants’ ideas about mechanisms of the programme theory*

1. Summary statement: participation in the meeting is accepted when General Practitioners’ needs are addressed and logistical barriers to attending are identified and tackled during the initial phase of the program.

Process outcome: People are familiar with the theoretical background of QC model

**Supplemental material 9 revised programme theory including illustrative quotes and complete list of supporting quotes**

Activities: invitation to an informative meeting with general information on QCs, addressing GP needs. A facilitator, preferably a GP, introduces QCs and explains how they work.

Context: Venue is close to working place and time convenient; CME Points are offered.

**Mechanisms:** "Reasoning"

*M1 People feel they have an important stake so participate*

*M2 People are engaged in their work, want to learn autonomously and therefore decide to participate*

*M3 People think the meeting will lead to self-satisfaction and reward.*

*M4 People seek new challenges in new areas and expect that these will increase their competence and help them relate to colleagues.*

*M5 Social pressure makes them participate because of membership in a network and the associated responsibilities of joining QCs.*

*M6 Health insurance companies identify GPs using resources inadequately and make them participate.*

2. Summary statement: involving GPs as facilitators as part of the team in QCs leads to interventions being more relevant to participants' needs and creates a sense of ownership by those delivering and receiving the intervention.

Process outcome: Group is facilitated in a professional way

Activities: A GP who is trained in facilitating structured small group work is introduced into the group

Context: A GP has completed 2 days' training in facilitating structured small group work

**Mechanisms:** "Reasoning"

*M1 since it is a GP who facilitates the program among other GPs, it becomes more relevant to participants and creates a sense of ownership.*

*M2 a facilitator who is a GP gains more credibility - the facilitator is perceived by the participants to be 'one of us'*

3. Summary statement: group bonding and a basic level of trust develop when the group autonomously decides on the structure of its meetings with the help of a trained facilitator in an open atmosphere without barriers.

Process outcome: Group Forming and basic level of trust

Activities: The facilitator introduces people to each other, opens discussions, clarifies and summarizes statements. Objectives, both for the team as a whole and for individual team members are discussed. Venue, duration and frequency of meetings are discussed, as well as individual tasks like writing the minutes.

Context: At an organizational level: professional and administrative autonomy are needed as well as rooms and equipment.

At a group level: group is arranged in a circle without barriers, such as screens

**Mechanisms:** "Reasoning"

*M1 The group members feel familiar with what they are going to cover and are able to participate actively.*

4. Summary statement: the social structure and the level of trust grow when the group autonomously decides on social norms like punctuality and feedback culture, with the help of a trained facilitator, provided that they are discussed and agreed upon in an open atmosphere without barriers.

Process outcome: Group Norming and increased level of trust

Activities: The facilitator opens discussions in the group, clarifies and summarizes statements as they discuss the ground rules of how people interact with each other. People are introduced to each other at a deeper level and resolve possible differences. Social norms like punctuality and further tasks within the group are talked over.

Context: At an organizational level: professional and administrative autonomy are needed as well as rooms and equipment.

At a group level: group is arranged in a circle without barriers, such as screens

**Mechanisms:** "Reasoning"

*M1 The group members feel familiar with the group and are able to relate to each other.*

5. Summary statement: Presentation of own clinical cases in a well facilitated group increases the feeling of reassurance and being acknowledged in the group, and increases self-esteem. Protected time, no excessive demands and no organizational barriers are prerequisites of this process.

Process outcome: satisfaction with the group process.

Activities: People present their clinical cases and share their experiences. The facilitator ensures interactive responses and respects each member's contributions.

**Supplemental material 9 revised programme theory including illustrative quotes and complete list of supporting quotes**

Context: There are neither organizational barriers nor excessive demands for the meetings. Protected time is provided: the meeting takes place during working hours and participants are freed from clinical duties during the sessions.

**Mechanisms:** "Reasoning"

*M1 By reconfirming their practice among colleagues, the feeling of security and predictability is increased.*

*M2 A sense of affiliation to the group and being acknowledged grows when common experiences are shared.*

*M3 When colleagues actively listen to experiences, it increases self-esteem.*

6. Summary statement: discussion of case reports makes the group active and creates a supportive and understanding culture among participants.

Process outcome: Active participation and relatedness to the group

Activities: the group discusses case reports and experiences with help of a facilitator, who elicits open communication.

**Mechanisms:** "Reasoning"

*M1 Family physicians are engaged in their work and focus on relevant, practical knowledge that is of immediate use to them.*

*M2 Common experience creates mutual understanding and gives a sense of collegiality.*

7. Summary statement: discussion of difficult experiences and exchange of emotional responses provides recognition of professional roles and increases group cohesion if confidentiality is guaranteed at a group and organizational level.

Process outcome: Appreciation of professional role and increase in group identity.

Activities: the group discusses difficult experiences that burden participants. The facilitator leads the discussion and supports narrators through active listening techniques.

Context: Confidentiality is guaranteed and granted, even at an organizational level

**Mechanisms:** "Reasoning"

*M1 Family physicians are engaged in their work and focus on relevant, practical knowledge that is of immediate use to them.*

*M2 Common experience creates mutual understanding and gives a sense of collegiality.*

**Supplemental material 9 revised programme theory including illustrative quotes and complete list of supporting quotes**

8. Summary statement: case discussions as a basis of challenging each other's position enable the group to reflect on their practice and to learn from each other in a cooperative atmosphere of mutual understanding.

Process outcome: Interactive learning and personal reflection on action

Activities: clinical cases are presented and different opinions discussed. The facilitator involves all QC members with an appropriate balance between comfort and challenge, depending on what level of trust the group has reached.

**Mechanisms:** "Reasoning"

*M1 Previous knowledge is activated through case discussions.*

*M2 The group supports and rewards exploratory behaviour by giving the feeling of competency, which enables participants to describe what they actually do.*

*M3 People are motivated to imitate those peers who are more competent and then receive positive feedback.*

9. Summary statement: when facilitators use different prompting techniques, they allow the group to reflect on their practice and acknowledge gaps in knowledge. The facilitator cautiously addresses performance while the management provides appropriate and trustworthy data.

Process outcome: Understanding and insight into gaps of knowledge

Activities: The group contemplates clinical cases and mirrors their practice by looking at diagnostic patterns or prescription habits. Emerging topics are discussed in the light of evidence-based information. The facilitator establishes and maintains a learning environment and acts cautiously when addressing performance.

The group reflects on video consultations or results of patient satisfaction surveys.

The group meets up with a local opinion leader and reflects on clinical scenarios.

Context: the management supports QCs to improve performance by allowing autonomy while setting expectations. Access to scientific knowledge, practice guidelines, quality indicators, a performance management system and an electronic medical record are provided to mirror current practice.

**Mechanisms:** "Reasoning"

**Supplemental material 9 revised programme theory including illustrative quotes and complete list of supporting quotes**

*M1 Previous knowledge is activated through case discussions.*

*M2 The group supports and rewards exploratory behaviour by giving the feeling of competency, which enables participants to describe what they actually do.*

*M3 People are motivated to imitate peers who are more competent and receive positive feedback.*

*M4 Critical reflection on experience and practice enables practitioners to identify learning needs.*

*M5 Addressing performance may cause anxiety and frustration among participants.*

10. Summary statement: The group creates new knowledge when they mirror and reflect current practice in a well facilitated learning environment, given that the management values their contributions.

Process outcome: Creation of new knowledge

Activities: The facilitator uses different techniques to reflect practice, such as brain-storming, followed by contentious discussions and reaching a consensus, professional reprocessing of patient situations and role play, raising awareness of emotions, and purposeful use of local experts.

Context: Management accepts that acknowledging unstated beliefs about innovations is essential for creating new knowledge and values QCs' contributions to the organization. Local adjustments of knowhow lead to customized knowledge and fit into organizational pluralism.

**Mechanisms:** "Reasoning"

*M1 Previous knowledge is activated through discussions of current practice*

*M2 People are motivated to imitate peers who are more competent and receive positive feedback.*

*M3 Critical reflection on experience and practice enables practitioners to identify new knowledge.*

*M4 Family physicians are engaged in their work and focus on relevant, practical knowledge that is of immediate use to them.*

11. Summary statement: Provided that the management acknowledges and accepts QC contributions, the group appraises and modifies new knowledge when they compare their experiences with each other and with evidence-based information.

Process outcome: Validation of new knowledge

Activities: Participants of the group compare their experiences and current practice with each other and with evidence-based information, using different facilitating techniques, such as contentious discussion and reaching a consensus.

**Supplemental material 9 revised programme theory including illustrative quotes and complete list of supporting quotes**

Context: the management agrees on shared decisions when it comes to the use of new knowledge and accepts a certain degree of diversity in the organization.

**Mechanisms:** "Reasoning"

*M1 The group supports and rewards exploratory behavior by giving the feeling of competency, which enables participants to test new knowledge.*

*M2 Analytical reflection on experience and practice enables practitioners to critically appraise new knowledge.*

12. Summary statement: When the group or individuals develop their plan of change it becomes a binding arrangement, provided that the management values their ideas regarding the use of new knowledge.

Process outcome: Commitment to change

Activities: the facilitator helps the group or the individual to make a binding plan of what they want to change in the light of new knowledge

Context: the management agrees on shared decisions when it comes to the use of new knowledge

**Mechanisms:** "Reasoning"

*M1 New knowledge that has been acquired in a learning environment influences individual perception concerning the perceived risks and benefits of making a change and allows a change of attitude and commitment to change (Health Belief Model)*

*M2 Individuals of a group take into account the social norms and practices of their peers when they take in knowledge and implement it (theory of reasoned action)*

13. Summary statement: the use of knowledge or skills in the working place is fostered when the group is endorsed by the management in making local adjustments and in removing barriers to innovations.

Process outcome: Use of knowledge in the working place

Activities: The group discusses factors that hinder or foster implementation of new knowledge or change of practice and carefully evaluates adjustments to local conditions.

Context: The management supports social processes that form and circulate knowledge and accepts a certain degree of diversity in the organization.

**Mechanisms:** "Reasoning"

**Supplemental material 9 revised programme theory including illustrative quotes and complete list of supporting quotes**

*M1 People feel that they are in control of and empowered by the process.*

*M2 As people have developed new knowledge or skills themselves, they have confidence in their ability to take action.*

*M3 The commitment to change creates a sense of urgency to use new knowledge or skills.*

*M4 People believe that new knowledge or skills they developed themselves are relevant and important to them.*

14. Summary statement: people can put new knowledge or skills into systematic use when they plan progressive goals they can follow under the guidance of a facilitator, in agreement with the management.

Process outcome: Systematic use of new knowledge in the working place and re-evaluation

Activities: Under the watch of the facilitator, the group compiles detailed information about how to follow progressive goals and continuously evaluate new knowledge.

Context: The management supports social processes that form and circulate knowledge and assists in carrying out the tasks.

**Mechanisms:** "Reasoning"

*M1 Social support and guidance in using new knowledge increases the ability to take action*

*M2 Confidence in the group's ability to take action increases when they use progressive and iterative goals*

*M3 Anxiety is reduced when the group demonstrates desired behaviour.*
