## Supplemental material 10 for "How and why do Quality Circles work for General Practitioners - a realist approach"

**Supplemental material 10 consolidated programme theory, including illustrative quotes and complete list of supporting quotes**

### 1. Preconditions

a) 'Need for autonomy and obligation'

If the administration at national level or at the level of health insurance companies entrusts GPs with QI and autonomy (so they can decide how to implement it) (C), then GPs might participate in QCs (O) because they feel they can take on the responsibility and make a difference (M).

*So, we got more money, but it was for the government no value for money … well … extra value for the money. The only obligation was to participate in the local QC which had to gather four times a year, and you had to participate at … at least two of them every year to keep your accreditation. But you should have an obligation to improve your quality in your practice (1).*

New CMO configuration: 'Being embedded in a system of QI'

If QCs are embedded in a QI system (an organisation that negotiates and signs contracts with governmental bodies or health insurance companies, trains and supervises facilitators, provides courses on QI in PHC, and easily accessed educational material, timely data on practice performance and protected time and space) (C), then participants will take on responsibility and work in a purposeful way (O) because they feel supported, empowered, and capable of meeting expectations (M).

*… embedding QCs in a system … organising during working time is one ... training facilitators is another one, in a continuous way and honouring in one way or another, maybe financially, especially for the extra hours and the extra work they put into it, and ... offering GPs the possibility of easily gathering data about their own practice ... in a much shorter time, getting feedback on your practice from a national level and getting it in a systematic way ... brought into the peer review would be a good way (1).*

*…so, you know the evaluation is mainly to help the person who is organising; the evaluation is really for the tutor, because they [the organisation] are structuring and organising the meetings and it is really seen as a support process … (2).*

*I think our problem is at the level of the organisational context. We don't get any support, we don't have protected time, we don't get any help to ... implement new things and do quality improvement … administrative support does not exist …and we have too much to do … too many patients a day (5).*

b) 'Feeling they have a say'

If an organisation, (e.g. a physician network organisation) has a decentralised *policy that* encourages use of local knowledge (C), then the QC takes on tasks (O) because members feel that they have a say in QI in their practice (M).

*No additional data.*

c) 'Participants know what to expect'

If the introductory workshop teaches the principles of QI in PHC and illustrates how QCs work (C), then potential members may be more willing to join QCs (O) because they know what to expect and feel that they can meet expectations (M).

*so ... some of them will work well [depending on] whether there is somebody who is inspired and wants to take the lead and knows something about peer review, but most of them are just nice meetings to see colleagues and ... have somebody give a presentation or have some food and drink. So, you should really teach them first! (1).*

**Supplemental material 10 consolidated programme theory, including illustrative quotes and complete list of supporting quotes**

| |
|---|
| *…because they are paid for it and but ... there has not been enough understanding in the medical corps to ... to do it and it usually comes on top of all the other ... (3).* |
| **2. Establishing the group** |
| a) ‘Sharing similar needs’<br><br>If the administration at the organisational level of QCs provides support (i.e. in training facilitators, data gathering, provision of evidence-based information), protects time and space and offers CME points and small financial incentives to QC participants (C), then the latter will meet in groups to exchange ideas (O) because GPs prefer learning in QCs (M); support generates positive expectations among participants (M) and GPs believe that QC meetings with their peers will be useful (M).<br><br>*And I think that ... the other thing that is important to the group is the CME / CPD points that they get and the funding from the government to attend meetings. That is all supporting the meetings as well (2).* |
| b) ‘Need for relatedness’<br><br>If a steady group of members engages in socially enjoyable contact, led by a skilled facilitator who, e.g. introduces people to each other, opens discussions, and clarifies and summarises statements (C), then group members will get to know each other and decide on rules that they are willing to follow, building a safe environment based on trust (O) because members want to be among and to interact with equals (M).<br><br>*…and we do that at dinner time so we can have some food together; we work, have dinner and we can enjoy food at the same time (4).*<br><br>*… but it became clear that we started to get to know each other and each other’s sensibilities and to dare to tell about how we handle things and we learnt how to handle each other in a respectful way. Now we have to see how it continues (1).* |
| c) ‘Need for autonomy and control’<br><br>If the group chooses its own topics and facilitator (C), then its members will feel they own the QC (O) because their need for autonomy - a feeling of being in control of their own behaviour - is satisfied (M).<br><br>*…exactly big autonomy, the groups decide, there is no pressure from the political system and there is no pressure from anybody and that is why this system is so successful – the doctors can choose (2).* |
| d) ‘Size of the group affects communication’<br><br>If group size exceeds 15 (C), then interaction among group participants decreases (O) because participants cannot keep up with each other and follow all conversations (M).<br><br>*For instance, if ... I think …15 people is too many … I think eight is enough and ... the stress increases if there are more … the smaller the group is, the better the trust and talking (4).* |

**Supplemental material 10 consolidated programme theory, including illustrative quotes and complete list of supporting quotes**

e) 'Variety of characters stimulates reflection – cognitive dissonance'

If members of the group have individual character traits and describe different professional experiences but accept each other's views (C), then they can learn from each other (O) because individual attitudes and behaviours will contrast with the knowledge of their peers and cause cognitive dissonance (a negative emotional state triggered by conflicting perceptions) that makes them reflect on their way of working (M).

*…. because you can learn from [other] people with more experience, ... you have a [another] way of thinking and [another] way of talking about stuff, situations, that are different I think, so I think it is about different knowledge (4).*

f) 'strong cognitive dissonance threatens self-image'

If individuals feel too strong a cognitive dissonance when integrating new knowledge (C), then they may disrupt group dynamics and halt the QC process (O) because their self-image is threatened and they fear losing their professional identity (M).

*Yes, we do, yeah, we have doctors who are ... difficult in the group, yes, and they are difficult because they have very firm views and they spend very little evidence on reality. Then it is very important that you have good group leaders and leadership … It is very few … you know trying to sabotage the group … and they don't tend to change behaviour (2).*

### 3. Learning environment

a) 'Feeling safe and not vulnerable'

If participants trust each other (C), then they can describe how they work and admit what they do not know (O), because they feel safe rather than vulnerable (M).

*…she told me, you know, one of the things I learnt from you, one of the things I experienced from you is that … opening up with difficult cases and showing that you don't know everything, is showing that you are vulnerable and not knowing what do with it …you build up trust because if you dare to do this, it gives us the confidence that we also can do that … (1)*

*We know each other very well, so I don't think anybody gets angry about this… and nobody ... gets emotionally the wrong way… if you understand what I mean (5).*

b) 'Need for competence and self-actualisation'

If the facilitator supports participants and encourages them to tell their stories and share their experiences in a safe environment, e.g. by encouraging interactive responses, through discussions and by summarising statements (C), then participants will be involved and share their positive experiences and failures (O) because they want to improve their professional competence (M), gain professional confidence (M) and fulfil their professional potential (M).

*… and the fact that you can explain it to the others makes you realise that ... you have a bit anxiety about it and all the others tell you that this ok – not just because they want to comfort you … then you realise that you became nervous about something very quickly … even if you did something good after all… the group at this moment is very … a peaceful place and a good way of being with yourself and your own way of practising and it increases your self-esteem as well (4).*

*But sometimes it is about our problems ... our professional life … about our patient, about some case … diagnostics or prescriptions (5).*

**Supplemental material 10 consolidated programme theory, including illustrative quotes and complete list of supporting quotes**

c) 'Previous knowledge is activated'

If participants exchange case stories and experiences while actively listening to each other in the presence of a skilled facilitator in a safe environment (C), then they will share their knowledge by telling their own relevant stories (O) because the process activates knowledge they already possess (M).

*It does satisfy us when we can discuss our own work and our own cases, and we feel closer in the group when we stimulate each other's thinking (4).*

d) 'Immediate relevance for the practice'

If QCs use the technique of experience-based learning (C), then knowledge becomes more relevant to GPs (O) because it relates to their everyday work and they can use it immediately (M).

*... a lot of the doctors will start with a clinical case, but then come to an overview and then discussions and the next step is organising the GP surgery for that – it is quick wins (3).*

e) 'Cognitive dissonance'

If participants discuss and reflect on their work processes (e.g. based on trustworthy data or personal experiences) during a professionally facilitated exchange of positive experiences or failures (C), then they discover knowledge gaps and identify learning needs and relevant topics (O) because their own attitudes and behaviours may differ from their peers', creating cognitive dissonance that makes them reconsider their own way of working (M).

*When for example a GP ... in a gr … group is saying that he does a particular thing that is purely not right, not evidence-based or in fact is wrong, then the group are very good … I think because they know each other… they do not agree with the doctor but they actually discuss it in the group and a few other doctors say what they would do which is usually different and they usually say ' you may consider this as a different way of doing it because if you do it your way, this is what I find happens…' and there is never an issue where somebody needs to feel bad, but they know that whatever they are currently doing is not what the others would (2).*

f) 'Social learning'

If the facilitator uses purposeful didactic techniques (e.g., brainstorming, contentious or consensus discussions, or role play) to keep the group active and to reward exploratory behaviour during reflection on the work process (C), then the group will create a learning environment that promotes the exchange of knowledge (O) because learning is a cognitive process in which participants observe and imitate their peers' behaviour to gain social approval (M).

*So, I think that the more experienced GPs bring in their cases into the groups and they discuss their experiences within the groups and I think this is very powerful for the group and the younger GPs bring in … they have the latest evidence in their head and the guidelines and they bring it in .., and the mix of managing the patient with the evidence and the guidelines and the practical bit from the older GP who has the experience, I think this is really the powerful bit in the group and … and this is where the learning really occurs (2).*

*Yes, ... in the beginning we thought this (sharing data) had to be in pairs or triplets because we thought that people were not willing to share, but that was quite wrong. They love to share! (3).*

### 4. <u>Adapting, creating and testing new knowledge</u>

a) 'Positive interdependence between the administration at the national level and GPs'

If the administration at national level requires continuous QC activities (C), then QCs will negotiate priorities and design creative solutions (O) because the tension between autonomy and obligation spurs the group to act and negotiate together to reach a common goal (M).

*It may be important for the emerging of QCs, that it becomes a mandatory thing [QI] and, after all, we have the same goals [as the health insurance companies] (4).*

b) ‘Threat to professional autonomy’

If GPs feel that the QC programme is only a top-down managerial intervention to reduce costs (C), then they will not be motivated and will not participate (O) because they feel unsafe and fear they lack autonomy in their clinical role (M).

*…no there are no demands, … that wouldn’t help, there can be wishes, but we decide how we do it… it wouldn’t work otherwise (3).*

c) ‘Positive interdependence among group members’

If participants maintain a learning environment based on trust that promotes the exchange of knowledge, assisted by facilitators who use professional techniques (e.g. contentious discussion, reaching consensus, and role play) (C), then participants will adapt and generate new knowledge for local use (O) because they see themselves as similar, and so act and negotiate cooperatively to achieve a common goal (M).

*I think that a group … cannot just be presented with things like, ‘here is the evidence, take it or leave it and goodbye’ and I don’t think that works. I think that people need to ... participate in the learning and they have to show what they are currently doing, whether it is the correct thing or not; it needs to be discussed and adjusted and shared within the group (2).*

d) ‘Identifying and removing barriers to change’

If participants, supported by skilled facilitators, address barriers to change (C), then they are more likely to implement the innovation (O), because participants help each other to develop strategies to identify and overcome these barriers (M).

*And I think you have to have guidelines that are workable for doctors who are, you know, seeing 30 to 40 people every day and, if they want to implement change for the better, they have to be feasible and practical and I think the only way to do that is to consider what they are currently doing. And what the barriers are to new care (2).*

e) ‘Need for competence, autonomy and relatedness’

If participants create new knowledge and plan an implementation strategy (C), then they feel satisfaction, responsibility and stewardship (O) because this fulfils their need for competence (being able to achieve specific objectives) (M), autonomy (a feeling of being in control of their own behaviour) (M) and relatedness (a sense of connection to a larger group) (M).

*No data*

f) ‘Intention to change’

If participants publicly announce that they intend to change (C), then they are more likely to implement the change (O) because they and others in the group all think it is a good idea and believe they can carry it through (M).

**Supplemental material 10 consolidated programme theory, including illustrative quotes and complete list of supporting quotes**

| |
|---|
| *… and I think that is the opportunity to state it [intention to change] … not everybody participates in that ... but ... most people do … and they'd say look this is what I learned, this is new for me, this is what I am … going to change in my practice (2).* |
| g) 'Testing new knowledge'<br>If participants validate and test new knowledge in a QC, moderated by a skilled facilitator, in a safe environment (C), then they feel confident putting that knowledge to use in everyday practice (O) because they have had the opportunity to practise and familiarise themselves with the innovation (M).<br>*… and I think that the idea of a quality circle meeting trying make changes dramatically is not practical. I think doctors need to look at ideas and look at the practical parts to see what they can do and change slowly over time (2).* |
| **5. Repeating the process** |
| a) 'Gaining confidence in an innovation'<br>If the group repeatedly practises implementing and adjusting to an innovation (C), then they trust their own competence and turn the innovation into a habit (O) because successful outcomes increase their confidence in their abilities (M).<br>*... then we meet again after four months and usually the … their quality improvement project ... didn't really happen or just a little bit, and we discuss the reasons for that and how we could amend that, etc. etc. (3).* |
| b) 'Repetition priming and automaticity'<br>If participants build a regular group and practise using QI tools (C), then they will successfully implement new knowledge into everyday practice (O) because responses improve with repetition: 'practice makes perfect' (M).<br>*… but the QC is really a double thing. It is about a theme but it is also about quality improvement. And the aim and goal is that they find it so rewarding that they use this this technique again and again … in their own surgeries and in their own groups (3).* |

**Supplemental material 10 consolidated programme theory, including illustrative quotes and complete list of supporting quotes**

### *Participants*

(1) GP in a rural practice, teacher at the University of Ghent (GP from Belgium)
(2) GP in a rural practice, small group educator for 18 years (GP from Ireland)
(3) Certified facilitator in GP vocational training, active in quality improvement and patient safety (GP from Norway).
(4) GP working in an urban area, facilitating a QC, researcher (GP in training from France)
(5) GP in a rural practice, teacher for GP vocational training (GP from Croatia).

Additional interviews: consolidation of the programme theory

### *Preconditions*

'Need for autonomy and obligation'

If the administration at the national level or at the level of health insurance companies entrusts GPs with QI and autonomy (puts them in control of how to do it) (C), then GPs may consider participating in QCs (O) because they feel they can take on the responsibility and make a difference (M).

*…we had quite a lot of criticism on the whole system because we did not feel it would really enhance quality and it was just used as a way of getting more money to the doctors without guarantees that quality would be enhanced, which is when we look back 25 years later, is exactly what happened. So, we got more money but it was for the government no value for money well extra value for the money. The only obligation was to participate in the local QC which had to gather four times a year, and you had to participate at ….at least two of them every year to keep your accreditation. But you should have an obligation to improve Your quality in your practice (1).*

*…., the only thing that is happening is at the national level the one who is responsible for the QC has to fill in after every QC who has been there and what was the subject of the meeting ... exactly there are no demands (1).*

*It may be important for the emerging of QCs, that it becomes a mandatory thing (4).*

*We have as an obligation in contracts with our insurance to have …… ... peer groups…… then ... I don't know how many times we should meet, actually.  But we don't have or get much money out of this (5)*

'Feeling of having a say'

If an organisation, (e.g., a physician network organisation) has a decentralised policy that encourages use of local knowledge (C), then the QC takes on tasks (O) because members feel that they have a say in QI in their practice (M).

*No data but confirming comments.*

'Participants know what to expect'

If the introductory workshop teaches the principles of QI in PHC and the workings of QCs (social persuasion) (C), this will increase the motivation of future participants to join QCs (O) because they learn what to expect and may feel that they are capable of meeting expectations (M).

*…they (QCs) are free to choose to what group they participate without any regulation and without any support of what is happening there without any control of what is happening there so ... some of them will work well (depending) whether there is somebody who is inspired and wants to take the lead and know something about peer review but most of them are just nice meetings to see colleagues and ... have somebody have a presentation or drink something and food. So, you should really teach them first! (1).*

*They get the knowledge about that from … … … the tutors, when they meet at these national workshops, of which there are three, they exchange ideas on useful quality tools and ways to use these tools among the groups and among the participants (2).*

*…because they paid for it and but ... there has not been enough understanding in the medical corps to ... to do it and usually comes on top of all the other …. (3).*

‘Quality Circles should be embedded in a system’

If QCs are embedded in a QI system (an organisation that negotiates and signs contracts with governmental bodies or health insurance companies, trains and supervises facilitators, provides courses on QI in PHC, and easy to access educational material, timely data on practice performance, and protected time and space) (C), then participants will take on responsibility and work in a purposeful way (O) because they feel supported, empowered, and capable of meeting expectations (M).

*But ... what did not happen is that the system of local QCs was really embedded in a movement or a way that would support people who participate that would make sure that people who took the lead really would support the facilitator the right way (1).*

*…by making a plan I mean having enough support on the content level which is there but also at the organisational level m... making it possible (to support facilitators) I do believe that the facilitator is very important (1)*

*…, the facilitator is the …at the start we …had some facilitator training a... 20 years ago, for some of the people who were interested but then that stopped because it was not financed by the government, and not supported anymore, a... and now for about 15 years there has not been a good generic facilitator training for those people who want to take on responsibility. And the ones who do that, it will be in their spare time they will not be paid for doing that e awarded in another way (1).*

*I can only tell that in our university in Ghent, that is one of the eight universities in Belgium, we try to learn (teach) the students during the last year, to work in peer review groups and then in the continuous education, the vocational training, they have to meet every two weeks, in groups of fifteen, so in the training, this tradition is established and there you have experienced facilitators being there to support these groups. But once they leave the training, and they start working as a GP, mmm this facilitating stops and they have to look for their own peer review groups and what is often happening, is that they cluster together, and make ….they already know each other and they build a new group with those people who started in the same region at the same time. ... and sometimes those are the most interested and the most interesting groups and they do really nice things, but of older doctors, we really don’t see that …that tradition (1).*

*I think peer review groups could be helpful in preventing burnout and finding on a local level way of cooperating to handle this problem of too much work ... even there if it is not supported or organised in a smart way from up, I think we will miss these chances (1).*

**Supplemental material 10 consolidated programme theory, including illustrative quotes and complete list of supporting quotes**

*…(embedding QCs in a system)…mmm ... organising during working time is one, ... training facilitators is another one, in a continuous way in and honouring in one or another way, maybe financially, especially for the extra hours and the extra work they put into it, and ... offering in the best way , offering GPs the possibility of easily gathering data of their own practice and being able to discuss that with their peers and colleagues would be the best if not, ... having in a much shorter time getting feedback on your practice from a national level and getting it in a systematic way ... brought into the peer review would be a good way (1).*

*It was the college that was in charge of the assessment to check the quality of the education because the Irish college of GPs has always been in control of …. of the quality and standards of education. But I think that is a good thing because I think that if your government spends money for an education system then it has to deliver what is relevant for a doctor working in primary cate at the moment. In the assessments, they try to see who is attending and how often and how big the groups should be what kind of educational material is covered and …. the three national workshops that we have and funded by the HSE executive we have the …. have to approve the programme and the teaching and how they deal with the groups (2).*

*The evaluation ..and usually there is a supportive evaluation so I'd have ..the year before that I had ... people that a group of doctors and you have two doctors who are familiar with this small group work and they come and visit an area and they'd sit in these groups and they talk how you can approve and it is mainly a support for the tutor I think because you have to look at what you are doing and you also get feedback from three people who are not usually attending your small group meetings. It is usually a very supportive structure and if they feel that it is something that is not appropriate or something you should change again, they actually there is an opportunity to do that as well. This is usually not seen as a negative process as far to my knowledge (2).*

*….. ... so you know the evaluation is mainly to help the person who is organising; the evaluation is really for the tutor, because they are structuring and organising the meetings and it is really would be seen as a support process …really it… it ..unless there are big problems within that group and if there are big problems in that group you have the opportunity to discuss them with the team who is coming and actually very often you can actually clarify or solve problems that are occurring within the group. …..(interviewee moves through the room – inaudible) …and be quite supportive you know and most of us see this positive So but it is a lot of work when I had a team visiting me I had to write a report and have all the names of the GPs attending, I had to have the structure of the group clarified and show what curriculum we have covered the last number of years ... and discuss how the curriculum was selected and about the needs assessment and you also highlight how educational sessions are evaluated you do carry out evaluations on the teaching you are doing (2).*

*…. and now our association tried to talk with our minister of health and the director of health insurance about we want to …implement I quality indicators in our everyday work ….in our electronic medical records. so, we tried to talk about that…. but nobody really heard us. … And unfortunately, …. we have only support from the association of GPs and a little support from university, but from university every support was only words…it was not anything substantial (5).*

*… and then the next step will be ... ... talk with health insurance so they give us more money so we can buy some new equipment for our practices so we can work more quality oriented and that we can think about quality (5).*

*I think our problem is at the level of the organisational context. We don't get any support, we don't have protected time, we don't get any help to ... implement something new and do quality improvement from the government… administrative support does not exist. ….and we have too much to do …. too many patients a day (5).*

*the QCs have become important at the university like the seventh and eighth year at the university ... when we do the specialisation about the GP or family medicine… but this is not very usual or common it is not nationally organised (4.)*

**Supplemental material 10 consolidated programme theory, including illustrative quotes and complete list of supporting quotes**

### *Establishing the group*

'Sharing similar needs'

If the administration at the organisational level of QCs provides administrative support (i.e. training of facilitators, data gathering and provision of evidence-based information), protected time and space, CME points, and small financial incentives to QC participants (C), then they will meet in groups to exchange ideas (O) because QCs are the preferred learning style of GPs (M), support generates positive expectations among participants (M), and GPs think QC meetings with their peers will be useful (M).

*…so obviously you get some CME credits you can use for accreditation (1)*

*…. some packets some information on a one topic or another in way so it can be used in QCs by the local people, often and this is working the best, is having someone who is coming with the information and carrying it into the QCs (1).*

*And I think that ... the other thing that is important to the group is the CME CPD points that they get and the funding from the government to attend meetings. That is all supporting the meetings as well (2).*

*...if you want to be recertified, every five years you have to document at least 20 hours in a QC (3).*

*'Need for relatedness'*

If a steady group of members engages in socially enjoyable contact, led by a skilled facilitator who, e.g., introduces people to each other, opens discussions, clarifies and summarizes statements (C), then group members will get to know each other and decide on rules that they are willing to follow and so build a safe environment based on trust (O) because members want to be among and to interact with equals (M).

*..this problem (no trust in the group because of competition about patient contacts) will be solved in a couple of years. When we started it was certainly that way but since about one in four is going to retire within the next five to ten years this will be solved and we get shortage of GPs and maybe that will make it easier for a peer review groups to have more trust and … and find each other to work together and to tackle new problems that may depend on shortage of GPs instead of too many (1).*

*…. it is the same, it is always the same 20 persons who are the member but once you will have 12 persons and the next time 6 will be the same but 6 will not have attended the last time and some come the next time again, so, the group is a fixed group, it is – of course, if you only have to participate twice a year, your group will not always be the same and it will vary a little bit, depending who is coming and who is not (1).*

*… but it became clear that we started to get to know each other and the sensibility of each other and to dare to tell about how we handle things and we learnt how to handle each other in a respectful way. Now we have to see how it continues (1).*

*I think ... the social aspect like you said you are right to discuss that because that is important. …. And I think that is an important part of the meetings (2).*

*And there is a rule in the group about honesty that if we discuss something that… that should stay in the group, it does not leave the group and that it stays in the group and I think that is respected because over the years there is much more honesty as the years go by (2).*

**Supplemental material 10 consolidated programme theory, including illustrative quotes and complete list of supporting quotes**

*We do have a meeting now I think in September or October where the doctors get together in a meeting in the afternoon and then we have a social gathering and for each of the group meetings we have coffee or tea and something to eat before the meeting this is important I think because a lot of doctors come for their surgery and they are tired they are fed up and they can have a cup of coffee and a bit of a (inaudible because she laughs) and they are going into the small group as a better doctor. the social aspect I think is very important…and we have half an hour with coffee and sandwich and then we start the meeting (2).*

*We have ... we would have done these rules in the very beginning when we started the groups, now we know each other for so long that there is no need to ... I think people are very respectful for each other and not necessarily to like each other because there people in the group who do not like each other and I think a norm like that would be difficult I think the rule is to be respectful and even if you don't like the person or agree with them that you are not disrespectful (2).*

*we usually start with what we call the round where everybody tells what case is on their minds buggering them or causing them problems and if some of them is very important ... we save some time for the end of the group (3).*

*I think the group make their own rules for conduct and in my group, we revise them quite often, so if we had some incident that was not so nice, we try to find better ways of behaviour towards each other and then the facilitator has quite a lot of authority, and if the facilitator is not able to exercise that, they can get help from four or five facilitator coordinators at the medical association. then they will come and help us in the group itself (3).*

*There should be like in many other countries … at least the impression I get from for instance Sweden, the Netherlands, Australia, New Zealand, there is a lot of government support - but in Norway we actually do not have much at all. So, there should be much more understanding from the mostly national bureaus but also form the local authorities how important this is. There has not been enough understanding in the medical corps to ...h to do it and usually comes on top of all the other work and it is usually unpaid. So, we have to do it at night and during weekends (3).*

*…and we do that at dinner time so we can have some food together, we have dinner and we can enjoy food at the same time (4).*

*… I think if you know people a little bit you feel more comfortable … to talk with if there are too many people who you never seen before and never talked to before, then it is difficult to open up and talk about (4).*

*this year ... we try to have kind of rules to be more organised in the group, so I think we try to keep it working, and … I think that is the challenge, but we should think and have deeper reflection about what the real impact on our practice is (4).*

'Need for autonomy and control'

If the group chooses its own topics and facilitator (C), then they will feel they own the QC (O) because this satisfies their need for autonomy, a feeling of being in control of one' s own behaviour (M).

*And they discuss cases, and a topic is picked for the month and an education module occurs around a particular area the doctors bring patients they are looking after and there is a discussion about the cases and the topic area and it is facilitated either by the leader of the group or the tutor, the CME tutor in a particular area (2).*

*…the group decides to change the programme based on new things that are happening or changes in medicine that are happening and (inaudible) there is a general structure plan for the year but then it changes if something changes … if some group says they would like to cover this or that particular area, there are changes during the year. So, the programme adapts to the needs of the group (2).*

**Supplemental material 10 consolidated programme theory, including illustrative quotes and complete list of supporting quotes**

*…exactly big autonomy, the groups decide there is no pressure from the political system and there is no pressure from anybody and that is why this system is so successful – the doctors can choose (2).*

*No there's is a group leader (facilitator), and they are free to elect him or her, and they have to fill in one sheet of paper, where they have to tell date and time and theme and list of attendees (3).*

'Size of the group affects communication'

If group size exceeds 15 (C), then interaction among group participants decreases (O) because participants cannot keep up with all of the other participants and follow their conversations (M).

*…the group would be 10 to 12 people at the most and they would have a group leader or a tutor in the group that is the facilitator in the group and these groups would meet regularly every month and they would know each other because they meet eight times a year. Knowing and trusting each other is really important when doctors talk about their patients (2).*

*For instance, if ... I think …15 people are too many …I think 8 is enough. and ... the stress increases if there are more…. the smaller the group is the better the trust and talking (4)*

'Variety of characters stimulates reflection – cognitive dissonance'

If members of the groups have individual character traits and describe differing professional experiences but accept each other's views (C), then they can learn from each other (O) because individual attitudes and behaviours will contrast with their peers' knowledge and cause cognitive dissonance that makes them reflect on their way of working (M).

*I think it would be logical and ... more (better) with more diversity and ... with more like an enrichment (4).*

*…. because you can learn from (other) people with more experience, … … you have a (another) way of thinking and a (another) way of talking about stuff, situations, that are different I think, so I think it is about different knowledge (4)*

'strong cognitive dissonance threatens self-image'

If individuals feel too strong a cognitive dissonance when integrating new knowledge (C), then they can disrupt group dynamics and the QC process halts (O) because this threatens their self-image and they feel at risk of losing their professional identity (M).

*Yes, we do yea, we have doctors who are ... difficult in the group, yes, and they are difficult because they have very firm views and they spend very little evidence on reality. Then it is very important that you have good group leaders and leadership … It is very few … you know trying to sabotage the group…. and they don't tend to change behaviour (2).*

*it is more about personal reasons ……. one (participant) is really expansive and always talking about her and compares everything with herself, and she pretends to know the way …. we can't really ... function and discuss as we wanted to, you know; it feels like competition … I don't know what happens … at that moment but …I don't think we have a good atmosphere then (4).*

### *Learning environment*

'Feeling safe and not vulnerable'

**Supplemental material 10 consolidated programme theory, including illustrative quotes and complete list of supporting quotes**

If participants trust each other (C), then they can describe how they work and admit what they don't know (O), because they feel safe rather than vulnerable (M).

*... if they start any discussion, that is one of the problems, most of the time they just invite some external speaker a specialist or someone with a special interest to come and present something and afterwards they will have questions to speaker and perhaps discuss a little bit in between depending on the speaker and perhaps the facilitator if he really wants to facilitate but that is most of the time what is happening (1).*

*… (one of the important things) ... is building the trust and building the trust…? I had a very nice compliment of one of my colleagues after wards which we have been working together ...she told me you know one of the things I learnt from you one of the things I experienced from you is that mm opening up with difficult cases and showing that you don't know everything is showing that you are vulnerable and not knowing what do with it …you build up trust because if you dare doing this gives us the confidence that we also can do that … (1).*

*...one of my experiences but that we had in the practice last month I think it was that we took up the discussion about cases with the trainee and we realised by discussing cases that ... you often find gaps in your knowledge (1).*

*…and if people know each other within the group they are very honest and very open and they just ….and they discuss worries and concerns and there is a lot of that if the group is functioning well and everybody is feeling comfortable and there is a good level of trust in the group and they can talk about their cases (2).*

*…when a doctor gets upset, that has happened over my years and usually there is a kind of…within the meeting and they upset I would usually ... deal with that situation during the meeting and if they are upset and they are quiet and then I will actually go to them after the meeting but I will never let a doctor go home with issues that somebody got upset because the last thing I think a doctor should go through in small groups is ending up feeling upset or demoralised (2).*

*there is surprisingly ... huge openness and some people take it up and tell 'I have made a mistake and I ... feel bad about it' (3).*

*…we talk about cases we have social bonding; we are a group who feels safe, it is like a safe climate, we talk about ... our difficult situations (4).*

*We know each other very well, so I don't think anybody gets angry for this…and nobody .... gets emotionally the wrong way… if you understand what I mean (5).*

'Need for competence and self-actualisation'

If the facilitator supports participants and encourages them to tell their stories and share their experiences in a safe environment, e.g., by encouraging interactive responses, through discussions and by summarizing statements, (C) then participants will be involved and share their positive experiences and failures (O), because they want to improve their competency, a sense of self-efficacy to achieve specific objectives (M), gain professional confidence (M) and achieve professional self-actualisation (M).

*…having to share feelings of sometimes being powerless in certain situations was one of the things that built up the group the group feeling and which made everybody feel relieved, maybe not relieved but feeling confident and this is going to work (1).*

*There is no kind of structure that is imposed on the group and that makes the group actively by into the learning process because a lot of doctors bring information into the group and they bring learning from other places into the group that they have obtained so it is a very, it is a mix of learning from various places (2).*

**Supplemental material 10 consolidated programme theory, including illustrative quotes and complete list of supporting quotes**

*... and people will actually discuss for instance difficult moments in cancer treatment or cancer care, people will bring stories about patients but they will often bring problems about members in the family and the difficulties of being a GP and having to cope with this, and the major problems about being a GP, and that is very powerful stuff because that is about the personal aspect of being a doctor (2)*

*that is an 'after' discussion – I often call this the hidden curriculum because I think that is very important, I think that ... a lot of doctors over the years have been in distress and it is important to talk about this (2).*

*the truth is that if you are a doctor and if you want to do a good job then you have to make quality improvement and patient safety a part of your profession ... broader knowledge (3).*

*And ... I think you learn a lot of basic things you need in order to be a g good doctor for your patients, you learn respect, you learn to hold yourself back to be able to let the other people speak and the other to take ... the front floor. Social control is quite important in many ways. ... and you learn that much easier in a group than on your own (3).*

*...someone tells which was a typical situation for one of them, for one of us – sorry… and sometimes we choose…. for example, ... we usually choose something that happened yesterday or the day before (4).*

*We prefer the clinical cases that we are difficult and where we have questions or bad emotions and ... … we prefer that kind of a (difficult) decision because we ……. because first, for the person who explains the situation; and this is a good way to be or to get rid of the pain and talk about that and for the others it is always interesting because most of the time one of us had already been or experienced ... or felt that pain or talked about a situation that is similar. …it is like ahh mutual understanding and we can understand and talk about it with each other, and it is ... a good feeling if you see other people had the same and we understand each other (4).*

*…but it is much more it is about personal feelings and points of views in life or fear ... or non-pleasant ... feelings or something with the people we have like difficult patients and our human relations with the patients, because no we don't have someone to summarise all the facts and all the feelings and ... there is no one who takes care of that what we do in the group (4).*

*I think it is important for psychological point of view not the feeling to be alone sharing your thoughts and experiences with friends and colleagues …. same GPs we have the same profession. I realise we are all in the same boat….it is also very stimulating to keep learning (4).*

*…and the fact that you can explain it to the others makes you realise that ... you have a bit anxiety about it and all the others tell you that this ok – not just because they want to comfort you …then you realise that you became nervous about something very quick …even if you did something good after all….the group at this moment is very ……… a peaceful place and a good way of being with yourself and your own way of practising and it increases your self-esteem as well (4).*

*But sometimes it is about our problems ... …... our professional life…about our patient about some case ……diagnostics or prescriptions (5).*

'Previous knowledge is activated'

If participants exchange case stories and experiences while actively listening to each other in the presence of a skilled facilitator in a safe environment (C), then they will share their knowledge by telling their own relevant stories (O) because the process activates knowledge they already possess (M).

*well .in different way …. it can be telling about a case even analysing a critical incident, telling about critical incidents is important to us, that can be a discussion of a guideline, a ... that can be a well something new out of the literature, ... these are the ways we want to do it (1).*

**Supplemental material 10 consolidated programme theory, including illustrative quotes and complete list of supporting quotes**

*it does satisfy us when we can discuss about our own work and about our own cases, and we feel closer in the group when we stimulate each other's thinking (4)*

*When we talk about cases, case discussions, that is the most efficient part of the hour, because every participant wants to ……… talk about and tell something about ... case ... about cases we experience. Sometimes, we have so different opinions and I f someone saw us from the outside, then …. they would think this is a crazy group…. but (laughs) but I think it is constructive…and we learn from each other because ... then, we talk about different aspects of the case (5).*

'Immediate relevance for the practice'

If QCs use the technique of experience-based learning (C), then knowledge becomes more relevant to GPs (O) because it relates to their everyday work and is therefore of immediate use (M).

*...a lot of the doctors will start with a clinical case, but then come to an overview and then discussions and the next step is organising the GP surgery for that -it is quick wins (3).*

*I think sometimes we … we have ... it is more like an administrative part about the administration the administrative things about how to do the replacement of GPs, like all the papers and all the declaration stuff that is necessary for this ….it helps…. I think it is very helpful to talk about that (4).*

*We think about …when one participant tells a story… talks about the case and after that we talk about what we think ...everyone in turn…we have only women participating in the group (laughs) …. we say what we think is correct and we talk about what each one of us would do in this situation, what we think she could do better…we look if we can find some evidence about that (medical facts) … and we can use it right after (5).*

'Cognitive dissonance'

If participants discuss and reflect on their work processes (e.g., based on trustworthy data or personal experiences) during a professionally facilitated exchange of positive experiences or failures (C), then they discover knowledge gaps and identify learning needs and relevant topics (O) because their own attitudes and behaviours may differ from their peers', creating cognitive dissonance that makes them reconsider their own way of working (M).

*…. the government ...m is now offering ... the possibility but it is quite informal it is not on a massive level they offer the possibility to discuss ... some indicators on polypharmacy to be discussed with a ... an expert of the government and ...... then they offer the results of the QC and individual results to the people who are participating there and then they start a discussion about polypharmacy and that is existing (1).*

*…well if you tell a story we are doing this in this way in our practice, ... another practice could tell, well in our practice we see thigs differently and we do it another way and or it could be that … we help think about the situation with a difficult patient how you … you can handle it in a different way then somebody else will tell you, well, what do you think about that maybe this could be a way or this …have you considered this with this patient and ... perhaps you could take up and discuss with the patient how he feels about that ……. it is often gives you the opportunity when you get stuck with difficult patients and mmm to get new energy and to have …mmm to listen to the way other people would handle it can help to open up and take new initiatives instead of blocking and having the feeling that you don't get any further with the patient (1).*

*…it is not just an easy push on one bottom but hard work ... on the other hand, we have a feedback from the government ... every two to three years which offers a lot of data about your prescription and about the population you treat and which you can use but that is always old data. We will now get one in the next months to come and that will contain data from 2015 it is now 2018!!! which will be analysed then so that is quite a problem (1).*

**Supplemental material 10 consolidated programme theory, including illustrative quotes and complete list of supporting quotes**

*Yes, Yes, prescription habits so ... ... prescription habits at the moment in Ireland is ... so if you are a public GP and you have a GMS number, then you get feedback on prescribing actually only in one area at the moment and that is benzodiazepines and you get that every year on benzodiazepines but you do not get feedback on anything else (2).*

*When for example a GP … in a gr ….group is saying that he does a particular thing that is purely not right not evidence based or in fact is wrong then the group are very good …I think because they know each other… they do not agree with the doctor but they actually discuss it in the group and a few other doctors say what they would do which is usually different and they usually say ' you may consider this as a different way of doing it because if you do it your way , this is what I find happens…' and there is never an issue where somebody needs to feel bad but they know that whatever they currently are doing is not what the other would (2).*

*I think because when you have had doctors I the group for a long, long time and working in practice for a long time I think you have to consider what people currently are doing and what they accept as appropriate for their practices or for their work. and I think to introduce new guidelines and new evidence you have to look at what people are currently doing and to get people to accept a change and see why this change would be necessary as well and sometimes the change is not necessary for the group; if you don't know whatever they are currently doing ,and if they are not exchanging ideas within the group then ..that really…they are not learning then because ... I think my criticism of guidelines and evidence is that they are not always practical to implement (2)*

*We use some data extraction software from the electronic health record so that every doctor gets his own indicators ... in a report that tailors the theme (3).*

*We usually do it (comparing each other's data) as a plenary thing and I can always say as a facilitator what about indicator 13 and then we go around the table what figures do you have and how would you explain them and the huge differences between the results. so, there is a …. a special part of data report of the indicator we go through in each meeting and we …when we have done that, they usually don't have use of the facilitator because the discussion is quite intense (3).*

*Firstly, I think they learn a lot about quality indicators and then you have to go into the matter why they differ so much. Why yours is so different from mine, and then you have to look at age spread of the population, my work, if I work a lot 'on call' for instance, which is different from sitting in the office all the time (3).*

*…yes, sometimes we choose difficult situations and sometimes we don't choose and we talk about the last situation we had the day before and sometimes like a simple disease that is not so difficult, so we talk about, because even if it seems to be easy we have different ways to do this and it is interesting to talk about even easy situations, because all the other do it in a different way (4).*

*We do have practice mirrors about hypertension, about diabetes and ... now we have some I work on some audit about prescription of warfarin, which gives a lot of interesting discussions (5).*

*… and I see only me ... is this ok or did do something wrong? but now we compared and compare two different practices in two different parts of Croatia and we have similar results, which surprised me (5).*

*We do that just like in case discussions; some of us have a little …presentation …we talk about guidelines or evidence-based information ….and we ahh we that …. colleagues talk about what they do in their practices and .... what she can do and why, …. giving the reasons…and after that we talk about …every participant talks about what she does in practice and what they don't do and the reason why they don' do it (5).*

'Social learning'

If the facilitator uses purposeful didactic techniques (e.g., brainstorming, contentious or consensus discussions, or role play) to keep the group active and to reward exploratory behaviour during reflection on the work process (C), then the group will create a learning environment that promotes knowledge exchange (O) because learning is a cognitive process in which participants observe and imitate their peers' behaviour to gain social approval (M).

*So I think that the more experienced GPs bring in their cases into the groups and they discuss their experiences within the groups and I think this is very powerful for the group and the younger GPs bring in …they have the latest evidence in their head and the guidelines and they bring it in ..and the mix of managing the patient with the evidence and the guidelines and the practical bit from the older GP who has the experience I think this is really the powerful bit in the group and … and this is where the learning really occurs (2).*

*…and I (Facilitator and tutor) don't have the arrogance to believe that they leave that meeting and go and change their practice but they are certainly aware of that their practice is not what the …. the rest of the group's is (2).*

*…case discussions are important so cases are a huge part of the group and the other thing we would sometimes do is ... a role play we also have used video consultation playing video cases or other reals life scenarios and the other thing we should use is discussion groups. So, you know like working groups for example I have twelve people and if I have something new, I might split the group into groups of four and so people would work within these smaller groups and then they carry their points of views back to the whole group. And it is not an individual but the whole small group who feeds back, it is the group it is a safer place (2).*

*yes, ... in the beginning we thought this (sharing data) had to be in pairs or triplets because we thought that people were not willing to share, but that was quite wrong. they love to share (3)!*

*…but we like to learn and understand how the others do; so, it is a learning from each other, yes that is what it is [in French] (4).*

### *Adapting, creating and testing new knowledge*

'Interdependence between health insurance companies/physician network organisations and GPs'

If physician network organisations require continuous QC activities (C), then QCs will negotiate priorities and design creative solutions(O) because the tension between autonomy and obligation spurs the group to act and negotiate together to reach a common goal (M).

*It may be important for the emerging of QCs, that it becomes a mandatory thing (QI) and after all, we have the same goals (as the health insurance companies) (4).*

*We have as an obligation in contracts with our insurance to have …… … peer groups…… then ... I don't know how many times we should meet, actually. But we don't have or get much money out of this (5).*

*'Threat to professional autonomy'*

If GPs feel that the QC programme is only a top-down managerial intervention to reduce costs (C), then they will not be motivated and will not participate (O) because they feel unsafe and think they lack autonomy in their clinical role (M).

*…no there are no demands, that wouldn't help, we have to do and there can be wishes how, but we decide….it wouldn't work otherwise (3).*

'Interdependence among group members'

If participants maintain a trusting learning environment that promotes knowledge exchange, assisted by facilitators who use professional techniques (e.g., contentious discussion, reaching consensus, and role play), (C), then participants will adapt and generate new knowledge for local use (O) because they see themselves as being similar, and so act and negotiate cooperatively to achieve a common goal (M).

*I think that a group …cannot just be presented with things like here is the evidence, take it or leave it and goodbye and I don't think that works, I think that people need to ... participate and in the learning and they have to show what they are currently doing whether it is the correct thing or not; it needs to be discussed and adjusted and shared within the group (2).*

'Identifying and removing barriers to change'

If participants, supported by skilled facilitators, address barriers to change (C), then they are more likely to implement the innovation (O), because participants help each other develop strategies to identify and overcome these barriers (M).

*And I think you have to have guidelines that are workable for doctors who are you know seeing 30 to 40 people every day and if they want to implement change for the better they have to be feasible and practical and I think the only way to do that is to consider what they are currently doing. And what the barriers are to new care (2).*

'Need for competence, autonomy and relatedness'

If participants create new knowledge and plan an implementation strategy (C), then they feel satisfaction, responsibility and stewardship (O), because this fulfils their need for competence (being able to achieve specific objectives) (M), autonomy (a feeling of being in control of their own behaviour) (M), and relatedness (a sense of connection to a larger group) (M).

*No data but confirming comments.*

'Intention to change'

If participants publicly announce their intention to change (C), then they are more likely to implement the change (O) because they and others in the group both think it is a good idea and believe they can carry it through (M).

*…we ask the group to give a feedback on how they feel that would change them or their practice and the routine of care for their patients. So, they usually the group …we end the meetings with a feedback a summary and a feedback and a feedback from the group what it is they feel they want to change ... and I think that is the opportunity to …. not everybody participates in that…. but ... most people do ….and they'd say look this is what I learned this is new for me this is what I am ... going to change in my practice (2).*

*We talk about …how we shall we implement the guidelines and shall we implement this in our everyday process, what steps we can implement and how and what we cannot implement and why not....... ... how do we need support from our hospital-based colleagues…? ... in some steps of the implementation of the guidelines…. and ... sometimes we need help of our medical association because in ... some steps when we talk about guidelines, we don't have the things (equipment) in our practice (5).*

'Testing new knowledge'

If participants validate and test new knowledge in a QC, moderated by a skilled facilitator, in a safe environment (C), then they feel confident putting that knowledge to use in everyday practice (O) because they have had the opportunity to practise and familiarise themselves with the innovation (M).

**Supplemental material 10 consolidated programme theory, including illustrative quotes and complete list of supporting quotes**

*..time to reflect on this practice is actually something that is very important and you have to figure out where the guidelines fit in and you reflect on what you are doing and the group and the process finds out the is correct the use of the guidelines and then for you as a practitioner you can look at that and see what practical that has changed over time and I think that the idea of a quality circle meeting is trying make changes dramatically is not practical I think doctors need to look at ideas and look at the practical parts to see what they can do and change slowly over time (2).*

### *Repeating the process*

'Gaining confidence in an innovation'

If the group repeatedly practices implementing and coping with an innovation (C), then they trust their own competence and turn the innovation into a habit (O) because successful outcomes increase confidence in their abilities (M).

*... then we meet again after four months and usually the …their quality improvement project ... didn't really happen or just a little bit, and we discuss the reasons for that and how we could amend that etc. etc. (3).*

*…and ... yes and then we present it and I also present it wherever we work and at the practices where we work (4).*

'Repetition priming and automaticity'

If participants build a steady group and practice using QI tools (C), then they will successfully implement new knowledge into everyday practice (O) because successful responses increase with repetition: 'practice makes perfect' (M).

*...but it is really a double thing. it is about a theme but it is also about quality improvement. And the aim and goal are that they find it so rewarding that they use this this technique again and again. ……in their own surgeries and in their own groups (3).*
