## Supplemental material 11 for "How and why do Quality Circles work for General Practitioners - a realist approach"

**Supplemental material 11 Summary of the QC process and its implications**

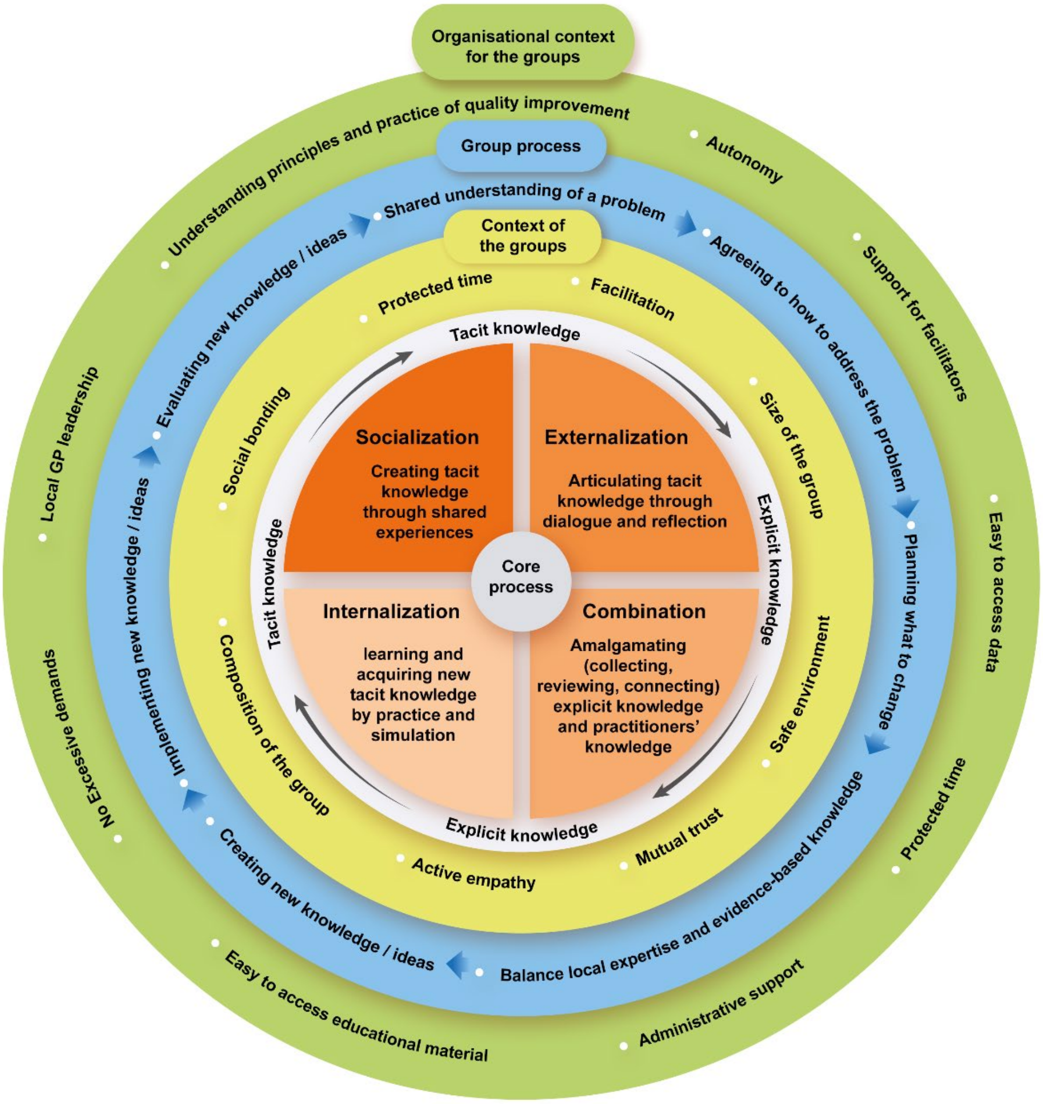


Legend:

The rings represent the levels of context and their associated processes. The core process is in the centre, illustrating the exchange of knowledge and the creation of innovations in QCs. The process is a spiral rather than a circle, because participants add experience and new knowledge at each turn of the cycle. The size and composition of the group, the social bonds between participants and their mutually benevolent attitude all foster mutual trust and create a safe environment in which participants can have frank discussions. Protected time and skilful facilitation lay the groundwork for a successful core process. At the next level, participants begin with a shared understanding of an issue and agree how to address it and what needs to be changed, ensuring the success of the group process. When QCs solve problems and innovate, they should balance local expertise (soft knowledge) with evidence-based information (hard knowledge); then they can generate new ideas to be tested and implemented in everyday practice. The QC process requires considerable professional and administrative support at the organisational level, so professional associations or university departments must teach QC members the principles and practices of QI and their use, and train and support facilitators. Organisations should also provide easy access to performance data and evidence-based material. Administrative organisations, whether health insurance companies or governmental organisations, should allow QCs to have professional and administrative autonomy and let them take the lead in QI, without placing excessive demands on the group or its members. The level of legislation required to entrust GPs with QI will vary depending on a country's health-care system, and could be enacted at national or local government level.
