## Supplementary material for "How and why do Quality Circles work for General Practitioners - a realist approach": Qualt Standards RAMESES

### QUALITY STANDARDS FOR REALIST SYNTHESIS (for researchers and peer-reviewers)

#### 1. The research problem

Realist synthesis is a theory-driven method that is firmly rooted in a realist philosophy of science and places particular emphasis on understanding causation and how causal mechanisms are shaped and constrained by social context. This makes it particularly suitable for reviews of certain topics and questions – for example, complex social programmes that involve human decisions and actions. A realist research question contains some or all of the elements of 'What works, how, why, for whom, to what extent and in what circumstances, in what respect and over what duration?' and applies realist logic to address the question. Above all realist research seeks to answer the 'why?' question. Realist synthesis always has explanatory ambitions. It assumes that programme effectiveness will always be partial and conditional and seeks to improve understanding of the key contributions and caveats.

| Criterion | Inadequate | Adequate | Good | Excellent |
|---|---|---|---|---|
| The research topic is appropriate for a realist approach | The research topic is:<br>• not appropriate for secondary research; and/or<br>• does not require understanding of how and why outcomes are generated. | The research topic is appropriate for secondary research. It requires understanding of how and why outcomes are generated and why they vary across contexts. | Adequate plus: Framing of the research topic reflects a thorough understanding of a realist philosophy of science (generative causation in contexts; mechanisms operating at other levels of reality than the outcomes they generate). | Good plus: There is a coherent argument as to why a realist approach is more appropriate for the topic than other approaches, including other theory based approaches. |
| The research question is constructed in such a way as to be suitable for a realist synthesis | The research question is not structured to reflect the elements of realist explanation. For example, it:<br>• only requires description; and/or<br>• only requires a numerical aggregation of outcomes; and/or<br>• only requires summary of processes; and/or<br>• specifies methods that are inadequate to generate realist understanding (e.g. 'a thematic analysis of …') | The research question includes a focus on how and why the intervention, or programme (or similar classes of interventions or programmes - where relevant) generates its outcomes, and contains at least some of the additional elements, "for whom, in what contexts, in what respects, to what extent and over what durations". | Adequate plus: The rationale for excluding any elements of 'the realist question' from the research question is explicit. The question has a narrow enough focus to be managed within a realist review. | Good plus: The research question is a model of clarity and as simple as possible. |

| 2. Understanding and applying the underpinning principles of realist reviews | | | | |
|---|---|---|---|---|
| Realist syntheses apply realist philosophy and a realist logic of enquiry. This influences everything from the type of research question to a review's processes (e.g. the construction of a realist programme theory, search, data extraction, analysis and synthesis to recommendations).<br>The key analytic process in realist review involves iterative testing and refinement of theoretically based explanations using empirical findings in data sources. The pertinence and effectiveness of each constituent idea is then tested using relevant evidence (qualitative, quantitative, comparative, administrative, and so on) from the primary literature on that class of programmes. In this testing, the ideas within a programme theory are re-cast and conceptualised in realist terms. Reviewers may draw on any appropriate analytic techniques to undertake this testing. | | | | |
| **Criterion** | **Inadequate** | **Adequate** | **Good** | **Excellent** |
| The review demonstrates understanding and application of realist philosophy and realist logic which underpins a realist analysis. | Significant misunderstandings of realist philosophy and/or logic of analysis are evident. Common examples include:<br>• programme/intervention activities or strategies are confused with mechanisms<br>• no attempts are made to uncover mechanisms<br>• outcomes are assumed to be caused by the programme/intervention<br>• relationship(s) between an outcome, its causal mechanism(s) and context(s) are not explained<br>• some theory is provided but this is not explicitly linked to outcome(s) | Some misunderstandings of realist philosophy and/or logic of analysis exist, but the overall approach is consistent enough that a recognisably realist analysis results from the process. | The review's assumptions and analytic approach are consistent with a realist philosophy at all stages of the review.<br><br>Where necessary a realist programme theory is developed and tested. | Good plus: Review methods, strategies or innovations used to address problems or difficulties within the review are consistent with a realist philosophy of science. |

| 3. Focussing the review | | | | |
|---|---|---|---|---|
| Because a realist review may generate a large number of avenues that might be explored and explained, and because resources and timescale are invariably finite, it may be necessary to 'contain' a review by progressively focusing both its breadth (how wide an area?) and depth (how much detail?). This important process needs to be considered from the start and may involve iterative rounds of discussion and negotiation with (for example) content experts, funders and/or users. It is typical and legitimate for the review's objectives, question and/or the breadth and depth of the review to evolve as the review progresses. | | | | |
| **Criterion** | **Inadequate** | **Adequate** | **Good** | **Excellent** |
| The review question is sufficiently and appropriately focussed. | The review question is too broad to be answerable within the time and resources allocated.<br><br>There is no evidence that progressive focussing occurred as the review was undertaken. | Attempts are made by the review team to progressively focus the review topic in a way that takes account of the priorities of the review and the realities of time and resource constraints.<br><br>Attempts are documented so that they can be described in publications as appropriate. | Adequate plus: The focussing process is iterative. Commissioners of the review are involved in decision-making about focussing.<br><br>Decisions made about which avenues are pursued and which are left open for further inquiry are recorded and made available to users of the review. | Good plus: The review team draws on external stakeholder expertise to drive the focussing process in order to achieve maximal end-user relevance. |

| 4. Constructing and refining a realist programme theory | | | | |
|---|---|---|---|---|
| Early in the review, the main ideas that went into the making of a class of interventions (the programme theory – which may or may not be realist in nature) are elicited. This initial programme theory sets out how and why a class of intervention is thought to 'work' to generate the outcome(s) of interest. This initial programme theory then needs to be 're-cast' in realist terms (a rough outline of the contexts in which, populations for which, and main mechanisms by which, particular outcomes are expected to be achieved.) This initial tentative theory will be progressively refined over the course of the review. | | | | |
| **Criterion** | **Inadequate** | **Adequate** | **Good** | **Excellent** |
| An initial realist programme theory is identified and developed. | A realist programme theory is not offered<br>or;<br>A program theory is offered but is not converted to a realist program theory at any stage of the review. | An initial program theory is identified and described in realist terms (that is, in terms of the relationship between contexts, mechanisms and outcomes).<br><br>The refined theory is consistent with the evidence provided. | Adequate plus: An initial realist programme theory is set out at the start. The theory is refined iteratively as the review progresses. | Good plus: The relationship between the programme theory and relevant substantive theory is identified.<br><br>Implications of the final theory for practice, and for refinements to substantive theory where appropriate, are described.<br><br>The final realist program theory comprises multiple context-mechanism-outcome configurations (describing the ways different mechanisms fire in different contexts to generate different outcomes) and an explanation of the pattern of CMOs. |

#### 5. Developing a search strategy

Searching in a realist review is guided by the objectives and focus of the review, and revised iteratively in the light of emerging data. Searching is directed at finding data that can be used to test theory, and may lie in a broad range of sources that may cross traditional disciplinary, programme and sector boundaries. The search phase is thus likely to involve searching for different sorts of data, or studies from different domains, with which to test different aspects of any provisional theory.

| Criterion | Inadequate | Adequate | Good | Excellent |
|---|---|---|---|---|
| The search process is such that it would identify data to enable the review team to develop, refine and test programme theory or theories. | The search is incapable of supporting a rigorous realist review. Common errors include:<br>• The search is driven by a methodological hierarchy of evidence (e.g. privileging RCTs) rather than the need to identify data to develop, refine or test program theory/ies<br>• The search process is not informed by the objectives and focus of the review<br>• The database(s) selected are narrow in the subject matter that they contain (e.g. limited to specific topics rather than extending to social science, psychology etc.)<br>• Searching is undertaken once only at the outset of the review and there is no iterative component | Searches are driven by the objectives and focus of the review.<br><br>The search strategy is piloted and refined to check that it is fit for purpose.<br><br>Documents are sought from a wide range of sources which are likely to contain relevant data for theory development, refinement and testing.<br><br>There is no restriction on the study or documentation type that is searched for. | Adequate plus: further searches are undertaken in light of greater understanding of the topic area. These searches are designed to find additional data that would enable further theory development, refinement or testing. | Good plus: the searching deliberately seeks out data from situations outside the program under study where it can be reasonably inferred that the same mechanisms(s) might be in operation. |

<table>
<tr><td colspan="5">6. Selection and appraisal of documents</td></tr>
<tr><td colspan="5">Realist review requires a series of judgements about the relevance and robustness of particular data for the purposes of answering specific questions within the overall review question.<br>An appraisal of the contribution of any section of data (within a document) should be made on two criteria:<ul><li>Relevance – whether it can contribute to theory building and/or testing; and</li><li>Rigour – whether the method used to generate that particular piece of data is credible and trustworthy.</li></ul>The selection and appraisal stage may need to run in parallel with the analysis stage.</td></tr>
<tr><th>Criterion</th><th>Inadequate</th><th>Adequate</th><th>Good</th><th>Excellent</th></tr>
<tr><td>The selection and appraisal process ensures that sources relevant to the review containing material of sufficient rigour to be included are identified. In particular, the sources identified allow the reviewers to make sense of the topic area; to develop, refine and test theories; and to support inferences about mechanisms.</td><td>The selection and appraisal process does not support a rigorous and complete realist review. For example:<ul><li>Selection is overly driven by methodological hierarchies (e.g. the restriction of the sources to RCTs to the exclusion of other forms of evidence)</li><li>Sources are appraised using a technical checklist for a particular method (e.g. assessment of quality for an RCT) rather than by making a defensible judgement on the relevance and rigour of the source</li><li>Selection and appraisal processes are overly restrictive and exclude materials that may be useful for a realist analysis</li><li>Selection and appraisal processes are not sensitive enough to exclude irrelevant materials</li></ul></td><td>Selection of a document for inclusion into the review is based on what it can contribute to the process of theory development, refinement and/or testing (i.e. relevance).<br><br>Appraisals of rigour judge the plausibility and coherence of the method used to generate data.</td><td>Adequate plus: During the appraisal process limitations of the method used to generate data are identified and taken into consideration during analysis and synthesis.</td><td>Good plus: Selection and appraisal demonstrate sophisticated judgements of relevance and rigour within the domain.</td></tr>
</table>

| 7. Data extraction | | | | |
|---|---|---|---|---|
| In a review, data extraction assists analysis and synthesis. Of particular interest to the realist reviewer are data that support the use of realist logic to answer the review's question(s) – e.g. data on context, mechanisms, and outcome configurations, demi-regularities, middle-range and/or programme theories. | | | | |
| **Criterion** | **Inadequate** | **Adequate** | **Good** | **Excellent** |
| The data extraction process captures the necessary data to enable a realist review. | The data extraction process does not capture the necessary data to enable a realist review. For example:<br>• Data extraction is undertaken mechanically and with no attention to how the data informs the review<br>• No or very limited piloting has been undertaken to test aspects of the data extraction process and improve it | Data extraction focuses on identification and elucidation of context-mechanism outcome configurations and refinement of program theory.<br>Piloting and refinement of the data extraction process has been undertaken where appropriate.<br>Quality control processes are in place to check that all review team members apply common processes and standards in data extraction. | Adequate plus: Data extraction processes support later processes of analysis (e.g. by organising data into sets relevant for later analysis). The data extracted is comprehensive enough to identify main CMO patterns. | Good plus: The data extraction process is continually refined as the review progresses, so as to capture relevant data as the review question is focussed and/or program theory is refined. |

| 8. Reporting | | | | |
|---|---|---|---|---|
| Realist reviews may be reported in multiple formats – lengthy reports, summary reports, articles, websites and so on. Reports should be consistent with the publication standards for realist synthesis. (See RAMESES publication standards: Realist syntheses at: http://onlinelibrary.wiley.com/doi/10.1111/jan.12095/full or http://www.biomedcentral.com/1741-7015/11/21). | | | | |
| **Criterion** | **Inadequate** | **Adequate** | **Good** | **Excellent** |
| The realist synthesis is reported using the items listed in the RAMESES Reporting standard for realist syntheses. | Key items are missing. For example<br>• No defined research question<br>• Limited or no reporting of the review's processes (i.e. methods used)<br>• Limited or no explanations and justifications provided for any adaptations made on the realist review process<br>• Insufficient detail is reported to enable readers to judge the plausibility and coherence of the findings | Most items reported. In particular the following items should be reported:<br>• Rationale for review<br>• Objectives and focus of review<br>• All method section items (i.e. items 5 to 11 in the RAMESES publication standards: Realist syntheses) | All items are reported clearly and in sufficient detail for an external reader to understand and to judge the methods used and the plausibility and coherence of the findings. | Good plus: The report is well written and easy to understand. Additional materials are made available for external readers to investigate aspects of the review in more detail. |

For details on how these quality standards were developed, please see:

Wong G, Greenhalgh T, Westhorp G, Pawson R..Development of methodological guidance, publication standards and training materials for realist and meta-narrative reviews: the RAMESES (Realist And Meta-narrative Evidence Syntheses - Evolving Standards) project. Health Serv Deliv Res 2014;2(30)

RCTs = randomised controlled trials
