## Supplementary material for "How and why do Quality Circles work for General Practitioners - a realist approach": Ethics_1

**MS IDREC**
Research Services, University of Oxford

University Offices, Wellington Square, Oxford, OX1 2JD
 http://www.medsci.ox.ac.uk

**CONFIDENTIAL**

Ref: MSD-IDREC-C1-2015-002

Dr Adrian Rohrbasser
Grubenstrass 52
9500 Wil
Switzerland

9th January 2015

Dear Dr Rohrbasser

**CUREC checklist**

I am writing to acknowledge receipt of your CUREC-1 form for your project: **Quality Circles - Audit**

On the basis of the information you have provided this has now been approved by the Medical Sciences IDREC **subject to:**

**a) it is your responsibility to comply with the requirements for administering any tests or questionnaires and if in doubt to contact the publisher of those tests or questionnaires.**

The reference number for this project is **MSD-IDREC-C1-2015-002** and is valid for a period of **48 months** from the CUREC 1 approval date, **7th January 2015**. Please may I remind you that your project may be reviewed at some stage during an annual audit of projects.

**Amendments**

Should you at some stage alter some of the techniques or procedures then you should first undertake a checklist (CUREC-1) to see whether these changes alter the ethics of the research. If these remain the same then the committee will require notification of the changes to lodge with the project. If they do not remain the same then you may need to complete a CUREC-2 form and undergo further scrutiny by the committee.

Please do not hesitate to contact me if you have any queries about this.

Yours sincerely

Gill Halstead

Gill Halstead
Research Ethics Co-ordinator, Medical Sciences
