## Supplementary material for "How and why do Quality Circles work for General Practitioners - a realist approach": Ethics_2

**MEDICAL SCIENCES**
**INTER-DIVISIONAL RESEARCH ETHICS COMMITTEE**

Research Services, University of Oxford, Wellington Square, Oxford, OX1 2JD


**CONFIDENTIAL**

30th March 2016

Dr Adrian Rohrbasser
Grubenstrass 52
9500 Wil
Switzerland

Dear Dr Rohrbasser

**Amendment of Research Ethics Application**

**Ref No: MSD-IDREC-C1-2015-002 (R44921/RE001)**

**Project Title: Quality Circles - Audit**

Thank you for submitting a request for an amendment to the above project, which has been reviewed on behalf of the Medical Sciences Inter-Divisional Research Ethics Committee (IDREC).

I am pleased to inform you that your requests to:

- use Google Hangout on Air, to interview European experts, to test the findings by case studies they provide when talking about their projects in their countries

- add Dr Geoff Wong in the role of Supervisor (replacing Dr Sharon Mickan)

have been judged as meeting appropriate ethical standards, on the basis of the information provided to the IDREC.

Please do not hesitate to contact me if you have any queries about this.

Yours Sincerely

H. Barnby-Porritt

Dr. Helen Barnby-Porritt
Research Ethics Manager, Medical Sciences
