## Supplementary material for "How and why do Quality Circles work for General Practitioners - a realist approach": Consens sheet

**Consent Form for**

### *Exploring why Quality Circles work in Primary Health Care: A realist approach about how, why and under what circumstances they work - 18/12/2014*

---

***We appreciate your interest in participating in this research! Quickly read through these terms before agreeing to participate.***

| ***Please tick the appropriate boxes*** | **Yes** | **No** |
|---|---|---|
| **Taking Part** | | |
| I have read and understood the research information sheet dated 17/12/2014. | ☐ | ☐ |
| I have been given the opportunity to ask questions about the project. | ☐ | ☐ |
| I agree to take part in the project. Taking part in the project will include being interviewed and recorded audio. | ☐ | ☐ |
| I understand that my taking part is voluntary; I can withdraw from the study at any time and I do not have to give any reasons for why I no longer want to take part. | ☐ | ☐ |
| **Use of the information I provide for this project only** | | |
| I understand my personal details such as phone number and address will not be revealed to people outside the project. | ☐ | ☐ |
| I understand that my words may be quoted in publications, reports, web pages, and other research outputs. | ☐ | ☐ |
| *Please choose **one** of the following two options:* | | |
| I would like my real name used in the above | ☐ | |
| I would **not** like my real name to be used in the above. | ☐ | |
| **Use of the information I provide beyond this project** | | |
| I agree for the data I provide to be archived at the UK Data Archive.[1] | ☐ | ☐ |
| I understand that other genuine researchers will have access to this data only if they agree to preserve the confidentiality of the information as requested in this form. | ☐ | ☐ |
| I understand that other genuine researchers may use my words in publications, reports, web pages, and other research outputs, only if they agree to preserve the confidentiality of the information as requested in this form. | ☐ | ☐ |
| **So we can use the information you provide legally** | | |
| I agree to assign the copyright I hold in any materials related to this project to Adrian Rohrbasser. | ☐ | ☐ |

**Consent Form for**

#### *Exploring why Quality Circles work in Primary Health Care: A realist approach about how, why and under what circumstances they work - 18/12/2014*

---

1st Interview

| | | | |
|---|---|---|---|
| ____________________________ | | ________________________ | _________ |
| Name of participant | [printed] | Signature | Date |
| _________________________ | | ___________________ | ________ |
| Researcher | [printed] | Signature | Date |

2nd interview

| | | | |
|---|---|---|---|
| ____________________________ | | ________________________ | _________ |
| Name of participant | [printed] | Signature | Date |
| _________________________ | | ___________________ | ________ |
| Researcher | [printed] | Signature | Date |

Project contact details for further information:

Adrian Rohrbasser
Family Physician
MSc in Evidence Based Health Care
DPhil student at the Department of Primary Care Health Sciences in Oxford
New Radcliffe House, 2nd floor, Walton Street, Jericho, OX2 6NW, Tel: 01865 289300 (N.B. New Radcliffe House postal address: Radcliffe Observatory Quarter, Woodstock Road, OX2 6GG).


Work: Santémed Gesundheitszentrum Wil, Friedtalweg 18, 9500 Switzerland, 0041719135400,


#### Notes:

1. At the end of this project the audio and digital data collected from interviews with participants will be deposited at the UK Data Service for use by future researchers

This work is licenced under the Creative Commons Attribution-Non-Commercial-Share Alike 2.0 UK: England & Wales License. To view a copy of this licence, visit http://creativecommons.org/licenses/by-nc-sa/2.0/uk/
