## Supplementary material for "How and why do Quality Circles work for General Practitioners - a realist approach": Information sheet

### Participant Information Sheet - 18/12/2014

Dr Adrian Rohrbasser, Department of Primary Care Health Sciences in Oxford, New Radcliffe House, 2nd floor, Walton Street, Jericho, OX2 6NW, Tel: 01865 289300 (N.B. New Radcliffe House postal address: Radcliffe Observatory Quarter, Woodstock Road, OX2 6GG),

#### Study Title and date

***Exploring why Quality Circles work in Primary Health Care: A realist approach about how, why and under what circumstances they work 18/12/2014***

#### Background and aims of the study

Quality Circles are small groups of 6 to 12 professionals from the same background who meet at regular intervals to consider their standard practice. Quality Circles are commonly used in primary health care in Europe to improve standard of health care over time. They represent a complex social intervention that occurs within a fast-changing society. Numerous controlled trials, reviews and studies have shown small but unpredictable positive effects on behaviour change. Although QCs seem to be effective, it is difficult to understand how the results are achieved and how to generalise them with confidence. This research will examine how configurations of components and contextual features within QCs result in changes in health professionals' behaviour. The objective is to increase the understanding of the active components that lead to improved clinical practice among participants.

#### Who is organising and funding this research?

The research for this study is being undertaken by Adrian Rohrbasser who is a DPhil (doctoral) student in the Department of Primary Care Health Sciences at the University of Oxford. This project is being supervised by Dr Sharon Mickan, University of Oxford, Department of Primary Health Care Sciences and Dr Janet Harris, University of Sheffield, School of Health and Related Research (ScHARR). Both are experienced scientists in knowledge translation and in health care. This research is done without any funding and has been reviewed and approved by the University of Oxford Central University Research Ethics Committee (CUREC).

#### Why have I been asked to participate?

You as a stakeholder are important in the evaluation of Quality Circles and are invited to take actively part in this research. You are a general practitioner or administrator and either participant in QCs, facilitator, tutor or representative of professional bodies, networks providing primary health care or representative of health insurance companies. You receive this leaflet as additional information after the telephone conversation with the researcher.

#### What will taking part in the study involve?

The researcher will contact you once more to clarify questions and give you an opportunity to discuss participation and its consequences. If you agree to participation, he will make an appointment with you. At the meeting, you will be asked to sign a consent form. You shall meet the researcher for an informal interview and discussion in a short while and after a period of about one year. The meeting will take about an hour and a half.

### Participant Information Sheet - 18/12/2014

Dr Adrian Rohrbasser, Department of Primary Care Health Sciences in Oxford, New Radcliffe House, 2nd floor, Walton Street, Jericho, OX2 6NW, Tel: 01865 289300 (N.B. New Radcliffe House postal address: Radcliffe Observatory Quarter, Woodstock Road, OX2 6GG),


The researcher records the interview and may also take some notes. Before the researcher starts the interview and discussion, please, feel free to ask questions that may have arisen in the meantime. You may withdraw from the study without consequences at any time by advising the researcher of this decision.

#### What will happen in the study?

Firstly, you are invited to help formulate the research questions and inform the researcher about possible underlying and competing theories that make Quality Circles work. It will take about an hour to talk about who is involved and what actually happens in Quality Circles, when and where they take place, why they are organised and whether and how you think they work.

Secondly, in a later stage, you are invited to participate in the analysis of the results. The researcher will present you literature findings on Quality Circles and ask you whether they are in agreement with your experience or not. He will ask you about your opinion on different propositional statements found in literature that explain how, why and for whom they work and under what circumstances. Both steps are significant and add relevance and credibility to this type of research.

#### What are the possible benefits of taking part?

You contribute with your knowledge and experience to the interpretation of the literature findings. Iterative consultations with you will keep the research focussed on the relevant questions. You can also participate in the interpretation of the results. You may disseminate the findings among your colleagues and make use of the findings to improve practice.

#### What are the possible disadvantages and risks of taking part?

We hope that the experience of taking part in this study will be enjoyable for you. However, there is the possibility with work that looks at professional experiences that some upsetting memories may be recalled or talked about. You will at no point be obliged to talk about any topic that makes you uncomfortable.

#### How will my interview be used?

The interviews collected in this study will be used alongside other data sources to uncover more about Quality Circles. Publications which come out of this project may use quotations from your interview. Once the interviews have been deposited at the end of the project, future researchers may also wish to use and quote your interview in their work. On the consent form you will be asked to confirm that you are happy to assign your copyright for the interview to the researcher, meaning that you consent for researchers to use and quote from your interview. In all works produced using your interview, your name and other personal information will be anonymised.

#### What happens to the interviews collected during the study?

Interviews will be transcribed and stored digitally, managed by the researcher Adrian Rohrbasser for the duration of the project. Only the researcher will have access to the interviews and personal information.

### Participant Information Sheet - 18/12/2014

Dr Adrian Rohrbasser, Department of Primary Care Health Sciences in Oxford, New Radcliffe House, 2nd floor, Walton Street, Jericho, OX2 6NW, Tel: 01865 289300 (N.B. New Radcliffe House postal address: Radcliffe Observatory Quarter, Woodstock Road, OX2 6GG),

#### What happens at the end of the project?

If you agree to participate in this project, the research will be written up as a thesis. All participants are able to request a summary of the research findings should they wish to by contacting the researcher Adrian Rohrbasser. On successful submission of the thesis, it will be deposited both in print and online in the University of Oxford, to facilitate its use in future research. The digital online copy of the thesis will be deposited with the Oxford University Research Archive (ORA) and will be published with open access, meaning that it will be available to all internet users. At the end of this project the audio and digital data collected from interviews with participants will be deposited at the UK Data Service for use by future researchers

#### What should I do if I have any concerns or complaints?

If you have a concern about any aspect of this project, please speak to the relevant researcher (Dr Adrian Rohrbasser, telephone number 0041796036531) or his supervisor, Dr Sharon Mickan telephone number 0061 (0) 423 662944 who will do his best to answer your query. The researcher should acknowledge your concern within 10 working days and give you an indication of how he intends to deal with it. If you remain unhappy or wish to make a formal complaint, please contact the chair of the Research Ethics Committee at the University of Oxford (using the contact details below) who will seek to resolve the matter in a reasonably expeditious manner:

Chair, Medical Sciences Inter-Divisional Research Ethics Committee;; Address: Research Services, University of Oxford, Wellington Square, Oxford OX1 2JD

Researcher: Dr Adrian Rohrbasser, Department of Primary Care Health Sciences in Oxford
Work: Santémed Gesundheitszentrum Wil, Friedtalweg 18, 9500 Switzerland, 0041719135400,
